# Subgrouping multimorbid patients with ischemic heart disease by means of unsupervised clustering: A cohort study of 72,249 patients defined comprehensively by diagnoses prior to presentation

**DOI:** 10.1101/2023.03.31.23288006

**Authors:** Amalie D. Haue, Peter C. Holm, Karina Banasik, Agnete T. Lundgaard, Victorine P. Muse, Timo Röder, David Westergaard, Piotr J. Chmura, Alex H. Christensen, Peter E. Weeke, Erik Sørensen, Ole B. V. Pedersen, Sisse R. Ostrowski, Kasper K. Iversen, Lars V. Køber, Henrik Ullum, Henning Bundgaard, Søren Brunak

**Author notes:** (SB). These authors contributed equally to this work (Amalie D. Haue, Peter C. Holm).

## Abstract

**Background:** There are no methods for classifying patients with ischemic heart disease (IHD) based on the entire spectrum of pre-existing diseases. Such methods might be clinically useful due to the marked differences in presentation and course of disease.

**Methods:** A population-based cohort study from a Danish secondary care setting of patients with IHD (2004-2016) and subjected to a coronary angiography (CAG) or coronary computed tomography angiography (CCTA). Data sources were The Danish National Patient Registry, in-hospital laboratory data, and genetic data from Copenhagen Hospital Biobank. Comorbidities included diagnoses assigned prior to presentation of IHD. Patients were clustered by means of the Markov Clustering Algorithm using the entire spectrum of registered multimorbidity. The two prespecified outcomes were: New ischemic events (including death from IHD causes) and death from non-IHD causes. Patients were followed from date of CAG/CCTA until one of the two outcomes occurred or end of follow-up, whichever came first. Biological and clinical appropriateness of clusters was assessed by comparing risks (estimated from Cox proportional hazard models) in clusters and by phenotypic and genetic enrichment analyses, respectively.

**Findings:** In a cohort of 72,249 patients with IHD (mean age 63.9 years, 63.1% males), 31 distinct clusters (C1-31, 67,136 patients) were identified. Comparing each cluster to the 30 others, seven clusters (9,590 patients) had statistically significantly higher or lower risk of new ischemic events (five and two clusters, respectively). 18 clusters (35,982 patients) had a higher or lower risk of death from non-IHD causes (12 and six clusters, respectively). All clusters at increased risk of new ischemic events, associated with risk of death from non-IHD causes as well. Cardiovascular or inflammatory diseases were commonly enriched in clusters (13), and distributions for 24 laboratory test results differed significantly across clusters. Clusters enriched for cerebrovascular diseases were generally not at increased risk of the two outcomes. Polygenic risk scores were increased in a total of 15 clusters (48.4%).

**Conclusions:** Clustering of patients with IHD based on pre-existing comorbidities identified subgroups of patients with significantly different clinical outcomes and presented a tool to rank pre-existing comorbidities based on their association with clinical outcomes. This novel method may support better classification of patients and thereby differentiation of treatment intensity depending on expected outcomes in subgroups.

## Introduction

Ischemic heart disease (IHD) is a common, chronic, and complex disease and mode of onset, disease burden and disease progression vary considerably between patients(1–3). This heterogeneity relates to several factors, but a major contribution is multimorbidity as more than 85% of IHD patients have been diagnosed with other chronic diseases; a phenomenon coined cardiometabolic multimorbidity(4,5). The increased mortality in patients with cardiometabolic multimorbidity is generally only related to single disease states, such as obstructive lung disease, diabetes, or stroke, although it is known that the risk of cardiovascular diseases is increased in many chronic, inflammatory disorders(6,7). As more patients at older age and with more and more co-morbidities are seen, new methods for characterizing and studying cardiometabolic multimorbidity are needed(8–12).

Unsupervised clustering algorithms can systematically reveal structures in large, feature-rich datasets and may be used to identify distinct patient subgroups within a heterogenous population(13). Proof-of-concept analyses of cardiovascular phenotypes, including IHD, heart failure, diabetes, and atrial fibrillation have already been performed(14–20). While these studies successfully identify subgroups resembling those from traditional analyses, they often fail to demonstrate that clustering analysis leads to novel understanding of a given dataset. Rather, they are typically restricted to characterize high-, medium-, and low-risk subgroups which by and large resemble more conservative approaches from an earlier, less data-rich, epoch(21).

For decades, Danish healthcare registries have had a strong position within epidemiological research(21–23). Given the opportunities for using clinical data more extensively, we carried out an unsupervised clustering analysis of 72,249 patients with IHD based on their entire disease history until IHD onset. Explicitly, we wanted to classify IHD based on the entire spectrum of multimorbidity. We identified distinct patient subgroups derived from a pool of 3,046 different diagnoses assigned prior to IHD onset. The biological and clinical factors characteristic to distinct patient subgroups identified by unsupervised clustering analysis were asserted by assessments of their associations with clinical outcomes and clinical characteristics, laboratory data, and genetics (Figure 1).

**Fig 1:**
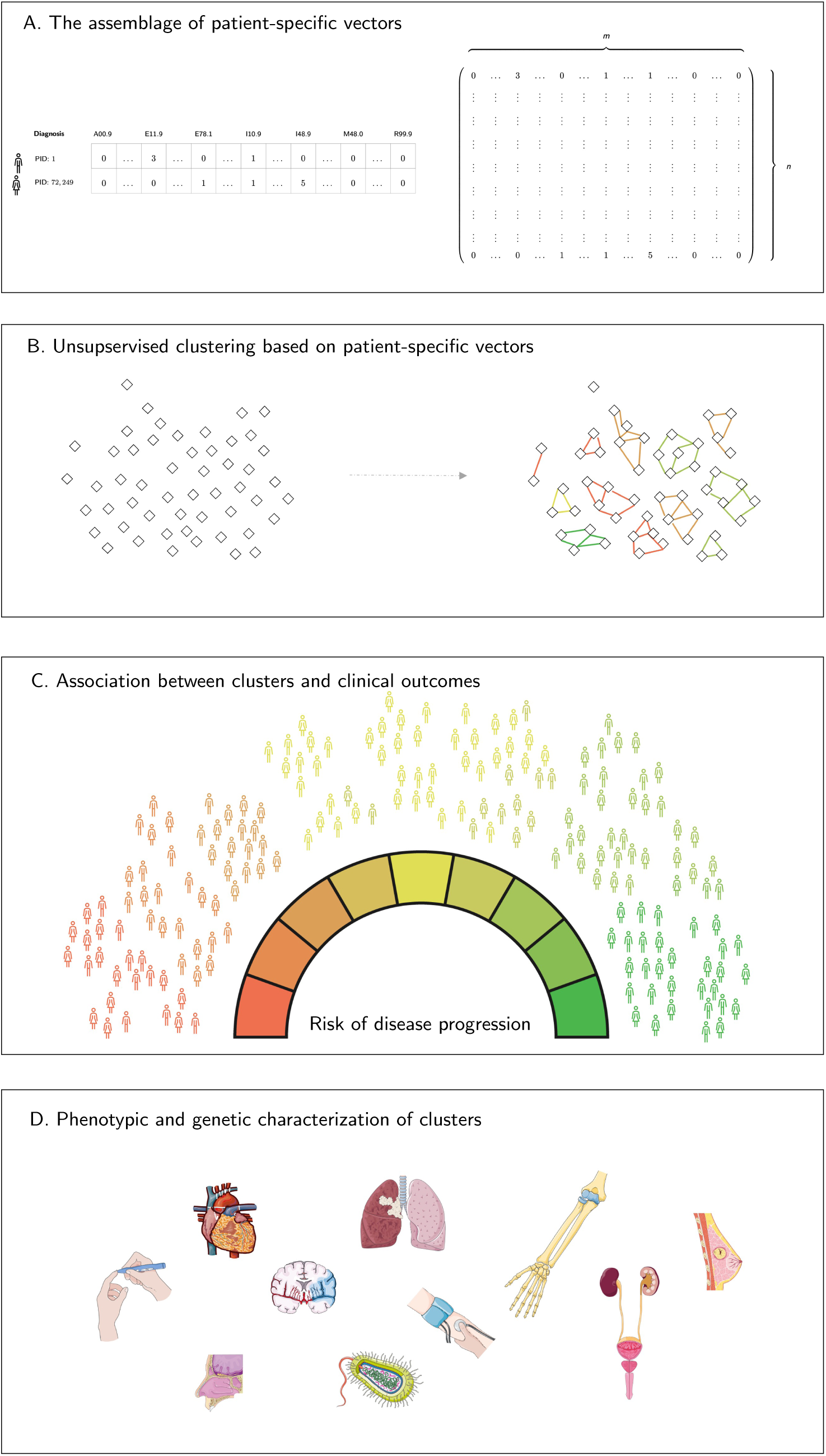
Graphical overview of study. Conceptual figure displaying the study design. A: Assemblage of patient-specific vectors that were the basis for construction of a matrix and an *n* x *m* matrix, where *n* corresponds to the number of included patients and *m* corresponds to the number of diagnoses. B: Unsupervised clustering of IHD patients using the MCL algorithm, which was the basis for performing unsupervised clustering to identify distinct clusters and associating them with clinical outcomes. C: Risk of disease progression (new ischemic events or death from non-IHD causes) in clusters. Color bar indicates increased, not altered, or decreased risk for patients in one cluster relative to the patients not in that cluster. D: Phenotypic and genetic characterization of clusters. Red: Increased risk of both outcomes. IHD: Ischemic heart disease. MCL: Markov Clustering.

## Methods

### Data sources, study population, and outcomes

Data from the Danish National Patient Registry (NPR) and the Danish Registry for Causes of Death were linked to in-hospital electronic health data covering the two Danish healthcare regions in Eastern Denmark (∼2.9 mil inhabitants), and the Copenhagen Hospital Biobank Cardiovascular Disease Cohort(22,24,25). Linkage of different healthcare data sources was obtained via the personal identification number and only patients admitted to a hospital in Eastern Denmark in years 2004 to 2016 were considered(26). We identified all patients in NPR who were assigned an ICD-10 code for IHD(27). To increase the positive predictive value of IHD diagnoses and align included patients in time, we further required that patients had been subjected to coronary arteriography (CAG) or coronary computed tomography angiography (CCTA). To qualify that CAG/CCTAs were conclusive for IHD, patients were only included if the CAG/CCTA was performed during a contact where patients were assigned an ICD-10 code for IHD. We set the earliest CAG/CCTA fulfilling this criterium as the index date and excluded patients with an index date before year 2004 or after 2016 (Fig 2).

**Fig 2:**
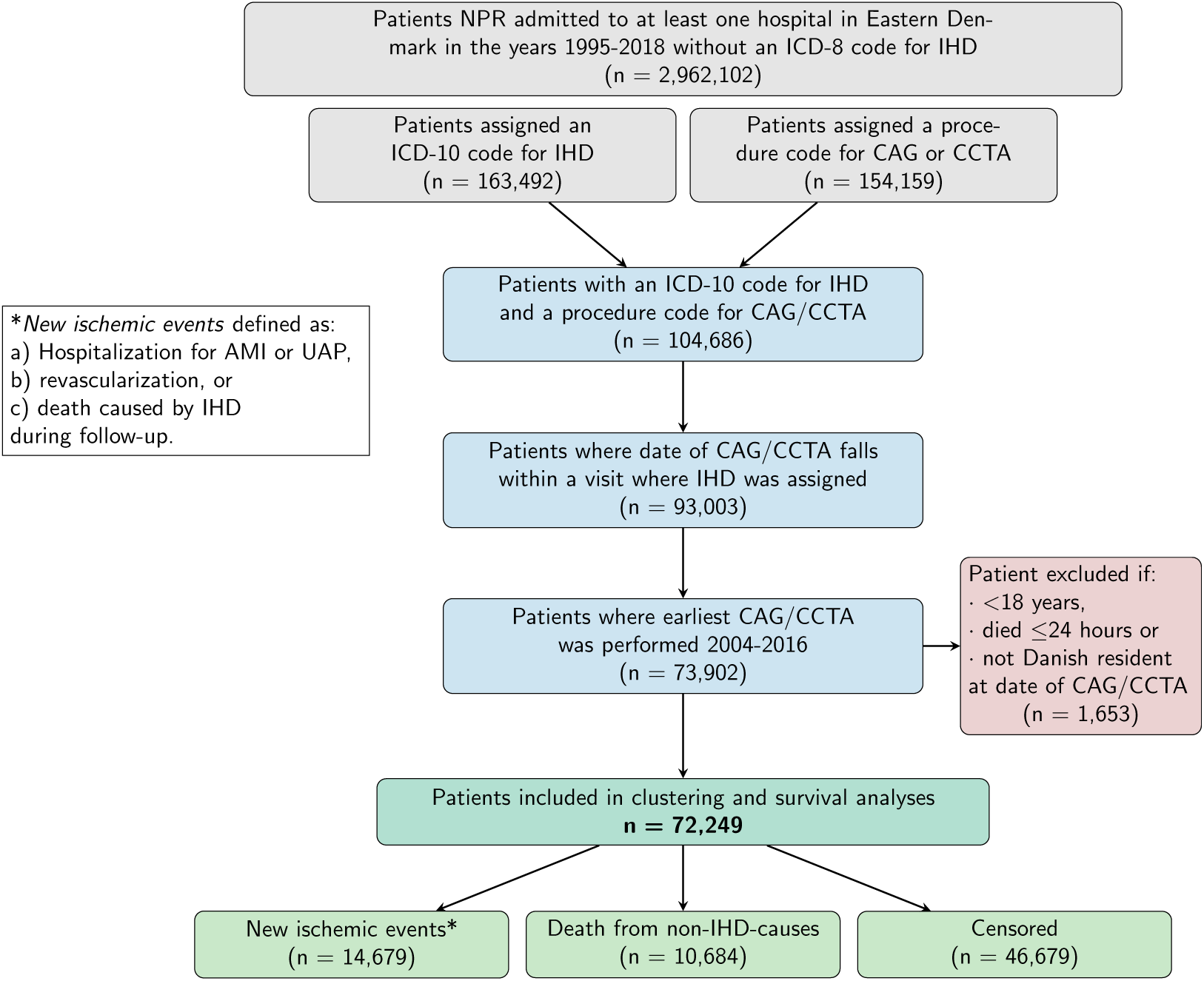
Flowchart: Data sources, study population, and outcomes. Gray: Identification. Blue: Screening. Red: Eligibility. Green: Inclusion and outcomes. AMI: Acute myocardial infarction. UAP: Unstable angina pectoris. NPR: The Danish National Patient Registry. IHD: ischemic heart disease (ICD-10 codes I20-I25). CAG: Coronary arteriography. CCTA: Coronary computed tomography angiography. ICD-10: International Statistical Classification of Diseases and Related Health Problems 10^th^ Revision. SKS: Sundhedsvæsenets Klassifikationssystem (The Danish Health Authority Classification System).

There were two predefined outcomes: 1) New ischemic events and 2) Death from other causes than IHD (non-IHD causes). The outcome “new ischemic event” was a composite outcome of a) hospitalization minimum 30 days after index for myocardial infarction or unstable angina pectoris (i.e., hospitalization with myocardial infarction or unstable angina pectoris as the primary diagnosis), b) revascularization not related to the index date, and c) any death where IHD was listed as the primary or secondary cause. Outcomes were obtained from NPR and Danish Registry for Causes of Death. Eligible codes for inclusion, outcomes and specific cutoffs are available in S1 Fig and S1 Table.

### Data preprocessing and application of the Markov cluster algorithm

We performed a clustering analysis of included patients based on their multimorbidity prior to their IHD diagnosis (index) using the Markov cluster (MCL) algorithm(28). Multimorbidity was represented as patient-specific vectors using diagnoses assigned prior to or at index. ICD-10 codes assigned to less than five patients (n=1,673) were excluded from the analysis. As we focused the studies on multimorbidity in IHD, ICD-10 codes for IHD (I20-I25) were excluded from patients-specific vectors. Thus, a total of 3,046 ICD-10 codes were the basis for constructing a patient similarity network that was used as MCL algorithm input. Patient-specific vectors of length 3,046 with integers indicating the number of times a patient had been assigned a particular ICD-10 code. The length of the vectors corresponded to the number of input features (ICD-10 codes). By combining the patient-specific vectors from all included patients, a matrix of size *n* x *m* was constructed, where *n* indicates the number of included patients and *m* indicates the number of input features (ICD-10 codes). Following a series of preprocessing steps described in S1 Appendix, a patient similarity network was created based on the *n* x *m* matrix and used as input for the MCL algorithm(29). Resulting clusters were denoted *C* followed by an integer indicating the rank of the clusters with respect to cluster size (number of patients in that cluster). Thus, C1 denotes the largest cluster and cluster-membership was used to denote a cluster as a covariate in subsequent analyses. Robustness of clustering was assessed by generating a series of diluted and shuffled versions of the resulting clusters (reference clustering), and their similarity was quantified using the variance of the information measure as previously described(30). Explicitly, a series of diluted and shuffled versions of the input graph were generated(31). In total, 20 variations of the input graph were constructed by shuffling and deleting edges, respectively. The variation in the graphs was then quantified by means of variation of the information meassure. Details regarding the MCL settings and a description of cluster robustness assessment are available in the S1 Appendix.

### Preprocessing of laboratory and genetic data

Clusters were characterized by laboratory and genetic data based on the subset of patients where these data types were available. A panel of 25 different lab parameters was included in the analyses. Only tests taken up to 90 days before index or at the day of index were included. Included lab tests were plasma levels of potassium, sodium, hemoglobin, estimated glomerular filtration rate (eGFR), creatinine, carbamide, glucose, troponin (I/T), HDL cholesterol, LDL cholesterol, total cholesterol, leukocytes, C-reactive protein, lymphocytes, monocytes, neutrophils, basophiles, platelets, INR, alanine transaminase, albumin, alkaline phosphatase, bilirubin, and triglyceride. For every cluster, a *score* was computed based on the number of patients with a lab test below, within, or above the standard reference value, indicated by –1, 0 and 1, respectively. The *score* was defined as the mean of the summarized values per cluster.

Autosomal genotype data were obtained by identifying included patients who were also among the study participants in the Copenhagen Hospital Biobank – Cardiovascular Disease Cohort(25). For included patients with genetic data available, we calculated polygenic risk scores (PRSs) for 14 traits, obtained from nine GWAS meta-analyses (atrial fibrillation, BMI-adjusted non-insulin diabetes, chronic kidney disease, HDL cholesterol levels, heart failure, LDL cholesterol levels, stroke, total cholesterol levels, triglyceride levels) and five GWAS (acute myocardial infarction, coronary artery disease, diastolic blood pressure, non-alcoholic fatty liver disease, systolic blood pressure)(38–41). PRSs were calculated using the “LDpred2-auto” algorithm, implemented in the R package “bigsnpr” (version 1.11.6) with R version 4.0.0 and the workflow management system Snakemake(42–44). Each trait’s PRS distribution was scaled to a mean of zero and a standard deviation of one.

### Statistical analyses of clusters identified by the MCL algorithm

As the study was designed to identify patient subgroups and not individual variation, clusters of size < 500 were excluded from the remaining analyses. Mean age at IHD onset in each cluster was compared to the mean age at onset in all the other clusters using Tukey’s Honest Significant Difference (HSD) method. Significance level was set to 0.05 and P-values were adjusted using the Holm method assuming 465 tests (adj. P-val.).

To investigate the association between cluster-membership and the competing risks of new ischemic events and death from non-IHD causes, we used Cox proportional-hazards models (Cox models). Patients were followed from index until occurrence of either of the two outcomes, or end of follow-up (year 2018), whichever came first. The dependent variable was either risk of new ischemic events or death from non-IHD causes, and the independent variables were cluster, sex, and age at index. To age-adjust the models, analyses were performed using restricted cubic spline with three knots for age at index. Follow-up time was truncated to a maximum of five years. For each cluster, hazard ratios (HRs) and 95% confidence intervals (CIs) were estimated by comparing HRs for the members of the cluster with the HRs with that of non-members.

Further characterization of clusters consisted of: (1) phenotypic enrichment analysis, (2) characterization of clusters with respect to their laboratory profiles and (3) a test for genetic enrichment. The phenotypic enrichment analysis was carried out based on ratios between observed (O) and expected (E) frequencies of diagnoses in the clusters (O/E-ratios). That is, ratios between the frequencies of ICD-10 codes in each cluster (observed frequencies) and the frequencies of ICD-10 codes in the entire population (expected frequencies) were calculated and expressed as O/E-ratios(45). In subsequent characterization of clusters, enrichment denoted O/E-ratios > 2, and clusters were characterized as having little enrichment if the sum of the ten largest O/E-ratios < 50. Inverse changes were used to denote O/E-ratios between 0 and 1.

Hierarchical clustering was applied to estimate the cluster similarity with respect to the laboratory tests using the Euclidean distance between the *score* of each cluster for each test.

For each of the fourteen traits we calculated PRSs for, we used Wilcoxon rank-sum tests to compare the PRS distribution of each cluster to the combined PRS distribution of PRSs in all other clusters. Resulting P-values were converted to the false discovery rate (FDR) to account for multiple testing, with a total of 434 tests. We report effect sizes as calculated by the “wilcox.test” function built into R version 4.0.0. Level of significance was set to FDR < 0.05, assuming 434 tests.

Further details regarding preprocessing and analyses of laboratory and genetic data are available in the S2 Appendix.

## Results

### Cohort demographics and co-morbidities

A total of 72,249 patients (63.1% males, mean age 63.9 years) were included (Table 1). Angina pectoris (I20) was the most common IHD diagnosis (38,239 patients, 52.9%), followed by acute myocardial infarction (I21) (33,229 patients, 46.0 %) and chronic IHD (I25) (22,750 patients, 31.5%). The most common co-morbidity prior to the IHD index was hypertension (I10.9) (24,818 patients, 34.4%) followed by dyslipidemia (E78.0) (12,780 patients, 17.7%) and non-insulin dependent diabetes (E11.9) (7,551 patients, 10.5%). Prior to index, the mean number of diagnoses per patient was 8.1. A total of 68,103 patients (94.3%) had co-morbidities registered prior to index. The overall incidence (new ischemic events and death from non-IHD causes) was 94 events per 1000 person-years (Table 1).

**Table 1:** Patient demographics, co-morbidities, and outcomes.

| **Cohort demographics** | **Total** | **Males** | **Females** |
| --- | --- | --- | --- |
| Number of patients (%) | 72,249 | 45,576 (63.1) | 26,673 (36.1) |
| Mean age at index (SD) | 63.9 (11.9) | 62.9 (11.6) | 65.6 (12.1) |
| **IHD manifestations (ICD-10)** | **Total** | **Males** | **Females** |
| Angina pectoris (I20) | 38,239 | 22,628 | 15,611 |
| Acute myocardial infarction (I21) | 33,299 | 27,720 | 10,579 |
| Subsequent myocardial infarction (I22) | 61 | 34 | 27 |
| Certain current complications following acute myocardial infarction (I23) | 138 | 92 | 46 |
| Other acute ischemic heart diseases (I24) | 1,341 | 814 | 527 |
| Chronic ischemic heart disease (I25) | 22,750 | 14,589 | 8,152 |
| **Common comorbidities (ICD-10)** | **Total** | **Males** | **Females** |
| Primary (essential) hypertension (I10.9) | 24,818 | 14,508 | 10,310 |
| Hypercholesterolemia (E78.0) | 12,780 | 7,842 | 4,938 |
| Non-insulin dependent diabetes (E11.9) | 7,551 | 4,891 | 2,660 |
| Atrial fibrillation and atrial flutter, unspecified (I48.9) | 7,075 | 4,509 | 2,566 |
| Heart failure, unspecified (I50.9) | 6,160 | 4,059 | 2,101 |
| Chest pain, unspecified (R07.9) | 5,863 | 3,441 | 2,422 |
| Senile cataract, unspecified (H25.9) | 5,764 | 2,795 | 2,969 |
| Pneumonia, unspecified (J18.9) | 5,469 | 3,236 | 2,260 |
| Hyperlipidaemia, unspecified (E78.5) | 5,002 | 3,306 | 1,696 |
| Chronic obstructive pulmonary disease (J44.9) | 4,621 | 2,449 | 2,172 |
| **Outcomes, number of cases** | **Total** | **Males** | **Females** |
| New ischemic events (%) | 14,679 | 10,152 | 4,527 |
| ■ Myocardial infarction | 5,833 | 3,709 | 2,124 |
| ■ Revascularization | 6,282 | 4,718 | 2,124 |
| ■ Death caused by IHD | 2,563 | 1,724 | 839 |
| Death from non-IHD causes (%) | 10,684 | 6,710 | 3,974 |
| Censored (%) | 46,886 | 28,713 | 18,172 |
| **Outcomes, time to event** | **Mean time to event in years (SD)** |  |  |
|  | **Total** | **Males** | **Females** |
| New ischemic events | 1.48 (1.40) | 1.49 (1.41) | 1.48 (1.40) |
| ■ Myocardial infarction | 2.40 (1.87) | 2.41 (1.89) | 2.38 (1.85) |
| ■ Revascularization | 2.25 (1.88) | 2.28 (1.89) | 2.16 (1.84) |
| ■ Death caused by IHD | 1.92 (1.13) | 1.95 (2.02) | 1.88 (2.05) |
| Death from non-IHD causes | 2.16 (1.50) | 2.14 (1.49) | 2.20 (1.51) |
| Censored | 4.37 (1.08) | 4.36 (1.09) | 4.39 (1.06) |
| Total | 3.72 (1.64) | 3.67 (1.67) | 3.81 (1.60) |

### Unsupervised clustering of multimorbid patients with IHD

In the cohort, the MCL algorithm identified 36 distinct clusters based on the set of 3,046 ICD-10 codes assigned to the patients prior to or at index. The 36 clusters contained a total of 68,084 patients. Expectedly, the remaining 4,365 patients (6.0% of included patients) that did not cluster were primarily patients with no diagnoses prior to index (>99%). Further, cluster robustness was assessed as described in Methods, where the variation of information measure less than 2 if 25% of the edges in the input graph were deleted or shuffled (S4 Figure). Next, the 31 of the 36 clusters with >500 patients (67,136 patients) were characterized (Table 2). Using Tukey’s HSD to compare the age at index between all 31 clusters (a total of 466 combinations), we found significant differences in 391 comparisons (84.1%, S3 Table). For demographics of patients that did not cluster or were in clusters of size < 500, see S4 Table.

**Table 2:** Cluster demographics, characteristics, and associations with outcomes.

| Cluster | Size | Mean age at index in years (SD) | Males | Females | New ischemic events |  | Death from non-IHD causes |  |
| --- | --- | --- | --- | --- | --- | --- | --- | --- |
|  |  |  |  |  | HR | Adj. P-val. | HR | Adj. P-val. |
| C1 | 7,191 | 64.8 (11.3) | 3,897 | 3,294 | 1.000 | > 0.050 | 0.856 | > 0.050 |
| C2 | 5,990 | 58.6 (11.5) | 2,862 | 3,127 | 0.825 | < **0.001** | 0.600 | < **0.001** |
| C3 | 4,641 | 56.8 (11.4) | 2,727 | 1,914 | 0.757 | < **0.001** | 0.586 | < **0.001** |
| C4 | 4,401 | 69.6 (10.2) | 2,853 | 1,548 | 0.920 | > 0.050 | 1.461 | < **0.001** |
| C5 | 4,290 | 63.9 (10.7) | 2,803 | 1,487 | 1.402 | < **0.001** | 1.629 | < **0.001** |
| C6 | 3,589 | 59.7 (10.9) | 2,388 | 1,201 | 0.969 | > 0.050 | 0.675 | < **0.001** |
| C7 | 3,309 | 63.8 (11.0) | 2,025 | 1,284 | 0.889 | > 0.050 | 0.611 | < **0.001** |
| C8 | 2,802 | 71.1 (10.9) | 1,867 | 935 | 0.943 | > 0.050 | 0.842 | > 0.050 |
| C9 | 2,581 | 63.7 (11.8) | 1,803 | 778 | 1.314 | < **0.001** | 1.789 | > 0.050 |
| C10 | 2,562 | 74.2 (9.6) | 1,225 | 1,337 | 0.978 | > 0.050 | 0.928 | > 0.050 |
| C11 | 2,292 | 66.1 (11.0) | 2,186 | 106 | 0.926 | > 0.050 | 0.650 | < **0.001** |
| C12 | 2,213 | 70.3 (10.2) | 2,068 | 145 | 0.920 | > 0.050 | 0.805 | > 0.050 |
| C13 | 2,070 | 58.6 (10.2) | 1,348 | 722 | 0.946 | > 0.050 | 0.577 | < **0.050** |
| C14 | 2,070 | 68.2 (9.6) | 1,030 | 1,010 | 1.146 | > 0.050 | 3.390 | < **0.001** |
| C15 | 2,040 | 63.9 (10.1) | 1,208 | 805 | 1.031 | > 0.050 | 0.784 | > 0.050 |
| C16 | 1,654 | 64.1 (12.1) | 1,013 | 641 | 1.107 | > 0.050 | 1.761 | < **0.001** |
| C17 | 1,281 | 65.3 (9.9) | 714 | 567 | 1.001 | > 0.050 | 1.761 | < **0.001** |
| C18 | 1,251 | 68.2 (9.8) | 802 | 449 | 1.790 | < **0.001** | 3.421 | < **0.001** |
| C19 | 1,168 | 58.5 (9.7) | 995 | 173 | 0.752 | > 0.050 | 1.571 | > 0.050 |
| C20 | 1,119 | 71.5 (11.3) | 713 | 406 | 1.213 | > 0.050 | 1.782 | < **0.001** |
| C21 | 1,000 | 61.0 (11.0) | 769 | 231 | 1.116 | > 0.050 | 0.890 | > 0.050 |
| C22 | 988 | 69.2 (10.4) | 516 | 472 | 1.023 | > 0.050 | 0.978 | > 0.050 |
| C23 | 935 | 58.7 (12.2) | 588 | 347 | 1.609 | < **0.001** | 2.275 | < **0.001** |
| C24 | 932 | 67.9 (10.1) | 28 | 904 | 0.787 | > 0.050 | 1.589 | < **0.001** |
| C25 | 860 | 56.2 (9.9) | 664 | 196 | 0.978 | > 0.050 | 2.691 | < **0.001** |
| C26 | 852 | 58.7 (12.1) | 391 | 461 | 0.939 | > 0.050 | 1.108 | > 0.050 |
| C27 | 823 | 65.1 (10.9) | 532 | 291 | 1.201 | > 0.050 | 1.289 | > 0.050 |
| C28 | 686 | 71.7 (8.0) | 673 | 13 | 0.866 | > 0.050 | 1.786 | < **0.001** |
| C29 | 550 | 57.2 (11.1) | 435 | 115 | 0.906 | > 0.050 | 0.985 | > 0.050 |
| C30 | 533 | 61.2 (11.7) | 391 | 172 | 1.874 | < **0.001** | 5.364 | < **0.001** |
| C31 | 520 | 64.4 (11.2) | 213 | 307 | 1.052 | > 0.050 | 1.484 | > 0.050 |
| NA* | 5,113 | 60.1 (11.1) | 3,878 | 1,235 | NA | NA | NA | NA |

\*Patients that did not cluster or were in clusters of size < 500

### Clusters, clinical outcomes, and phenotypic enrichment

To assess if the unsupervised clustering identified patient subgroups at different risks of disease progression, we used cluster-membership (C1-C31) as a covariate in a series of Cox models. A total of 14,679 patients experienced a new ischemic event during follow-up and 10,684 patients died from other causes than IHD. Mean follow-up time was 3.72 years (Table 1). Risks for new ischemic events and death from non-IHD causes in each cluster were compared to the pooled risk for patients in the remaining 30 clusters. The survival analysis demonstrated that the MCL algorithm stratified patients according to risk of new ischemic events and death from non-IHD causes (Fig 3). Comparing each cluster (n=1) to all the others (n=30), a total of seven clusters (20,221 patients) had a statistically significantly higher or lower risk of new ischemic events (Adj. P-val. < 0.05). Five clusters (9,590 patients) and two clusters (10,631 patients) were at increased and decreased risk of new ischemic events, respectively. Similarly, a total of 18 clusters (43,173 patients) had a statistically significantly higher or lower risk of death from non-IHD causes (Adj. P-val. < 0.05); where 14 clusters (21,282 patients) and four clusters (21,891 patients) were at increased or decreased risk of death from non-IHD causes. All clusters at increased risk of new ischemic events, associated with risk of death from non-IHD causes as well. The same was true for the two clusters at decreased risk of new ischemic events, i.e., these clusters were at decreased risk of death from non-IHD causes as well. A total of 13 clusters, (23,963 patients) were not have altered risk of the two outcomes, when compared to the other clusters (Table 2).

**Fig 3:**
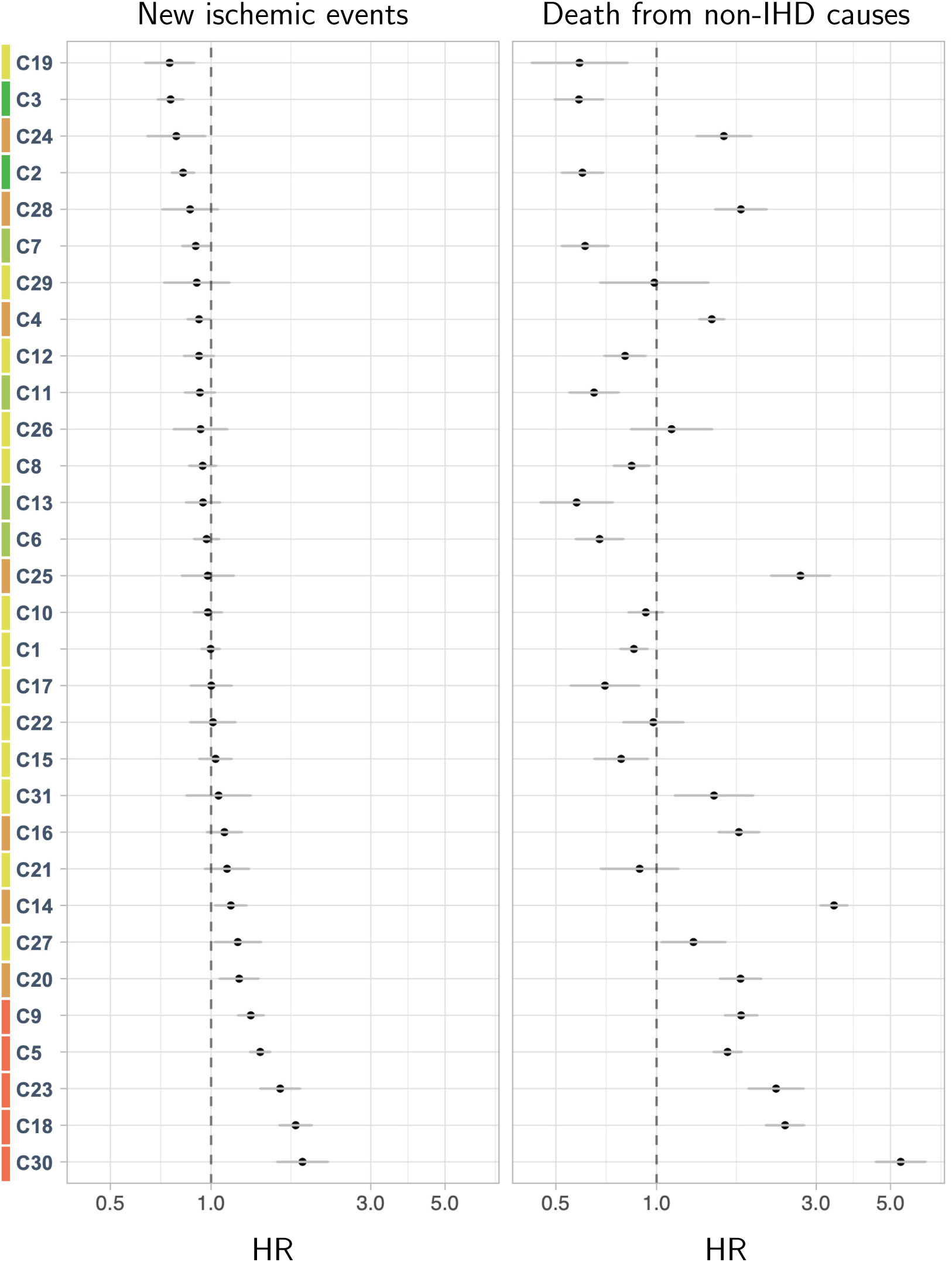
Risk of new ischemic events and non-IHD causes stratified by cluster. Forest plots where clusters are shown against HR for new ischemic events (left) and death from non-IHD causes (right). X-axis: HR for a single cluster relative to mean HR of the 30 other clusters. Y-axis: Clusters arranged by risk of new ischemic events, increasing risk from top to bottom. Colors indicating significance. Dark green: Reduced risk of new ischemic events and death from non-IHD causes. Lighter green: Reduced risk of death from non-IHD causes. Yellow: No significance. Orange: Increased risk of death from non-IHD causes. Red: Increased risk of new ischemic events and increased risk of death from non-IHD causes. IHD: Ischemic heart disease. HR: Hazzard ratio.

The distribution of O/E-ratios was heavily left-skewed as less than 99% (n=101) of all O/E-ratios were >10 and roughly 7% (n=887) of all O/E-ratios were >2. About 60% of all O/E-ratios (n=8,056) were in the range of 0 and 1 corresponding to inverse changes. Generally, clusters that had high risk of new ischemic events or death from non-IHD causes were also characterized by large, summarized O/E-values corresponding to a high degree of multimorbidity (S5 Table 5). The results of the enrichment analysis were summarized according to nine different disease categories: (1) diabetes mellitus, (2) cardiac diseases, (3) diseases affecting the upper airways, (4) cerebrovascular diseases, (5) infections and other acquired diseases, (6) gynecologic diseases, (7) Inflammatory and degenerative of the musculoskeletal system, (8) diseases of the urinary system, and (9) hypertension (Fig. 4).

**Fig 4:**
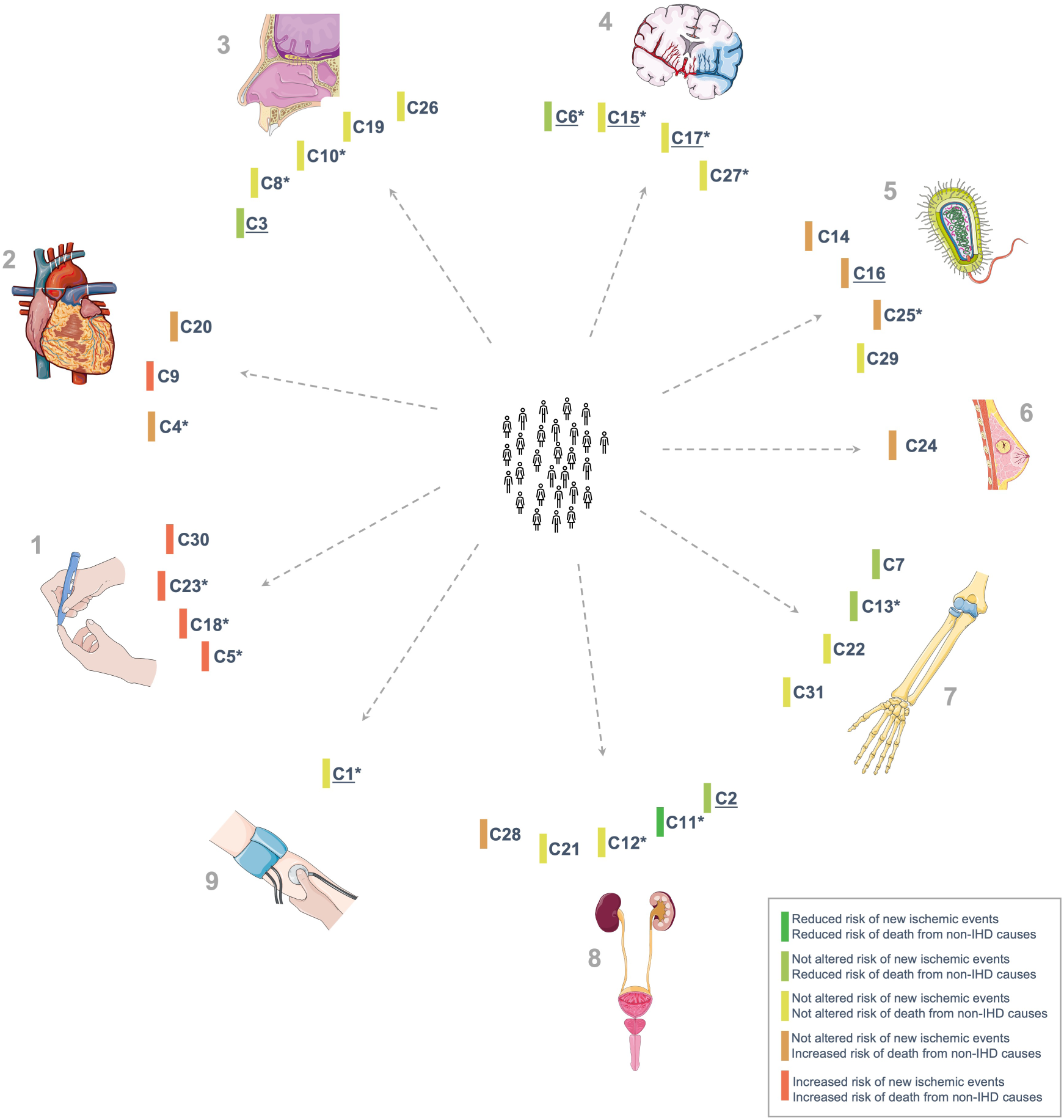
Infographic summarizing the results of the study. Center: Study cohort. Periphery: Graphical overview of results from clustering analysis, survival analysis and characterization of clusters. Arrows indicate disease categories (for details, see text). 1: Diabetes mellitus. 2: Cardiac diseases. 3: Diseases affecting the upper airways. 4: Cerebrovascular diseases. 5: Infections and other acquired diseases. 6: Gynecologic diseases. 7: Inflammatory and degenerative of the musculoskeletal system. 8: Diseases of the urinary system. 9: Hypertension. C1-31: Clusters. “Underline” indicates little enrichment. “*” indicates genetic enrichment. For underlying data, see S5 and S7 Tables.

An in-depth characterization of clusters enriched for cardiometabolic or-vascular diseases, degenerative or inflammatory diseases and clusters characterized by little enrichment and inverse changes is provided in the following paragraphs.

### Clusters enriched for cardiometabolic and-vascular diseases

Four of the five clusters at increased risk of new ischemic events (and death from non-IHD causes) were enriched for diabetes (C5, C18, C23, and C30). In these four clusters, HRs ranged from 1.40 (C5, 95%CI: 1.30;1.50, adj. P-val. < 0.001) to 1.88 (C30, 95%CI: 1.60;2.00, adj. P-val. < 0.001) with a significant difference in age at index (C5: 63.9 years, C30: 61.2 years, Adj. P-val. < 0.001, TukeyHSD). C18 and C23 were only enriched for insulin-dependent diabetes, but differed in that C18 was also enriched for insulin-dependent diabetes with vascular complications and periphery atherosclerosis. In contrast, C5 was only enriched for non-insulin dependent diabetes and included diabetes with as well as without complications. Lastly, C30 was only enriched for diabetes with complications (insulin and non-insulin dependent) and was the diabetes cluster enriched for chronic kidney disease and bacterial infections, as well (S5 Table 5).

Other cardiac diseases that displayed enrichment were supraventricular arrythmias (C4), cardiomyopathies (C9), and valve diseases (C20). Of the three clusters, only C9 had increased risk of new ischemic events (HR: 1.31 (C9, 95%CI: 1.20;1.44, Adj. P-val: < 0.001). Risk of death from non-IHD causes was 1.79 (95%CI: 1.60;2.00, adj. P-val. < 0.001). In contrast, C4 and C20 only had increased risk of death from non-IHD causes with HRs of 1.49 (C4, 95%CI: 1.34;1.59, adj. P-val. < 0.001) and 1.78 (C20, 95%CI: 1.54;2.04, adj. P-val. < 0.001). Interestingly, the cluster enriched for cerebrovascular diseases (C27) did not have altered risk of any of the two outcomes. In sum, all clusters that had increased risk of new ischemic events were enriched for cardiometabolic diseases, albeit not all clusters enriched for cardiometabolic and-vascular diseases had increased risk of new ischemic events (Table 2 and S5 Table 5).

### Clusters enriched for degenerative or inflammatory diseases

Six clusters (C7, C13, C14, C22, C26, and C31) were enriched for diagnoses describing degenerative or inflammatory diseases, i.e., osteoarthritis (C7), degenerative spine disease (C13 and C22), chronic obstructive pulmonary disease (C14), asthma (C26), and rheumatoid arthritis (C31). Remarkably, none of the four clusters had increased risk of new ischemic events and only one cluster (C14) had increased risk of death from non-IHD causes (HR: 3.39, 95%CI: 3.09;3.71, adj. P-val. < 0.001). Conversely, C7 and C13 had reduced risk of death from non-IHD causes (C7, HR: 0.61, 95%CI: 0.52;0.72, adj. P-val. < 0.001 and C13, HR: 0.58, 95%CI: 0.45;0.74, adj. P-val. < 0.001). Age at index for the clusters enriched for degenerative or inflammatory diseases range between 58.6 years (C13) and 69.2 years (C22) (Table 2). Taken together, these findings hint to the dual nature of inflammation as a potential disease modifier as well as a risk factor.

### Clusters characterized by little enrichment and inverse changes

Six clusters (C1, C2, C3, C6, C15, and C17) were characterized by little enrichment, which included the two clusters with reduced risk of new ischemic events (C2, HR: 0.82, 95%CI: 0.76;0.89, adj. P-val. < 0.001 and C3, HR: 0.76, 95%CI: 0.52;0.69, adj. P-val. < 0.001). Not surprisingly, none of these six clusters had increased risk of either of the two outcomes, but three clusters (C2, C3, and C6) had reduced risk of death from non-IHD causes (C2, HR: 0.60, 95%CI: 0.52;0.69, adj. P-val. < 0.001, C3, HR: 0.59, 95%CI: 0.59;0.69, adj. P-val. < 0.001 and C6, HR: 0.68, 95%CI: 0.57;0.79, adj. P-val. < 0.001) (Table 2). It was a common attribute of the clusters without altered risk of any of the two outcomes that O/E-ratios for hypertension and dyslipidemia were among the largest. In contrast, diabetes, heart failure, and chronic obstructive pulmonary disease frequently displayed inverse changes (O/E-ratios < 1) in these clusters (S5 Table). Taken together, these observations indicate that risk of disease progression in this populations necessitates a more sophisticated analysis of multimorbidity.

For a list with results of the enrichment analysis for all clusters, including the 13 clusters not described above, S5 Table 5.

### Clusters and their association with laboratory measurements and genetic data

Clusters were also characterized by means of datatypes not included among the MCL algorithm input features. For patients in the 31 clusters, we had laboratory measurements on 30,755 (49.5%) and genetic data on 19,422 (31.3%). To assess if the phenotypic differences captured by the MCL algorithm were also reflected in laboratory measurements, we tested if the distributions of test results within and out of reference ranges differed significantly. There were significantly different distributions of tests within and out of reference ranges in clusters for the 24 most frequent tests. Overall, this indicates that the phenotypic patterns within the entire spectrum of cardiovascular multimorbidity registered before index correlate with results of clinical laboratory tests (S6 Table). Thus, these findings are a strong indicator that the patterns captured by the MCL algorithm are biologically relevant. For a graphical summary of the laboratory scores in each cluster, see S5 Figure.

Finally, we identified 41 cases (out of 434 tests) where the PRS distribution for a specific trait in a cluster was significantly different from that trait’s combined PRS distribution of the other 30 clusters. Among these cases, we found the largest effects size to be a higher genetic risk for atrial fibrillation in cluster C4 (0.57, FDR < 0.001) as well as a higher genetic risk for non-insulin dependent diabetes in cluster C5 (0.55, FDR < 0.001). These findings are congruent with the results of the enrichment analysis for C4 and C5, respectively. In contrast, C1 (phenotypically characterized by inverse changes) had relatively large, positive effect sizes for systolic as well as diastolic blood pressure (0.20 and 0.16, FDR < 0.001). Similarly, there were positive effect sizes for total cholesterol and triglycerides in C6, which was also characterized by little phenotypic enrichment as well as a high degree of inverse changes. A list of significant effect sizes for the 41 significant cases, see S7 Table.

## Discussion

In this study, we developed a novel, data-driven method for structuring the entire spectrum of multimorbidity by means of an unsupervised clustering analysis. In a cohort of 72,249 patients with IHD patients, we identified 31 distinct clusters (67,136 patients) based on 3,046 diagnoses assigned prior to or at index. By comparing risk of new ischemic events and death from non-IHD causes across clusters and then performing an enrichment analysis, we found that clusters at increased risk of new ischemic events were enriched for diabetes (four clusters) or cardiomyopathies (one cluster). Neither the cluster enriched for supraventricular arrythmias, nor valve diseases had increased risk of new ischemic events. Degenerative and inflammatory diseases were enriched in a total of six clusters and displayed no clear trend in their relation to the outcomes. The results of the enrichment analysis were supported by trends in laboratory test results and clusters enriched for supraventricular arrythmias and non-insulin diabetes also had congruently, higher genetic risks.

The results of the study agree with common knowledge on risk of IHD, while also adding insights to the disease-diseases associations, which are currently underappreciated in the literature. The fact that clusters enriched for diabetes were generally the most high-risk clusters serves as a methodological reality check(6). Added value of the study lies in the fact that the method allows for a more sophisticated description of such associations, as the method allows to study the entire spectrum of multimorbidity. For example, four clusters were enriched for diabetes, which is in line with the current paradigm that a single term is insufficient to describe a multifactorial disease, such as diabetes(17,30). By integrating different data types, the findings indicate how phenotypic and genetic data complement each other, by exemplifying (1) that clustering analysis facilitates stronger genetic signals in patient subgroups and (2) that genetic data may unveil patterns not captured by phenotypic data alone.

In addition, the method developed in this study and subsequent findings add perspective to the relatively limited body of literature regarding associations between chronic inflammatory and cardiovascular diseases(7). While previous studies have concluded that the risk of cardiovascular diseases is increased in most chronic inflammatory disorders, the results of our study indicate that pre-existing degenerative or inflammatory disorders in patients with IHD do not increase the risk of new ischemic events.

The pre-selected outcomes in the present study are also a unique aspect of the study, as previous clustering analyses within the cardiovascular domain studies have mainly analyzed all-cause mortality(18,19). This aspect of the study allows to distinguish between risk of progression related to IHD and risk of progression that is related to comorbidity drawing attention to important aspects of multimorbidity in this domain. For example, clusters enriched for supraventricular arrythmias and chronic obstructive pulmonary disease, respectively, only had increased risk of death from non-IHD causes. The study design, including the enrichment analysis, also revealed that classical risk factors for IHD (e.g., hypertension and dyslipidemia) did not drive the clustering. This finding agrees with previously published comorbidity phenotypes in patients with IHD(19). We argue that the present study displays that continuous exploration and characterization of multimorbidity in IHD are key elements in optimizing the exploit the full potential of continuously developing treatment strategies.

Previous clustering analyses within the cardiovascular domain have typically included either thousands of patients or hundreds of input features, but not both(15,16). For example, Hall et al. defined multimorbidity using only eight different chronic conditions, whereas Crowe et al. defined multimorbidity with reference to 20 predefined conditions(18,19). Thus, the scale of our study exceeds that of previous work, as it includes more than 70,000 patients and more than 3,000 input features. And further, we limited the risk of introducing bias by not exerting feature selection prior to clustering.

The two main limitations with respect to the data foundation are that (1) owing to the novelty of the method, there were no standardized way of assessing the representation of multimorbidity and (2) it was only a subset for which laboratory and genetic data were available. These challenges are naturally overcome in clustering analyses based on data from randomized controlled trials, such as the studies by Inohara et al, and Karwath et al.(16,20) However, in the present, data-rich era, we argue that it is highly important to develop methods for structuring and studying other data than what is being collected for trials. Ideally, the two approaches, based on nationwide data and randomized controlled trials, respectively, will complement each other; and will facilitate more precise identification of patients who are likely to benefit from different treatment options as well as guide optimized selection of patients for randomized controlled trials.

In conclusion, the study further showcases the strengths of a more fine-grained analysis of patient subgroups, which, in turn, may pave the way for successful implementation of precision medicine. Owing to its flexibility, the comprehensive, data-driven analysis of cardiovascular multimorbidity represents a novel method for characterizing multimorbidity in IHD with great potential of applying it to other diseases of interest or other clinical data. Such trends may guide clinical decision making in cases, where for example it is not obvious how to manage the angiographic findings or the combination of drugs that a specific patient will benefit most from.

In conclusion, the present study cements the complexity of multimorbid patients with IHD and exemplifies the clinical relevance of a more fine-grained patient subgrouping by carrying out a cluster-based risk-stratifying the cohort. Further, owing to its flexibility, the comprehensive, data-driven method of cardiovascular multimorbidity presented here represents a novel method for characterizing multimorbidity in IHD with great potential. Improved patient subgrouping may be critical guide future clinical decision making in cases, where it is non-trivial how to manage the angiographic findings or to find the optimal combination of drugs for a given patient.

## Data Availability

The code and data used in this study to generate the results are available upon reasonable request to the authors.

## Non-standard abbreviations

CAG: Coronary arteriography
CCTA: Coronary computed tomography angiography
ICD-10: International Statistical Classification of Diseases and Related Health Problems 10th Revision
IHD: Ischemic heart disease
MCL: Markov clustering
NPR: Danish National Patient Registry
O/E-ratio: Observed-expected-ratio
PRS: Polygenic risk score

## Acknowledgement

The authors would like to thank (1) research programmer, Troels Siggaard, Novo Nordisk Foundation Center for Research, University of Copenhagen, Denmark for continuous and reliable infrastructure support, and (2) Head of Cardiovascular Research, Hilma Hólm, deCODE genetics, Iceland for insightful comments

## Ethics approvals and data access and

The study was approved by The National Ethics Committee (1708829, ‘Genetics of CVD’—a genome-wide association study on repository samples from Copenhagen Hospital Biobank), The Danish Data Protection Agency (ref: 514-0255/18-3000, 514-0254/18-3000, SUND-2016-50), The Danish Health Data Authority (ref: FSEID-00003724 and FSEID-00003092), and The Danish Patient Safety Authority (3-3013-1731/1/). Danish personal identification numbers were pseudonymized prior to any analysis. Study design, methods and results were reported in agreement with the STROBE statement(46).

Application for registry data access can be made to the Danish Health Data Authority (contact). Anyone wishing access to the data and use them for research will be required to meet research credentialing requirements as outlined at the authority’s web site: <u>sundhedsdatastyrelsen.dk/da/english/health_data_and_registers/research_services</u>. Requests are normally processed within three to six months.

## Code availability statement

The code used to generate the results including the clustering pipeline will be made publicly available upon publication.

## Supplemental material

S1 Fig: Classification of new ischemic events.

S1 Table: Eligible codes for inclusion and outcomes

S1 Appendix: Construction of patient similarity network, MCL algorithm settings and assessment of cluster robustness

- S2 Fig: Selection of number of components.
- S3 Fig: Limiting edge-density and average node degree in sex-specific similarity networks.

S2 Appendix: Preprocessing of laboratory data

- S2 Table: Laboratory codes included in assessment of data quality and completeness S3 Appendix: Calculation of polygenetic risk scores for 14 traits

S4 Fig: Results of robustness analysis.

S3 Table: Comparison of mean age at index in 31 cluster using Tukey’s HSD

S4 Table: Demographics for patients not cluster or were in clusters of size < 500

S5A-B Table: Cluster-wise summarized O/E-ratios, 10 largest O/E-ratios and 10 lowest O/E-ratios.

S6 Table: Chi-squared test for distribution laboratory values in clusters S7 Table: Traits with significantly different PGS distributions in clusters

