## Supplemental Material for "Subgrouping multimorbid patients with ischemic heart disease by means of unsupervised clustering: A cohort study of 72,249 patients defined comprehensively by diagnoses prior to presentation"

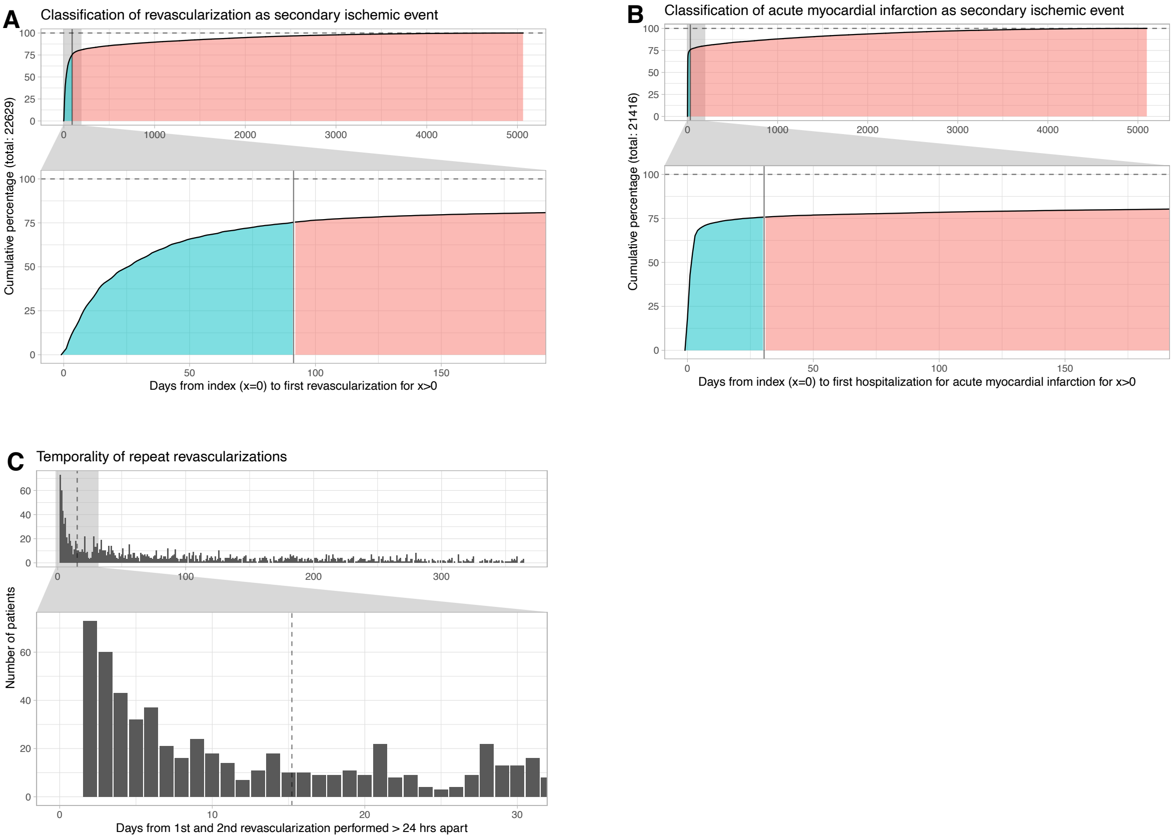


**S1 Fig: Classification of new ischemic events.** A: Time from index to first revascularization vs. percentage of patients revascularized. Blue corresponds to events related to establishment of IHD. Red corresponds to events considered new ischemic events. B: Time from index to first hospitalization for acute myocardial infarction vs. percentage of patients hospitalized. Blue corresponds to events related to index. Red corresponds to events considered new ischemic events. C: Distribution of days between revascularization for patients subjected to >1 performed >24 hours apart. Revascularizations performed <2 weeks apart were analyzed as a single event performed at date of the earliest revascularization. Marked by dashed line. IHD: Ischemic heart disease.

### S1 Table: Eligible codes for inclusion and outcomes

| **ICD-10[1] chapter IX** | | | **Definition, level 3** |
|---|---|---|---|
| **Block** | **Level 3** | **Level 4** | |
| **R94** | I20 | I20.0*, I20.1, I20.8, I20.9 | Angina pectoris |
| | I21* | I21.0, I21.1, I21.2, I21.3, I21.4, I21.9 | Acute myocardial infarction |
| | I23 | I25.0, I25.1, I25.2, I25.23, I25.4, I25.5, I25.6, I25.8, I25.9 | Certain current complications following acute myocardial infarction |
| | I24 | I24.0, I24.1, I24.8, I24.9 | Certain current complications following acute myocardial infarction |
| | I25 | I25.0, I25.1, I25.2, I25.23, I25.4, I25.5, I25.6, I25.8, I25.9 | Chronic ischemic heart disease |

| **Nomesco[2] code** | **Procedure** |
|---|---|
| FNA* | Connection to coronary artery from internal mammary artery |
| FNB* | Connection to coronary artery from gastroepiploic artery |
| FNC* | Aorto-coronary venous bypass |
| FND* | Aorto-coronary bypass using prosthetic graft |
| FNE* | Coronary bypass using free arterial graft |
| FNF* | Coronary thrombendarterectomy |
| FNG* | Expansion and recanalisation of coronary artery |

| **SKS[3] code** | **Procedure** |
|---|---|
| UXAC85[A-D] | Coronary arteriography |
| UXCC00A | Coronary computed tomography angiography |

| **SHAK[4] code** | **Hospital** |
|---|---|
| 1301 | Rigshospitalet |
| 1309 | Bispebjerg og Frederiksberg Hospitaler |
| 1330 | Amager og Hvidovre Hospital |
| 1351 | Amager Hospital |
| 1401 | Frederiksberg Hospital |
| 1501 | Gentofte Hospital |
| 1502 | Glostrup Hospital |
| 1516 | Herlev og Gentofte Hospital |
| 2000 | Hospitalerne i Nordsjælland |
| 2501 | Amtssygehuset i Roskilde |
| 3800 | Region Sjællands Sygehusvæsen |
| 4001 | Bornholms Hospital |

[1] ICD-10 = WHO International classification of diseases and health related problems 10th edition. Danish version where code types A, B and G included in our definition of primary and secondary codes.
[2] NOMESCO = Nordic Medico-Statistical Committee
[3] SKS = Sundhedsstyrelses klassifikationsstystem [Danish]
[4] SHAK = Sygehus- og afdelingsklassifikation [Danish]
[*] Included in the composite outcome new ischemic events. For ICD-10 codes only code types A (primary) and in-hospital patients.

**S1 Appendix: Construction of patient similarity network, MCL algorithm settings and assessment of cluster robustness**

To define the patient similarity network, a lower rank approximation of the *n* x *m* matrix was created using the "truncatedSVD" implecpmentation of SVD from the python package scikit-learn with 41 components, 10 iterations, and fixed random seed of 42 to ensure reproducible results. Thus, the 3,046 diagnoses were represented in 41 components based on this lower rank approximation. By selecting 41 components, the accumulated explained variance ratio was 0.50 (S2 Fig). To reduce the density of the patient similarity network, while still retaining an informative topology, all edges with an edge weight less than 0.3 were removed and the number of edges connected to each node were limited using the "#ceilnb" transformation from "mcl-edge"; a maximum of 8000 was used (S3 Fig). The weights of the remaining edges in the network were shifted such that the lowest weight was 0.0 as recommended in the MCL manual. The final pre-processed networks were then used as input for the "mcl" implementation of the MCL algorithm (1). We used a pruning scheme of -P 7000 -S 800 -R 900 -pct 90, and a pre-inflation factor of 0.5 to make edge-weights more homogenous. For the MCL clustering, we selected a pre-inflation parameter of 2.0 corresponding to the default in the MCL manual (2).

For cluster robustness assessment, diluted versions of the reference clustering were generated by deleting edges with a probability of α, where α would range between 0 and 50% (3). An α of 0 would leave the network unchanged. In contrast, the shuffled versions of the network had the same number of nodes and edges as the reference clustering. The shuffled networks were generated as described by Karrer et al. (4). Finally, the generated clusters were compared to the reference clustering and the variances were quantified with reference to the so-called variation of information measure (VI) (5).

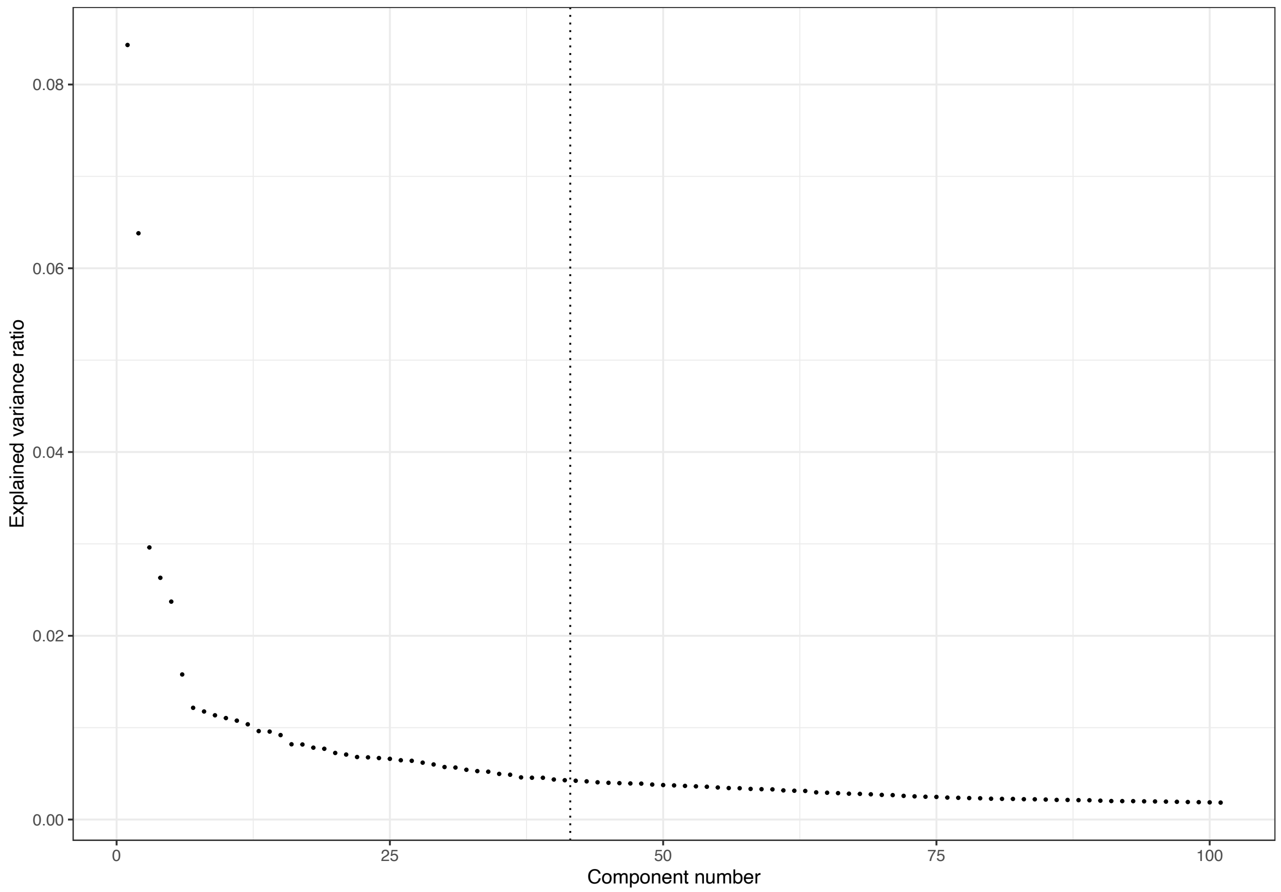


**S2 Fig: Selection of number of components.** X-axis: Component number ranked by explained variance ratio. Y-axis: Explained variance ratio. Dashed horizontal line indicates the cutoff.

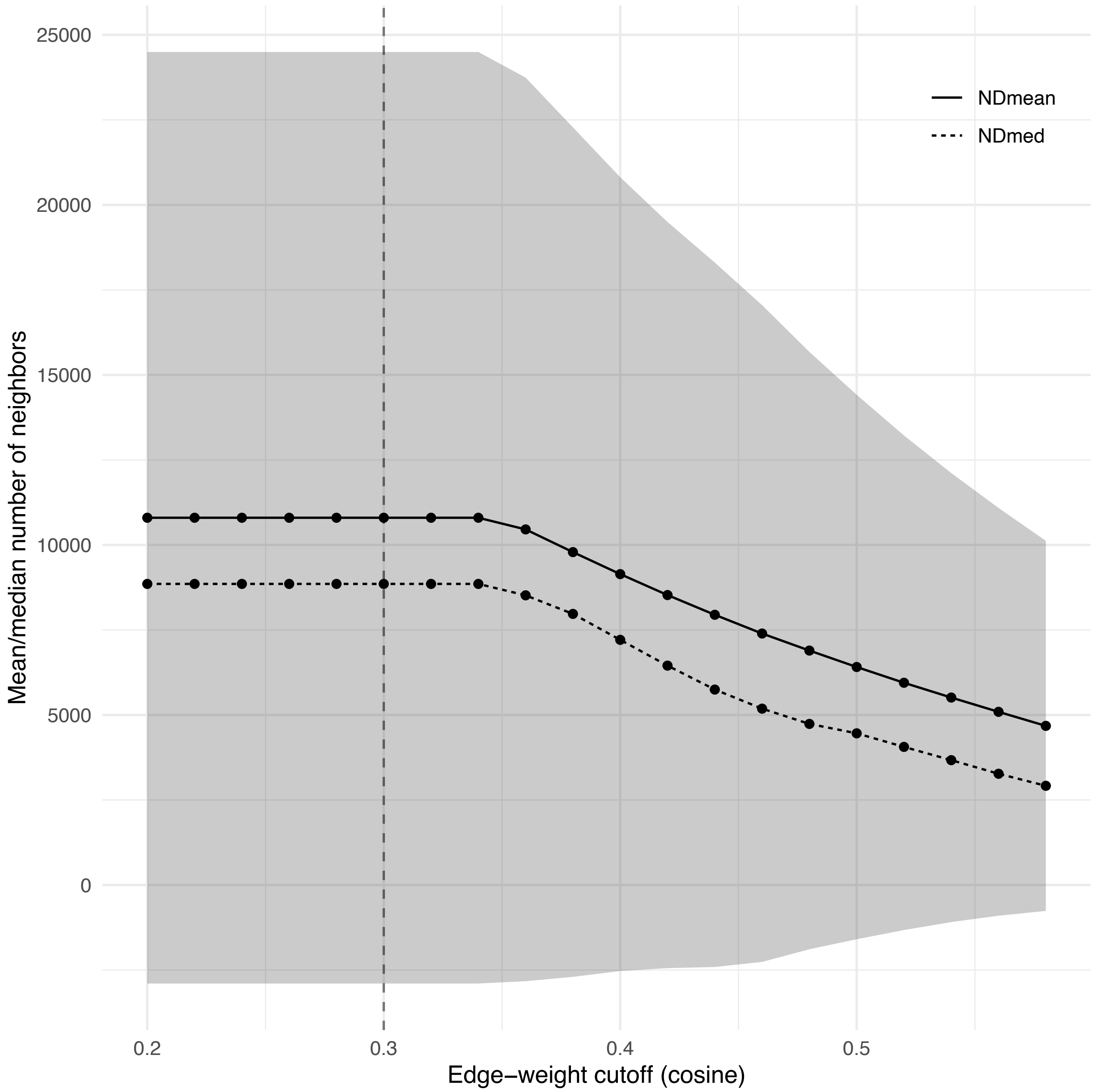


**S3 Fig: Limiting edge-density and average node degree in sex-specific similarity networks**. Mean/median number of neighbors against edge-weight cutoff in the patient similarity network. Only edges with a weight higher than 0.3 (as indicated by the vertical, dashed bar) were retained.

### S2 Appendix: Preprocessing of laboratory data

The laboratory test results from the EHR data were originally archived in the administrative biochemical databases Labka and BCC (6). In relation to the EHR data used in this study, Labka covers the hospitals with SHAK codes 1301, 1309, 1330, 1351, 1401, 1501, 1516 and 4001 in the Capital Region of Denmark and BCC covers the hospitals with SHAK codes 2000 and 2501 in Region Zealand for the periods 2009-2016 and 2012-2016, respectively (S2 Table). Biochemical laboratory tests were either classified in accordance with the Nomenclature, Properties and Units (NPU) or local systems (7). Reference intervals were provided from the laboratories that analyzed the blood tests. Biochemical data was expected to be available for the patients where the index procedure was performed at a hospital located in either the Capital Region or Region Zealand at a time that was covered by the two databases.

**S2 Table: Laboratory codes included in assessment of data quality and completeness**

| Blood analyte | NPU codes and local systems |
|---|---|
| Sodium | NPU03429, GEN00992, NPU03796, POC00022, 240, POC00021, POC00023, GEN00990 |
| Potassium | NPU03230, GEN00995, POC00019, POC00018, POC00020, GEN00993 |
| Hemoglobin | NPU02319, GEN00989, NPU02321, NPU02320, NPU02322, NPU17007, POC00013, NPU04208, NPU01393, POC00012, POC00014, NPU29057, GEN00987 |
| Creatinine / EGFR | NPU04998, NPU03918, NPU09102, NPU19661, NPU14048, NPU03800, HLL00037, DNK35131, POC00109, RHB00941, NPU28842 |

A total of 48,957 patients (30,736 males and 18,221 females) were included from a hospital where biochemical data was available (67.8% of the entire cohort). As an indicator for data completeness and quality, the number of patients where available biochemical data at time of index agreed with the clinical standard of care was assessed. This implied that patients had sodium, potassium, hemoglobin, and creatinine (or estimated glomerular filtration rate) measured maximum 90 days before or at the day of index. The 31,224 patients who fulfilled this requirement were included in the biochemical analysis. Laboratory measurements available for at least 50% of these patients were included in the analysis. In cases where patients had more than one test available in the period up from 90 days before to index, the test closest to index was used. As listed in the main text included samples were plasma levels of potassium, sodium, hemoglobin, estimated glomerular filtration (eGFR), creatinine, carbamide, glucose, troponin (I/T), HDL cholesterol, LDL cholesterol, total cholesterol, leukocytes, C-reactive protein, lymphocytes, monocytes, neutrophils, basophils, platelets, INR, alanine transaminase, albumin, alkaline phosphatase, bilirubin, and triglyceride. All analyses of biochemical data were performed in R 3.6.2 using the “ComplexHeatmap” and “circlize” packages .

### S3 Appendix: Calculation of polygenetic risk scores for 14 traits

Polygenic risk scores were calculated using the LDpred2 framework, implemented in the R package bigsnpr (v1.11.6) with R version 4.0.0 and the workflow management system Snakemake (11–13). In preparation for PGS calculations, autosomal genotype data from 242,644 individuals in the Copenhagen Hospital Biobank – Cardiovascular Disease Cohort (CHB-CVDC) (14) was filtered to only include variants present in LDpred2's recommended set of 1,054,330 reference variants. This recommended set is based on the reference set HapMap3 from the International HapMap project, which was established by genotyping 1.6 million single nucleotide polymorphisms (SNPs) in 1,184 individuals from 11 global populations (15). Any missing genotype information was assumed to be the affected locus' reference allele.

We matched the remaining set of 994,643 genotyped variants with variants found in summary statistics data corresponding to 14 traits, obtained from nine GWAS meta-analyses (atrial fibrillation (16), BMI-adjusted type 2 diabetes (17), chronic kidney disease (18), HDL cholesterol levels (19), heart failure (20), LDL cholesterol levels (19), stroke (21), total cholesterol levels (19), triglyceride levels (19)) and five GWAS (acute myocardial infarction (22), coronary artery disease (23), diastolic blood pressure (24), non-alcoholic fatty liver disease (25), systolic blood pressure (24)). Variants present in both genotype and summary statistics data were then subject to LDpred2's recommended standard deviation quality control. After variant matching and quality control, a mean of 963,354 (S.D. 87,774) variants remained for subsequent per-chromosome risk score calculation for each of the 14 traits. We used the LDpred2-auto algorithm with 30 Gibbs sampling chains, 1,000 burn-in iterations and 500 iterations after burn-in. The initial values for the 30 sampling chains were a) the LDSC regression estimate for heritability $h^2$ (same for all chains); b) one of 30 initial values for the proportion of causal variants *p*, evenly spaced on a logarithmic scale from $10^{-4}$ to 0.5.

Variant effect sizes were calculated from each set of 30 sampling chains (per trait and chromosome) through a three-step process, which serves to ensure that the model (spanning 30 chains) successfully converged: 1) computing the standard deviations of each chains' predicted scores, 2) keeping only the chains within three median absolute deviations from the median standard deviation, 3) averaging the effect sizes of the remaining chains. Across the 308 per-chromosome models (14 traits times 22 chromosomes), 28.9 chains were included in the final score on average. For each individual, we calculated per-chromosome risk scores by multiplying the average variant effect sizes with the individual's corresponding genotype, and then added the per-chromosome risk scores up into one genome-wide PGS. To ease comparisons across traits, each trait's PGS distribution was scaled to a mean of zero and a standard deviation of one.

### References, S1-3 Appendices

1. Van Dongen SM. Graph clustering by flow simulation. PhD thesis, University of Utrecht, May 2000.

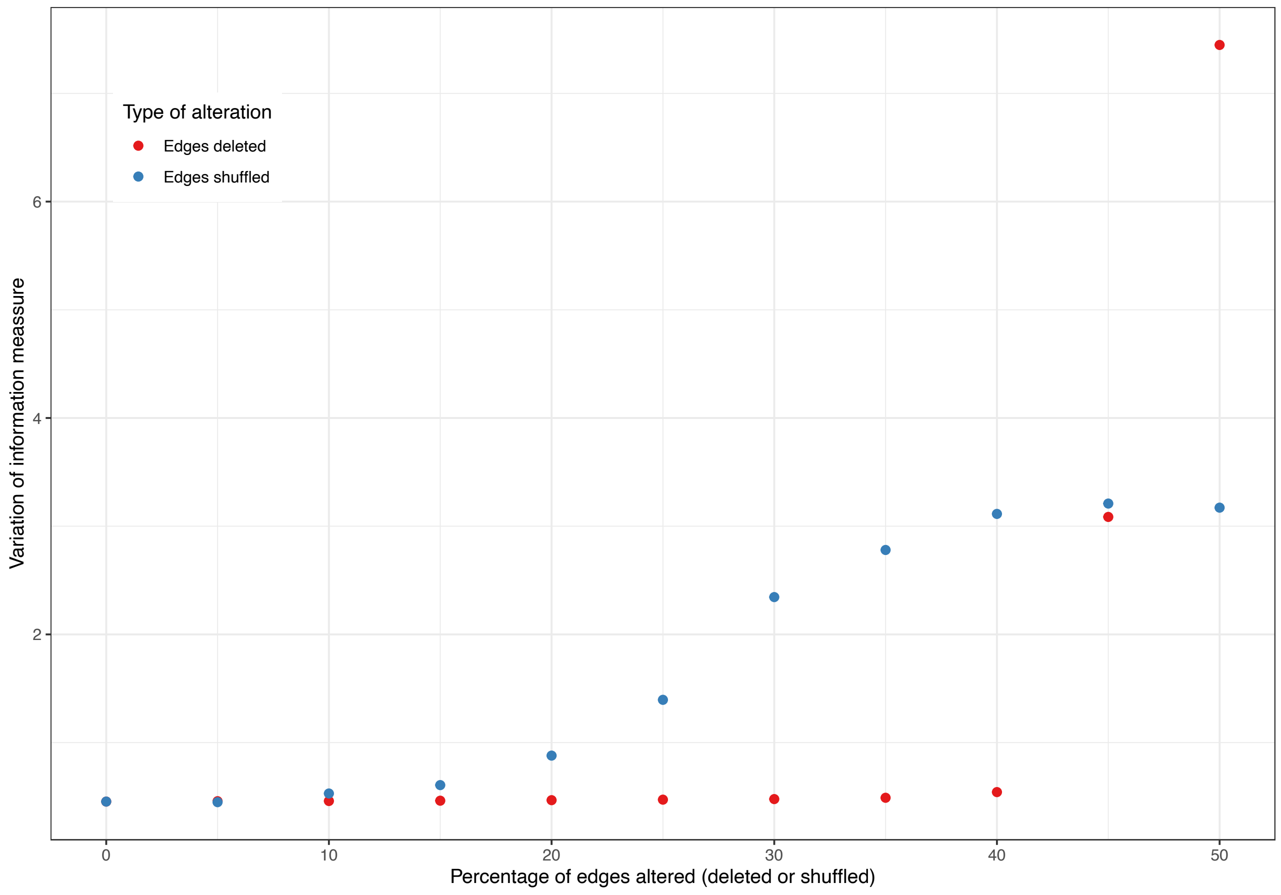


**S4 Fig: Results of robustness analysis.** X-axis: Percentage of altered edges (deleted or removed). Y-axis: Variation of information measure compared to the reference graph. Legend: Type of alteration, with 10 mutations of the reference graph for each type.

**S3 Table: Comparison of mean age at index in 31 cluster using Tukey's HSD**

See file *S3Table.pdf*

**S4 Table: Demographics for patients that did not cluster or were in clusters of size < 500**

| **Cohort demographics** | **Total** | **Males** | **Females** |
|---|---|---|---|
| Number of patients | 5,113 | 3,878 | 1,235 |
| Mean age at index (SD) | 60.7 | 60.0 | 63.0 |
| **Outcomes, number of cases** | **Total** | **Males** | **Females** |
| New ischemic events | 995 | 780 | 175 |
| Death from non-IHD causes | 352 | 274 | 78 |
| Censored | 3,624 | 2,707 | 917 |
| **Outcomes, time to event** | **Mean time to event in years (SD)** | | |
| | **Total** | **Males** | **Females** |
| New ischemic events | 1.55 (1.41) | 1.59 (1.43) | 1.39 (1.32) |
| Death from non-IHD causes | 2.25 (1.50) | 2.18 (1.47) | 2.5 (1.49) |
| Censored | 4.54 (0.95) | 4.52 (0.96) | 2.47 (1.49) |
| Total | 4.02 (1.52) | 3.98 (1.54) | 4.17 (1.44) |

**S5a Table: Degree of enrichment (sum) and top-10 O/E-ratios per cluster**

**S5b Table: Bottom-10 O/E-ratios < 1 per cluster**

See file *S5Table.pdf*

**S6 Table: Chi-squared test for distribution laboratory values in clusters**

| Component | P-val. | Adj. P-val. |
|---|---|---|
| Alanine transaminase (ALAT) | 4.78 e-22 | 1.15e-20 |
| Albumin | 4.81e-22 | 1.15e-20 |
| Alkaline phosphatase | 2.01e-22 | 4.82e-21 |
| Bilirubin | 1.09e-13 | 2.60e-12 |
| C-reactive protein (CRP) | 1.65e-96 | 3.95e-95 |
| Carbamide | 5.49-e200 | 1.32e-198 |
| Cholesterol HDL | 1.99e-66 | 4.77e-65 |
| Cholesterol LDL | 4.86e-53 | 1.17e-51 |
| Cholesterol total | 2.64e-58 | 6.34 e-57 |
| Coagulation factor II + VII + X | 7.96e-280 | 1.91e-278 |
| Creatinine | 9.28e-302 | 2.23e-300 |
| Eosinophils | 4.43e-6 | 1.06e-4 |
| Estimated glomerular filtration rate (eGFR) | 0 | 0 |
| Glucose | 0 | 0 |
| Hemoglobin | 2.77e-218 | 6.65e-217 |
| Leukocytes | 1.42e-39 | 3.41e-38 |
| Lymphocytes | 1.54e-17 | 3.69e-16 |
| Monocytes | 1.06e-11 | 2.55e-10 |
| Neutrophils | 5.69e-20 | 1.36e-18 |
| Platelets | 2.39e-23 | 5.73e-22 |
| Potassium | 9.03e-32 | 2.17e-30 |
| Sodium | 2.24e-74 | 5.38e-74 |
| Triglyceride | 2.10e-60 | 5.04 e-59 |
| Troponin | 7.10e-73 | 1.70e-71 |

**S7 Table:** Traits with significantly different PGS distributions in clusters

| Cluster | n | trait | effect | effect size | FDR |
|---|---|---|---|---|---|
| C1 | 2,025 | Systolic Blood Pressure | + | 0.20 | <0.0005 |
| | | Diastolic Blood Pressure | + | 0.16 | <0.0005 |
| | | Total Cholesterol | - | -0.08 | 0.026 |
| C4 | 1,532 | Atrial Fibrillation | + | 0.57 | <0.0005 |
| | | Heart Failure | + | 0.08 | 0.031 |
| | | Coronary Artery Disease | - | -0.12 | 0.001 |
| | | T2D (BMI-adj.) | - | -0.11 | 0.001 |
| | | Acute Myocardial Infarction | - | -0.08 | 0.031 |
| | | Triglyceride | - | -0.08 | 0.044 |
| | | Total Cholesterol | - | -0.08 | 0.046 |
| C5 | 1,136 | T2D (BMI-adj.) | + | 0.55 | <0.0005 |
| | | NAFLD | + | 0.11 | 0.021 |
| C6 | 860 | Total Cholesterol | + | 0.21 | <0.0005 |
| | | Triglyceride | + | 0.20 | <0.0005 |
| | | LDL Cholesterol | + | 0.15 | 0.001 |
| | | Coronary Artery Disease | + | 0.15 | 0.001 |
| | | Diastolic Blood Pressure | - | -0.13 | 0.015 |
| | | Systolic Blood Pressure | - | -0.11 | 0.040 |
| C8 | 817 | Systolic Blood Pressure | - | -0.16 | 0.001 |
| | | Stroke | - | -0.12 | 0.023 |
| | | Coronary Artery Disease | - | -0.12 | 0.028 |
| | | Diastolic Blood Pressure | - | -0.11 | 0.031 |
| | | LDL Cholesterol | - | -0.10 | 0.047 |
| C10 | 744 | Coronary Artery Disease | - | -0.13 | 0.017 |
| | | Acute Myocardial Infarction | - | -0.13 | 0.021 |
| C11 | 718 | Stroke | - | -0.11 | 0.040 |
| | | Heart Failure | - | -0.11 | 0.040 |
| C12 | 649 | Coronary Artery Disease | - | -0.14 | 0.013 |
| | | Acute Myocardial Infarction | - | -0.12 | 0.033 |
| C13 | 606 | Diastolic Blood Pressure | - | -0.12 | 0.040 |
| C15 | 588 | LDL Cholesterol | + | 0.14 | 0.020 |
| | | Total Cholesterol | + | 0.14 | 0.026 |
| C17 | 348 | Systolic Blood Pressure | + | 0.16 | 0.040 |
| C18 | 481 | T2D (BMI-adj.) | + | 0.15 | 0.023 |
| | | Acute Myocardial Infarction | + | 0.15 | 0.028 |
| | | Atrial Fibrillation | - | -0.13 | 0.049 |
| C23 | 290 | T2D (BMI-adj.) | + | 0.27 | 0.001 |
| C25 | 297 | Coronary Artery Disease | + | 0.24 | 0.002 |
| | | Acute Myocardial Infarction | + | 0.18 | 0.032 |
| | | Heart Failure | + | 0.18 | 0.031 |
| C27 | 231 | Stroke | + | 0.24 | 0.015 |

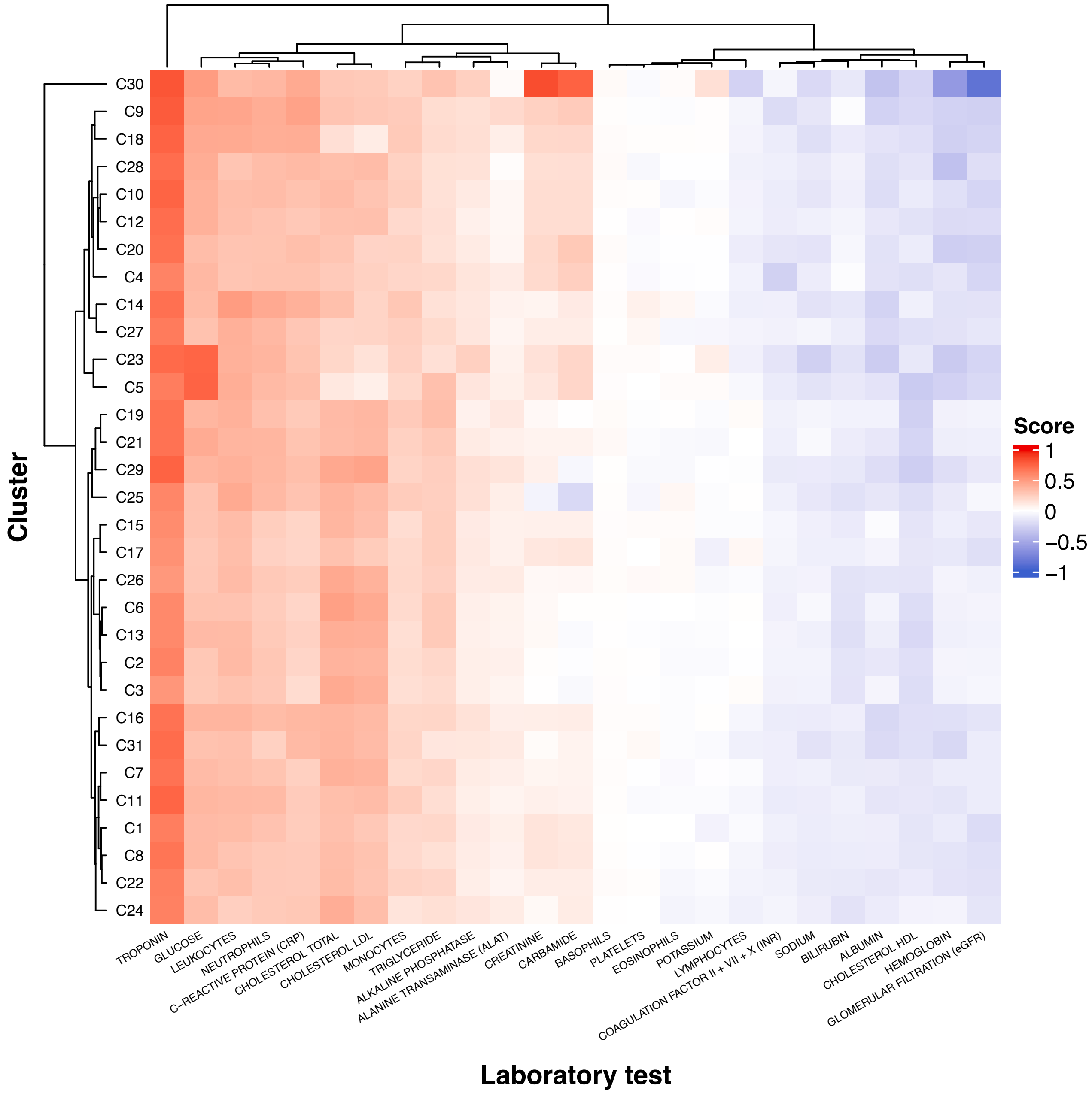


**S5 Fig: Heatmap of clusters based on laboratory profiles.** Summary of results from the phenotypic characterization of clusters based on laboratory data. *Score* refers to the mean summarized values per cluster, where values were assigned based on the results of the laboratory test per patient. Values of -1, 0, and 1, indicates below, within or above reference range, respectively. For details, see Methods. X-axis: Laboratory test. Y-axis: Cluster.
