## Supplementary material for "Subgrouping multimorbid patients with ischemic heart disease by means of unsupervised clustering: A cohort study of 72,249 patients defined comprehensively by diagnoses prior to presentation": S3 Table

### S3 Table: Comparison of mean age at index in 31 cluster using Tukey's HSD

| contrast | null.value | estimate | conf.low | conf.high | adj.p.value |
|---|---|---|---|---|---|
| C2-C1 | 0 | -6.18 | -6.89 | -5.47 | 0 |
| C3-C1 | 0 | -7.96 | -8.73 | -7.20 | 0 |
| C4-C1 | 0 | 0,221527778 | 04.02 | 05.57 | 0 |
| C5-C1 | 0 | -0.920 | -1.70 | -0.139 | 3.60e- 3 |
| C6-C1 | 0 | -5.12 | -5.95 | -4.29 | 0 |
| C7-C1 | 0 | -0.992 | -1.84 | -0.141 | 4.40e- 3 |
| C8-C1 | 0 | 06.29 | 05.39 | 07.20 | 0 |
| C9-C1 | 0 | -0.0638 | -0.993 | 0,600694444 | 1 e+ 0 |
| C10-C1 | 0 | 09.43 | 08.50 | 10.04 | 0 |
| C11-C1 | 0 | 01.32 | 0,240972222 | 02.29 | 1.42e- 4 |
| C12-C1 | 0 | 05.48 | 04.50 | 06.46 | 0 |
| C13-C1 | 0 | -6.13 | -7.14 | -5.12 | 0 |
| C14-C1 | 0 | 03.47 | 02.45 | 04.48 | 0 |
| C15-C1 | 0 | -0.908 | -1.93 | 0,078472222 | 1.79e- 1 |
| C16-C1 | 0 | -0.761 | -1.86 | 0,238194444 | 7.29e- 1 |
| C17-C1 | 0 | 0,355555556 | -0.715 | 0,093055556 | 1.00e+ 0 |
| C18-C1 | 0 | 03.39 | 02.15 | 0,210416667 | 3.19e-13 |
| C19-C1 | 0 | -6.27 | -7.55 | -4.99 | 0 |
| C20-C1 | 0 | 0,301388889 | 05.44 | 08.04 | 0 |
| C21-C1 | 0 | -3.78 | -5.15 | -2.41 | 3.24e-13 |
| C22-C1 | 0 | 04.38 | 03.01 | 0,261111111 | 0 |
| C23-C1 | 0 | -6.09 | -7.50 | -4.68 | 0 |
| C24-C1 | 0 | 03.10 | 0,089583333 | 04.51 | 3.94e-13 |
| C25-C1 | 0 | -8.56 | -10.0 | -7.10 | 0 |
| C26-C1 | 0 | -5.98 | -7.45 | -4.52 | 0 |
| C27-C1 | 0 | 0,239583333 | -1.14 | 0,099305556 | 1.00e+ 0 |
| C28-C1 | 0 | 0,313194444 | 05.29 | 08.53 | 0 |
| C29-C1 | 0 | -7.55 | -9.34 | -5.76 | 0 |
| C30-C1 | 0 | -3.55 | -5.37 | -1.73 | 9.14e-11 |
| C31-C1 | 0 | -0.335 | -2.17 | 01.50 | 1.00e+ 0 |
| C3-C2 | 0 | -1.78 | -2.58 | -0.993 | 3.02e-13 |
| C4-C2 | 0 | 11.00 | 10.02 | 11.08 | 0 |
| C5-C2 | 0 | 05.26 | 04.45 | 06.07 | 0 |
| C6-C2 | 0 | 01.06 | 0,140972222 | 0,104861111 | 1.32e- 3 |
| C7-C2 | 0 | 05.19 | 04.31 | 06.06 | 0 |
| C8-C2 | 0 | 12.05 | 11.05 | 13.04 | 0 |
| C9-C2 | 0 | 06.11 | 05.16 | 07.07 | 0 |
| C10-C2 | 0 | 15.06 | 14.07 | 16.06 | 0 |
| C11-C2 | 0 | 07.50 | 06.50 | 08.49 | 0 |
| C12-C2 | 0 | 11.07 | 10.07 | 12.07 | 0 |
| C13-C2 | 0 | 0,356944444 | -0.981 | 01.08 | 1 e+ 0 |
| C14-C2 | 0 | 0,420138889 | 0,375694444 | 10.07 | 0 |
| C15-C2 | 0 | 05.27 | 04.23 | 06.31 | 0 |
| C16-C2 | 0 | 05.42 | 04.29 | 06.54 | 0 |
| C17-C2 | 0 | 0,297916667 | 05.44 | 0,356944444 | 0 |
| C18-C2 | 0 | 09.57 | 08.31 | 10.08 | 0 |
| C19-C2 | 0 | -0.0893 | -1.38 | 01.21 | 1 e+ 0 |
| C20-C2 | 0 | 12.09 | 11.06 | 14.02 | 0 |
| C21-C2 | 0 | 02.40 | 01.02 | 0,179166667 | 3.07e- 8 |
| C22-C2 | 0 | 10.06 | 09.17 | 12.00 | 0 |

| contrast | null.value | estimate | conf.low | conf.high | adj.p.value |
|---|---|---|---|---|---|
| C23-C2 | 0 | 0,607638889 | -1.34 | 01.51 | 1 e+ 0 |
| C24-C2 | 0 | 09.28 | 0,351388889 | 10.07 | 0 |
| C25-C2 | 0 | -2.38 | -3.86 | -0.908 | 5.53e- 7 |
| C26-C2 | 0 | 0,135416667 | -1.29 | 0,088888889 | 1 e+ 0 |
| C27-C2 | 0 | 06.52 | 05.02 | 08.03 | 0 |
| C28-C2 | 0 | 13.01 | 11.05 | 14.07 | 0 |
| C29-C2 | 0 | -1.38 | -3.18 | 0,297222222 | 5.03e- 1 |
| C30-C2 | 0 | 0,127083333 | 0,555555556 | 04.46 | 2.82e- 5 |
| C31-C2 | 0 | 0,266666667 | 0,19375 | 0,339583333 | 0 |
| C4-C3 | 0 | 12.08 | 11.09 | 13.06 | 0 |
| C5-C3 | 0 | 07.04 | 06.19 | 0,354166667 | 0 |
| C6-C3 | 0 | 0,141666667 | 0,106944444 | 0,176388889 | 0 |
| C7-C3 | 0 | 0,317361111 | 06.05 | 0,353472222 | 0 |
| C8-C3 | 0 | 14.03 | 13.03 | 15.02 | 0 |
| C9-C3 | 0 | 0,354166667 | 0,313194444 | 0,395138889 | 0 |
| C10-C3 | 0 | 17.04 | 16.04 | 18.04 | 0 |
| C11-C3 | 0 | 09.28 | 08.25 | 10.03 | 0 |
| C12-C3 | 0 | 13.04 | 12.04 | 14.05 | 0 |
| C13-C3 | 0 | 0,1 | 0,531944444 | 0,146527778 | 4.82e- 8 |
| C14-C3 | 0 | 11.04 | 10.04 | 12.05 | 0 |
| C15-C3 | 0 | 07.06 | 0,276388889 | 08.14 | 0 |
| C16-C3 | 0 | 07.20 | 06.04 | 08.36 | 0 |
| C17-C3 | 0 | 08.48 | 07.20 | 0,427083333 | 0 |
| C18-C3 | 0 | 11.04 | 10.01 | 12.06 | 0 |
| C19-C3 | 0 | 0,090277778 | 0,256944444 | 03.02 | 6.19e- 4 |
| C20-C3 | 0 | 14.07 | 13.04 | 16.01 | 0 |
| C21-C3 | 0 | 04.18 | 0,136805556 | 05.59 | 9.64e-14 |
| C22-C3 | 0 | 12.03 | 10.09 | 13.08 | 0 |
| C23-C3 | 0 | 0,102083333 | 0,292361111 | 03.32 | 5.07e- 4 |
| C24-C3 | 0 | 11.01 | 0,417361111 | 12.05 | 0 |
| C25-C3 | 0 | -0.599 | -2.10 | 0,627777778 | 1.00e+ 0 |
| C26-C3 | 0 | 0,109722222 | 0,327083333 | 03.49 | 3.37e- 4 |
| C27-C3 | 0 | 08.31 | 0,304166667 | 0,433333333 | 0 |
| C28-C3 | 0 | 14.09 | 13.02 | 16.05 | 0 |
| C29-C3 | 0 | 0,284722222 | -1.42 | 02.24 | 1.00e+ 0 |
| C30-C3 | 0 | 04.41 | 02.56 | 06.27 | 2.13e-13 |
| C31-C3 | 0 | 0,335416667 | 0,261111111 | 09.50 | 0 |
| C5-C4 | 0 | -5.71 | -6.58 | -4.84 | 0 |
| C6-C4 | 0 | -9.91 | -10.8 | -9.00 | 0 |
| C7-C4 | 0 | -5.78 | -6.71 | -4.85 | 0 |
| C8-C4 | 0 | 01.50 | 0,363888889 | 02.48 | 3.38e- 6 |
| C9-C4 | 0 | -4.85 | -5.86 | -3.85 | 0 |
| C10-C4 | 0 | 0,211111111 | 0,169444444 | 0,253472222 | 0 |
| C11-C4 | 0 | -3.47 | -4.52 | -2.43 | 0 |
| C12-C4 | 0 | 0,478472222 | -0.366 | 0,093055556 | 8.21e- 1 |
| C13-C4 | 0 | -10.9 | -12.0 | -9.84 | 0 |
| C14-C4 | 0 | -1.32 | -2.41 | -0.239 | 1.76e- 3 |
| C15-C4 | 0 | -5.70 | -6.79 | -4.61 | 0 |
| C16-C4 | 0 | -5.55 | -6.72 | -4.38 | 0 |
| C17-C4 | 0 | -4.28 | -5.56 | -2.99 | 0 |
| C18-C4 | 0 | -1.40 | -2.70 | -0.102 | 1.70e- 2 |
| C19-C4 | 0 | -11.1 | -12.4 | -9.73 | 0 |

| contrast | null.value | estimate | conf.low | conf.high | adj.p.value |
|---|---|---|---|---|---|
| C20-C4 | 0 | 0,107638889 | 0,411111111 | 03.30 | 2.86e- 5 |
| C21-C4 | 0 | -8.57 | -9.99 | -7.15 | 0 |
| C22-C4 | 0 | -0.409 | -1.83 | 01.02 | 1.00e+ 0 |
| C23-C4 | 0 | -10.9 | -12.3 | -9.42 | 0 |
| C24-C4 | 0 | -1.69 | -3.15 | -0.228 | 5.18e- 3 |
| C25-C4 | 0 | -13.4 | -14.9 | -11.8 | 0 |
| C26-C4 | 0 | -10.8 | -12.3 | -9.26 | 0 |
| C27-C4 | 0 | -4.45 | -5.98 | -2.91 | 2.72e-13 |
| C28-C4 | 0 | 02.12 | 0,317361111 | 0,179166667 | 6.71e- 4 |
| C29-C4 | 0 | -12.3 | -14.2 | -10.5 | 0 |
| C30-C4 | 0 | -8.34 | -10.2 | -6.48 | 0 |
| C31-C4 | 0 | -5.13 | -7.00 | -3.25 | 3.35e-13 |
| C6-C5 | 0 | -4.20 | -5.12 | -3.29 | 0 |
| C7-C5 | 0 | -0.0714 | -1.01 | 0,600694444 | 1 e+ 0 |
| C8-C5 | 0 | 07.21 | 06.23 | 08.20 | 0 |
| C9-C5 | 0 | 0,594444444 | -0.152 | 0,101388889 | 2.61e- 1 |
| C10-C5 | 0 | 10.04 | 09.34 | 11.04 | 0 |
| C11-C5 | 0 | 02.24 | 01.19 | 03.29 | 8.06e-13 |
| C12-C5 | 0 | 06.40 | 05.34 | 07.46 | 0 |
| C13-C5 | 0 | -5.21 | -6.29 | -4.12 | 0 |
| C14-C5 | 0 | 04.39 | 03.30 | 05.48 | 0 |
| C15-C5 | 0 | 0,0875 | -1.08 | 01.11 | 1 e+ 0 |
| C16-C5 | 0 | 0,111111111 | -1.01 | 01.33 | 1 e+ 0 |
| C17-C5 | 0 | 01.43 | 0,099305556 | 0,133333333 | 1.04e- 2 |
| C18-C5 | 0 | 04.31 | 03.01 | 0,250694444 | 0 |
| C19-C5 | 0 | -5.35 | -6.68 | -4.01 | 0 |
| C20-C5 | 0 | 0,3375 | 06.30 | 09.02 | 0 |
| C21-C5 | 0 | -2.86 | -4.28 | -1.44 | 1.70e-11 |
| C22-C5 | 0 | 05.30 | 0,185416667 | 0,300694444 | 0 |
| C23-C5 | 0 | -5.17 | -6.63 | -3.71 | 0 |
| C24-C5 | 0 | 04.02 | 02.56 | 05.49 | 2.89e-13 |
| C25-C5 | 0 | -7.64 | -9.16 | -6.13 | 0 |
| C26-C5 | 0 | -5.06 | -6.58 | -3.55 | 0 |
| C27-C5 | 0 | 01.27 | -0.275 | 0,139583333 | 3.30e- 1 |
| C28-C5 | 0 | 0,349305556 | 06.17 | 09.49 | 0 |
| C29-C5 | 0 | -6.63 | -8.47 | -4.80 | 0 |
| C30-C5 | 0 | -2.63 | -4.49 | -0.769 | 4.58e- 5 |
| C31-C5 | 0 | 0,40625 | -1.29 | 02.47 | 1.00e+ 0 |
| C7-C6 | 0 | 04.13 | 03.15 | 05.11 | 0 |
| C8-C6 | 0 | 11.04 | 10.04 | 12.04 | 0 |
| C9-C6 | 0 | 05.06 | 04.01 | 06.10 | 0 |
| C10-C6 | 0 | 14.06 | 13.05 | 15.06 | 0 |
| C11-C6 | 0 | 06.44 | 05.36 | 07.52 | 0 |
| C12-C6 | 0 | 10.06 | 09.51 | 11.07 | 0 |
| C13-C6 | 0 | -1.01 | -2.12 | 0,077083333 | 1.59e- 1 |
| C14-C6 | 0 | 08.59 | 07.47 | 0,424305556 | 0 |
| C15-C6 | 0 | 04.21 | 03.09 | 05.34 | 0 |
| C16-C6 | 0 | 04.36 | 03.16 | 05.56 | 0 |
| C17-C6 | 0 | 0,252083333 | 04.32 | 0,315972222 | 0 |
| C18-C6 | 0 | 08.51 | 07.18 | 0,433333333 | 0 |
| C19-C6 | 0 | -1.15 | -2.51 | 0,150694444 | 2.81e- 1 |
| C20-C6 | 0 | 11.09 | 10.05 | 13.02 | 0 |

| contrast | null.value | estimate | conf.low | conf.high | adj.p.value |
|---|---|---|---|---|---|
| C21-C6 | 0 | 01.34 | -0.107 | 0,138194444 | 1.21e- 1 |
| C22-C6 | 0 | 09.50 | 08.05 | 11.00 | 0 |
| C23-C6 | 0 | -0.970 | -2.46 | 0,359027778 | 8.23e- 1 |
| C24-C6 | 0 | 08.22 | 0,301388889 | 0,424305556 | 0 |
| C25-C6 | 0 | -3.44 | -4.98 | -1.90 | 3.25e-13 |
| C26-C6 | 0 | -0.862 | -2.41 | 0,472222222 | 9.65e- 1 |
| C27-C6 | 0 | 05.47 | 0,1875 | 07.03 | 0 |
| C28-C6 | 0 | 12.00 | 10.03 | 13.07 | 0 |
| C29-C6 | 0 | -2.43 | -4.29 | -0.579 | 3.36e- 4 |
| C30-C6 | 0 | 01.57 | -0.307 | 03.45 | 2.91e- 1 |
| C31-C6 | 0 | 0,221527778 | 0,145138889 | 0,297916667 | 2.61e-13 |
| C8-C7 | 0 | 07.29 | 06.25 | 08.32 | 0 |
| C9-C7 | 0 | 0,644444444 | -0.135 | 0,110416667 | 2.10e- 1 |
| C10-C7 | 0 | 10.04 | 09.36 | 11.05 | 0 |
| C11-C7 | 0 | 02.31 | 01.21 | 03.41 | 1.70e-12 |
| C12-C7 | 0 | 06.47 | 05.36 | 07.58 | 0 |
| C13-C7 | 0 | -5.14 | -6.27 | -4.00 | 0 |
| C14-C7 | 0 | 04.46 | 03.32 | 0,25 | 0 |
| C15-C7 | 0 | 0,583333333 | -1.06 | 01.23 | 1 e+ 0 |
| C16-C7 | 0 | 0,160416667 | -0.988 | 01.45 | 1.00e+ 0 |
| C17-C7 | 0 | 01.50 | 0,119444444 | 0,141666667 | 7.96e- 3 |
| C18-C7 | 0 | 04.38 | 03.04 | 0,259027778 | 0 |
| C19-C7 | 0 | -5.28 | -6.65 | -3.90 | 0 |
| C20-C7 | 0 | 0,342361111 | 06.33 | 09.13 | 0 |
| C21-C7 | 0 | -2.79 | -4.25 | -1.33 | 3.09e-10 |
| C22-C7 | 0 | 05.37 | 0,188194444 | 0,308333333 | 0 |
| C23-C7 | 0 | -5.10 | -6.60 | -3.60 | 0 |
| C24-C7 | 0 | 04.10 | 02.59 | 0,25 | 3.44e-13 |
| C25-C7 | 0 | -7.57 | -9.12 | -6.02 | 0 |
| C26-C7 | 0 | -4.99 | -6.55 | -3.44 | 0 |
| C27-C7 | 0 | 01.34 | -0.240 | 0,146527778 | 2.64e- 1 |
| C28-C7 | 0 | 0,354166667 | 06.20 | 0,416666667 | 0 |
| C29-C7 | 0 | -6.56 | -8.43 | -4.70 | 0 |
| C30-C7 | 0 | -2.56 | -4.45 | -0.668 | 1.53e- 4 |
| C31-C7 | 0 | 0,45625 | -1.25 | 02.57 | 1.00e+ 0 |
| C9-C8 | 0 | -6.36 | -7.46 | -5.25 | 0 |
| C10-C8 | 0 | 03.14 | 02.03 | 04.25 | 3.03e-13 |
| C11-C8 | 0 | -4.98 | -6.12 | -3.84 | 0 |
| C12-C8 | 0 | -0.813 | -1.96 | 0,234722222 | 6.78e- 1 |
| C13-C8 | 0 | -12.4 | -13.6 | -11.2 | 0 |
| C14-C8 | 0 | -2.83 | -4.00 | -1.65 | 2.02e-13 |
| C15-C8 | 0 | -7.20 | -8.38 | -6.02 | 0 |
| C16-C8 | 0 | -7.05 | -8.31 | -5.80 | 0 |
| C17-C8 | 0 | -5.78 | -7.15 | -4.42 | 0 |
| C18-C8 | 0 | -2.90 | -4.28 | -1.52 | 1.41e-12 |
| C19-C8 | 0 | -12.6 | -14.0 | -11.2 | 0 |
| C20-C8 | 0 | 0,309027778 | -0.987 | 0,102777778 | 1.00e+ 0 |
| C21-C8 | 0 | -10.1 | -11.6 | -8.58 | 0 |
| C22-C8 | 0 | -1.91 | -3.41 | -0.414 | 6.53e- 4 |
| C23-C8 | 0 | -12.4 | -13.9 | -10.9 | 0 |
| C24-C8 | 0 | -3.19 | -4.72 | -1.66 | 2.46e-12 |
| C25-C8 | 0 | -14.9 | -16.4 | -13.3 | 0 |

| contrast | null.value | estimate | conf.low | conf.high | adj.p.value |
|---|---|---|---|---|---|
| C26-C8 | 0 | -12.3 | -13.9 | -10.7 | 0 |
| C27-C8 | 0 | -5.95 | -7.55 | -4.34 | 0 |
| C28-C8 | 0 | 0,427777778 | -1.11 | 02.34 | 1.00e+ 0 |
| C29-C8 | 0 | -13.8 | -15.7 | -12.0 | 0 |
| C30-C8 | 0 | -9.84 | -11.8 | -7.93 | 0 |
| C31-C8 | 0 | -6.63 | -8.56 | -4.70 | 0 |
| C10-C9 | 0 | 09.50 | 08.37 | 10.06 | 0 |
| C11-C9 | 0 | 01.38 | 0,152777778 | 02.54 | 3.01e- 3 |
| C12-C9 | 0 | 05.54 | 04.37 | 0,3 | 0 |
| C13-C9 | 0 | -6.06 | -7.26 | -4.87 | 0 |
| C14-C9 | 0 | 03.53 | 02.33 | 0,217361111 | 1.45e-13 |
| C15-C9 | 0 | -0.844 | -2.05 | 0,25 | 6.94e- 1 |
| C16-C9 | 0 | -0.697 | -1.97 | 0,401388889 | 9.74e- 1 |
| C17-C9 | 0 | 0,4 | -0.808 | 0,108333333 | 1.00e+ 0 |
| C18-C9 | 0 | 03.46 | 02.06 | 0,225694444 | 3.13e-13 |
| C19-C9 | 0 | -6.20 | -7.63 | -4.78 | 0 |
| C20-C9 | 0 | 0,305555556 | 05.35 | 08.25 | 0 |
| C21-C9 | 0 | -3.72 | -5.22 | -2.21 | 3.27e-13 |
| C22-C9 | 0 | 04.45 | 0,147916667 | 0,275 | 1.72e-13 |
| C23-C9 | 0 | -6.03 | -7.57 | -4.48 | 0 |
| C24-C9 | 0 | 03.17 | 0,084722222 | 0,215972222 | 6.14e-12 |
| C25-C9 | 0 | -8.50 | -10.1 | -6.91 | 0 |
| C26-C9 | 0 | -5.92 | -7.52 | -4.32 | 0 |
| C27-C9 | 0 | 0,284027778 | -1.21 | 02.03 | 1.00e+ 0 |
| C28-C9 | 0 | 0,317361111 | 05.23 | 0,382638889 | 0 |
| C29-C9 | 0 | -7.49 | -9.39 | -5.59 | 0 |
| C30-C9 | 0 | -3.48 | -5.41 | -1.56 | 4.48e- 9 |
| C31-C9 | 0 | -0.271 | -2.22 | 0,088888889 | 1 e+ 0 |
| C11-C10 | 0 | -8.12 | -9.28 | -6.95 | 0 |
| C12-C10 | 0 | -3.95 | -5.13 | -2.78 | 0 |
| C13-C10 | 0 | -15.6 | -16.8 | -14.4 | 0 |
| C14-C10 | 0 | -5.97 | -7.17 | -4.76 | 0 |
| C15-C10 | 0 | -10.3 | -11.5 | -9.13 | 0 |
| C16-C10 | 0 | -10.2 | -11.5 | -8.92 | 0 |
| C17-C10 | 0 | -8.92 | -10.3 | -7.54 | 0 |
| C18-C10 | 0 | -6.04 | -7.44 | -4.64 | 0 |
| C19-C10 | 0 | -15.7 | -17.1 | -14.3 | 0 |
| C20-C10 | 0 | -2.69 | -4.15 | -1.24 | 1.25e- 9 |
| C21-C10 | 0 | -13.2 | -14.7 | -11.7 | 0 |
| C22-C10 | 0 | -5.05 | -6.57 | -3.54 | 0 |
| C23-C10 | 0 | -15.5 | -17.1 | -14.0 | 0 |
| C24-C10 | 0 | -6.33 | -7.88 | -4.78 | 0 |
| C25-C10 | 0 | -18.0 | -19.6 | -16.4 | 0 |
| C26-C10 | 0 | -15.4 | -17.0 | -13.8 | 0 |
| C27-C10 | 0 | -9.09 | -10.7 | -7.47 | 0 |
| C28-C10 | 0 | -2.52 | -4.26 | -0.783 | 2.17e- 5 |
| C29-C10 | 0 | -17.0 | -18.9 | -15.1 | 0 |
| C30-C10 | 0 | -13.0 | -14.9 | -11.1 | 0 |
| C31-C10 | 0 | -9.77 | -11.7 | -7.82 | 0 |
| C12-C11 | 0 | 04.16 | 0,15 | 05.37 | 0 |
| C13-C11 | 0 | -7.45 | -8.67 | -6.22 | 0 |
| C14-C11 | 0 | 02.15 | 0,636805556 | 03.38 | 2.38e- 8 |

| contrast | null.value | estimate | conf.low | conf.high | adj.p.value |
|---|---|---|---|---|---|
| C15-C11 | 0 | -2.23 | -3.46 | -0.989 | 5.77e- 9 |
| C16-C11 | 0 | -2.08 | -3.38 | -0.772 | 9.60e- 7 |
| C17-C11 | 0 | -0.806 | -2.22 | 0,421527778 | 9.55e- 1 |
| C18-C11 | 0 | 02.07 | 0,452083333 | 03.50 | 1.84e- 5 |
| C19-C11 | 0 | -7.59 | -9.04 | -6.13 | 0 |
| C20-C11 | 0 | 05.42 | 0,190277778 | 0,3125 | 0 |
| C21-C11 | 0 | -5.10 | -6.63 | -3.56 | 0 |
| C22-C11 | 0 | 03.06 | 01.52 | 0,208333333 | 3.32e-11 |
| C23-C11 | 0 | -7.41 | -8.98 | -5.84 | 0 |
| C24-C11 | 0 | 0,096527778 | 0,147916667 | 03.36 | 7.19e- 3 |
| C25-C11 | 0 | -9.88 | -11.5 | -8.26 | 0 |
| C26-C11 | 0 | -7.30 | -8.93 | -5.68 | 0 |
| C27-C11 | 0 | -0.973 | -2.62 | 0,466666667 | 9.33e- 1 |
| C28-C11 | 0 | 05.59 | 0,182638889 | 07.35 | 0 |
| C29-C11 | 0 | -8.87 | -10.8 | -6.95 | 0 |
| C30-C11 | 0 | -4.87 | -6.81 | -2.92 | 2.88e-13 |
| C31-C11 | 0 | -1.65 | -3.62 | 0,218055556 | 2.82e- 1 |
| C13-C12 | 0 | -11.6 | -12.8 | -10.4 | 0 |
| C14-C12 | 0 | -2.01 | -3.26 | -0.770 | 4.92e- 7 |
| C15-C12 | 0 | -6.39 | -7.63 | -5.14 | 0 |
| C16-C12 | 0 | -6.24 | -7.56 | -4.93 | 0 |
| C17-C12 | 0 | -4.97 | -6.39 | -3.55 | 0 |
| C18-C12 | 0 | -2.09 | -3.52 | -0.656 | 1.82e- 5 |
| C19-C12 | 0 | -11.7 | -13.2 | -10.3 | 0 |
| C20-C12 | 0 | 01.26 | -0.227 | 0,134722222 | 2.66e- 1 |
| C21-C12 | 0 | -9.26 | -10.8 | -7.72 | 0 |
| C22-C12 | 0 | -1.10 | -2.65 | 0,3125 | 6.69e- 1 |
| C23-C12 | 0 | -11.6 | -13.2 | -9.99 | 0 |
| C24-C12 | 0 | -2.38 | -3.96 | -0.796 | 6.89e- 6 |
| C25-C12 | 0 | -14.0 | -15.7 | -12.4 | 0 |
| C26-C12 | 0 | -11.5 | -13.1 | -9.83 | 0 |
| C27-C12 | 0 | -5.14 | -6.79 | -3.48 | 0 |
| C28-C12 | 0 | 01.43 | -0.340 | 03.20 | 3.67e- 1 |
| C29-C12 | 0 | -13.0 | -15.0 | -11.1 | 0 |
| C30-C12 | 0 | -9.03 | -11.0 | -7.08 | 0 |
| C31-C12 | 0 | -5.82 | -7.79 | -3.84 | 1.31e-13 |
| C14-C13 | 0 | 09.59 | 08.33 | 10.09 | 0 |
| C15-C13 | 0 | 05.22 | 0,190972222 | 06.49 | 0 |
| C16-C13 | 0 | 05.37 | 04.03 | 0,298611111 | 0 |
| C17-C13 | 0 | 0,294444444 | 05.20 | 08.08 | 0 |
| C18-C13 | 0 | 09.52 | 08.07 | 11.00 | 0 |
| C19-C13 | 0 | -0.141 | -1.62 | 01.34 | 1 e+ 0 |
| C20-C13 | 0 | 12.09 | 11.04 | 14.04 | 0 |
| C21-C13 | 0 | 02.35 | 0,547222222 | 0,188194444 | 6.61e- 6 |
| C22-C13 | 0 | 10.05 | 0,398611111 | 12.01 | 0 |
| C23-C13 | 0 | 0,250694444 | -1.56 | 0,085416667 | 1 e+ 0 |
| C24-C13 | 0 | 09.23 | 0,335416667 | 10.08 | 0 |
| C25-C13 | 0 | -2.44 | -4.08 | -0.793 | 1.07e- 5 |
| C26-C13 | 0 | 0,1 | -1.50 | 0,096527778 | 1 e+ 0 |
| C27-C13 | 0 | 06.47 | 0,222222222 | 08.14 | 0 |
| C28-C13 | 0 | 13.00 | 11.03 | 14.08 | 0 |
| C29-C13 | 0 | -1.43 | -3.37 | 0,357638889 | 5.91e- 1 |

| contrast | null.value | estimate | conf.low | conf.high | adj.p.value |
|---|---|---|---|---|---|
| C30-C13 | 0 | 02.58 | 0,425 | 04.54 | 3.42e- 4 |
| C31-C13 | 0 | 0,263194444 | 0,18125 | 0,345833333 | 2.55e-13 |
| C15-C14 | 0 | -4.37 | -5.65 | -3.10 | 0 |
| C16-C14 | 0 | -4.23 | -5.57 | -2.89 | 0 |
| C17-C14 | 0 | -2.96 | -4.40 | -1.51 | 6.09e-12 |
| C18-C14 | 0 | -0.0752 | -1.53 | 01.38 | 1 e+ 0 |
| C19-C14 | 0 | -9.74 | -11.2 | -8.25 | 0 |
| C20-C14 | 0 | 03.27 | 0,094444444 | 0,220833333 | 5.00e-13 |
| C21-C14 | 0 | -7.25 | -8.81 | -5.68 | 0 |
| C22-C14 | 0 | 0,634722222 | -0.655 | 02.48 | 9.43e- 1 |
| C23-C14 | 0 | -9.56 | -11.2 | -7.96 | 0 |
| C24-C14 | 0 | -0.364 | -1.96 | 01.24 | 1.00e+ 0 |
| C25-C14 | 0 | -12.0 | -13.7 | -10.4 | 0 |
| C26-C14 | 0 | -9.45 | -11.1 | -7.80 | 0 |
| C27-C14 | 0 | -3.12 | -4.79 | -1.45 | 9.54e-10 |
| C28-C14 | 0 | 03.44 | 0,0875 | 05.23 | 1.89e-10 |
| C29-C14 | 0 | -11.0 | -13.0 | -9.08 | 0 |
| C30-C14 | 0 | -7.02 | -8.99 | -5.05 | 0 |
| C31-C14 | 0 | -3.80 | -5.79 | -1.81 | 2.86e-10 |
| C16-C15 | 0 | 0,102083333 | -1.20 | 01.49 | 1 e+ 0 |
| C17-C15 | 0 | 01.42 | -0.0271 | 0,14375 | 6.33e- 2 |
| C18-C15 | 0 | 04.30 | 0,141666667 | 0,261111111 | 1.24e-13 |
| C19-C15 | 0 | -5.36 | -6.85 | -3.87 | 0 |
| C20-C15 | 0 | 0,336805556 | 06.14 | 09.16 | 0 |
| C21-C15 | 0 | -2.87 | -4.44 | -1.31 | 2.33e- 9 |
| C22-C15 | 0 | 05.29 | 0,175 | 0,309722222 | 0 |
| C23-C15 | 0 | -5.18 | -6.79 | -3.58 | 0 |
| C24-C15 | 0 | 04.01 | 02.41 | 0,251388889 | 2.86e-13 |
| C25-C15 | 0 | -7.66 | -9.30 | -6.01 | 0 |
| C26-C15 | 0 | -5.08 | -6.73 | -3.42 | 0 |
| C27-C15 | 0 | 01.25 | -0.422 | 0,147916667 | 5.49e- 1 |
| C28-C15 | 0 | 0,348611111 | 06.03 | 0,417361111 | 0 |
| C29-C15 | 0 | -6.65 | -8.59 | -4.70 | 0 |
| C30-C15 | 0 | -2.64 | -4.61 | -0.669 | 2.01e- 4 |
| C31-C15 | 0 | 0,397916667 | -1.42 | 02.56 | 1.00e+ 0 |
| C17-C16 | 0 | 01.27 | -0.234 | 0,1375 | 2.71e- 1 |
| C18-C16 | 0 | 04.15 | 0,127777778 | 0,254861111 | 3.14e-13 |
| C19-C16 | 0 | -5.51 | -7.05 | -3.96 | 0 |
| C20-C16 | 0 | 07.50 | 0,272916667 | 09.07 | 0 |
| C21-C16 | 0 | -3.02 | -4.64 | -1.40 | 1.10e- 9 |
| C22-C16 | 0 | 05.14 | 03.51 | 0,303472222 | 0 |
| C23-C16 | 0 | -5.33 | -6.99 | -3.67 | 0 |
| C24-C16 | 0 | 0,184722222 | 02.21 | 05.52 | 2.42e-13 |
| C25-C16 | 0 | -7.80 | -9.50 | -6.10 | 0 |
| C26-C16 | 0 | -5.22 | -6.93 | -3.52 | 0 |
| C27-C16 | 0 | 01.11 | -0.621 | 0,140972222 | 8.50e- 1 |
| C28-C16 | 0 | 0,338194444 | 0,265972222 | 09.51 | 0 |
| C29-C16 | 0 | -6.79 | -8.79 | -4.80 | 0 |
| C30-C16 | 0 | -2.79 | -4.80 | -0.772 | 8.57e- 5 |
| C31-C16 | 0 | 0,295833333 | -1.61 | 02.46 | 1.00e+ 0 |
| C18-C17 | 0 | 0,144444444 | 01.27 | 04.49 | 7.47e- 9 |
| C19-C17 | 0 | -6.78 | -8.42 | -5.14 | 0 |

| contrast | null.value | estimate | conf.low | conf.high | adj.p.value |
|---|---|---|---|---|---|
| C20-C17 | 0 | 06.23 | 04.57 | 0,352777778 | 0 |
| C21-C17 | 0 | -4.29 | -6.00 | -2.58 | 2.71e-13 |
| C22-C17 | 0 | 0,185416667 | 02.16 | 05.58 | 2.99e-13 |
| C23-C17 | 0 | -6.60 | -8.34 | -4.86 | 0 |
| C24-C17 | 0 | 02.59 | 0,588888889 | 04.33 | 9.90e- 6 |
| C25-C17 | 0 | -9.08 | -10.9 | -7.29 | 0 |
| C26-C17 | 0 | -6.50 | -8.29 | -4.71 | 0 |
| C27-C17 | 0 | -0.167 | -1.98 | 0,086111111 | 1 e+ 0 |
| C28-C17 | 0 | 06.40 | 04.48 | 08.31 | 0 |
| C29-C17 | 0 | -8.07 | -10.1 | -6.00 | 0 |
| C30-C17 | 0 | -4.06 | -6.15 | -1.97 | 1.10e-10 |
| C31-C17 | 0 | -0.847 | -2.95 | 01.26 | 1.00e+ 0 |
| C19-C18 | 0 | -9.66 | -11.3 | -8.01 | 0 |
| C20-C18 | 0 | 03.35 | 0,088888889 | 05.01 | 1.86e-11 |
| C21-C18 | 0 | -7.17 | -8.89 | -5.46 | 0 |
| C22-C18 | 0 | 0,686805556 | -0.734 | 0,132638889 | 9.52e- 1 |
| C23-C18 | 0 | -9.48 | -11.2 | -7.73 | 0 |
| C24-C18 | 0 | -0.289 | -2.04 | 01.46 | 1 e+ 0 |
| C25-C18 | 0 | -12.0 | -13.7 | -10.2 | 0 |
| C26-C18 | 0 | -9.38 | -11.2 | -7.58 | 0 |
| C27-C18 | 0 | -3.05 | -4.86 | -1.23 | 1.26e- 7 |
| C28-C18 | 0 | 03.52 | 01.59 | 05.44 | 2.68e- 9 |
| C29-C18 | 0 | -10.9 | -13.0 | -8.87 | 0 |
| C30-C18 | 0 | -6.94 | -9.03 | -4.85 | 0 |
| C31-C18 | 0 | -3.73 | -5.84 | -1.61 | 1.43e- 8 |
| C20-C19 | 0 | 13.00 | 11.03 | 14.07 | 0 |
| C21-C19 | 0 | 02.49 | 0,516666667 | 04.23 | 3.54e- 5 |
| C22-C19 | 0 | 10.06 | 0,395833333 | 12.04 | 0 |
| C23-C19 | 0 | 0,122916667 | -1.60 | 0,107638889 | 1 e+ 0 |
| C24-C19 | 0 | 09.37 | 07.59 | 11.01 | 0 |
| C25-C19 | 0 | -2.29 | -4.11 | -0.476 | 8.54e- 4 |
| C26-C19 | 0 | 0,197222222 | -1.54 | 02.11 | 1 e+ 0 |
| C27-C19 | 0 | 0,292361111 | 0,220138889 | 08.46 | 0 |
| C28-C19 | 0 | 13.02 | 11.02 | 15.01 | 0 |
| C29-C19 | 0 | -1.29 | -3.38 | 0,561111111 | 8.99e- 1 |
| C30-C19 | 0 | 0,133333333 | 0,41875 | 0,225 | 5.58e- 4 |
| C31-C19 | 0 | 0,272916667 | 0,180555556 | 08.07 | 2.91e-13 |
| C21-C20 | 0 | -10.5 | -12.3 | -8.76 | 0 |
| C22-C20 | 0 | -2.36 | -4.12 | -0.589 | 2.25e- 4 |
| C23-C20 | 0 | -12.8 | -14.6 | -11.0 | 0 |
| C24-C20 | 0 | -3.63 | -5.43 | -1.84 | 1.18e-11 |
| C25-C20 | 0 | -15.3 | -17.1 | -13.5 | 0 |
| C26-C20 | 0 | -12.7 | -14.6 | -10.9 | 0 |
| C27-C20 | 0 | -6.39 | -8.25 | -4.53 | 0 |
| C28-C20 | 0 | 0,11875 | -1.79 | 02.13 | 1 e+ 0 |
| C29-C20 | 0 | -14.3 | -16.4 | -12.2 | 0 |
| C30-C20 | 0 | -10.3 | -12.4 | -8.16 | 0 |
| C31-C20 | 0 | -7.07 | -9.22 | -4.92 | 0 |
| C22-C21 | 0 | 08.16 | 06.35 | 0,443055556 | 0 |
| C23-C21 | 0 | -2.31 | -4.15 | -0.469 | 9.66e- 4 |
| C24-C21 | 0 | 0,311111111 | 05.04 | 0,384027778 | 0 |
| C25-C21 | 0 | -4.78 | -6.67 | -2.90 | 2.43e-13 |

| contrast | null.value | estimate | conf.low | conf.high | adj.p.value |
|---|---|---|---|---|---|
| C26-C21 | 0 | -2.20 | -4.09 | -0.316 | 4.30e- 3 |
| C27-C21 | 0 | 04.13 | 02.22 | 06.03 | 5.42e-13 |
| C28-C21 | 0 | 10.07 | 0,380555556 | 12.07 | 0 |
| C29-C21 | 0 | -3.77 | -5.92 | -1.62 | 1.78e- 8 |
| C30-C21 | 0 | 0,161111111 | -1.94 | 02.40 | 1 e+ 0 |
| C31-C21 | 0 | 03.45 | 01.26 | 0,252083333 | 1.42e- 6 |
| C23-C22 | 0 | -10.5 | -12.3 | -8.63 | 0 |
| C24-C22 | 0 | -1.28 | -3.13 | 0,396527778 | 7.22e- 1 |
| C25-C22 | 0 | -12.9 | -14.8 | -11.1 | 0 |
| C26-C22 | 0 | -10.4 | -12.3 | -8.47 | 0 |
| C27-C22 | 0 | -4.04 | -5.95 | -2.13 | 1.26e-12 |
| C28-C22 | 0 | 02.53 | 0,358333333 | 04.54 | 9.36e- 4 |
| C29-C22 | 0 | -11.9 | -14.1 | -9.78 | 0 |
| C30-C22 | 0 | -7.93 | -10.1 | -5.75 | 0 |
| C31-C22 | 0 | -4.72 | -6.91 | -2.52 | 6.49e-13 |
| C24-C23 | 0 | 09.19 | 07.32 | 11.01 | 0 |
| C25-C23 | 0 | -2.47 | -4.38 | -0.559 | 4.88e- 4 |
| C26-C23 | 0 | 0,074305556 | -1.81 | 02.02 | 1 e+ 0 |
| C27-C23 | 0 | 06.44 | 04.50 | 08.37 | 0 |
| C28-C23 | 0 | 13.00 | 11.00 | 15.00 | 0 |
| C29-C23 | 0 | -1.46 | -3.64 | 0,495138889 | 7.74e- 1 |
| C30-C23 | 0 | 02.54 | 0,239583333 | 0,218055556 | 5.09e- 3 |
| C31-C23 | 0 | 0,261111111 | 03.54 | 0,359027778 | 3.28e-13 |
| C25-C24 | 0 | -11.7 | -13.6 | -9.75 | 0 |
| C26-C24 | 0 | -9.09 | -11.0 | -7.17 | 0 |
| C27-C24 | 0 | -2.76 | -4.69 | -0.822 | 3.68e- 5 |
| C28-C24 | 0 | 0,18125 | 0,095138889 | 0,266666667 | 9.20e-10 |
| C29-C24 | 0 | -10.7 | -12.8 | -8.48 | 0 |
| C30-C24 | 0 | -6.65 | -8.85 | -4.45 | 0 |
| C31-C24 | 0 | -3.44 | -5.65 | -1.22 | 2.37e- 6 |
| C26-C25 | 0 | 02.58 | 0,431944444 | 04.54 | 3.02e- 4 |
| C27-C25 | 0 | 0,396527778 | 0,314583333 | 10.09 | 0 |
| C28-C25 | 0 | 15.05 | 13.04 | 17.05 | 0 |
| C29-C25 | 0 | 01.01 | -1.20 | 03.22 | 9.98e- 1 |
| C30-C25 | 0 | 05.01 | 0,1375 | 07.25 | 3.14e-13 |
| C31-C25 | 0 | 08.23 | 0,276388889 | 10.05 | 0 |
| C27-C26 | 0 | 06.33 | 04.35 | 08.31 | 0 |
| C28-C26 | 0 | 12.09 | 10.08 | 15.00 | 0 |
| C29-C26 | 0 | -1.57 | -3.78 | 0,447222222 | 6.70e- 1 |
| C30-C26 | 0 | 02.44 | 0,138194444 | 0,213194444 | 1.45e- 2 |
| C31-C26 | 0 | 0,253472222 | 03.40 | 0,354166667 | 2.79e-13 |
| C28-C27 | 0 | 06.56 | 04.47 | 0,379166667 | 0 |
| C29-C27 | 0 | -7.90 | -10.1 | -5.67 | 0 |
| C30-C27 | 0 | -3.89 | -6.14 | -1.64 | 3.41e- 8 |
| C31-C27 | 0 | -0.680 | -2.95 | 01.59 | 1.00e+ 0 |
| C29-C28 | 0 | -14.5 | -16.8 | -12.1 | 0 |
| C30-C28 | 0 | -10.5 | -12.8 | -8.12 | 0 |
| C31-C28 | 0 | -7.24 | -9.60 | -4.89 | 0 |
| C30-C29 | 0 | 04.01 | 01.54 | 06.47 | 4.11e- 7 |
| C31-C29 | 0 | 07.22 | 0,218055556 | 0,423611111 | 2.62e-13 |
| C31-C30 | 0 | 03.21 | 0,499305556 | 0,257638889 | 5.28e- 4 |
| M-F | 0 | -2.75 | -2.91 | -2.58 | 0 |
