## Supplementary material for "Subgrouping multimorbid patients with ischemic heart disease by means of unsupervised clustering: A cohort study of 72,249 patients defined comprehensively by diagnoses prior to presentation": S5 Table

### S5A Table: Degree of enrichment (sum) and top-10 O/E-ratios per cluster

| Clst | Sum | ICD-10 | # code | # clst | Obs | Exp | O/E-ratio | description |
|---|---|---|---|---|---|---|---|---|
| C1 | 345 | I109 | 24818 | 7191 | 1.000 | 0.294 | 3.40 | Essential (primary) hypertension |
| C1 | 345 | H350 | 548 | 7191 | 0.016 | 0.007 | 2.24 | Background retinopathy and retinal vascular changes |
| C1 | 345 | E871 | 393 | 7191 | 0.012 | 0.005 | 2.23 | Hypo-osmolality and hyponatraemia |
| C1 | 345 | I159 | 414 | 7191 | 0.012 | 0.005 | 2.15 | Secondary hypertension, unspecified |
| C1 | 345 | I959 | 283 | 7191 | 0.008 | 0.004 | 2.15 | Hypotension, unspecified |
| C1 | 345 | E789 | 417 | 7191 | 0.012 | 0.006 | 2.10 | Disorder of lipoprotein metabolism, unspecified |
| C1 | 345 | E785 | 5002 | 7191 | 0.133 | 0.067 | 1.98 | Hyperlipidaemia, unspecified |
| C1 | 345 | I119 | 534 | 7191 | 0.013 | 0.007 | 1.85 | Hypertensive heart disease without (congestive) heart failure |
| C1 | 345 | D251 | 256 | 7191 | 0.006 | 0.004 | 1.83 | Intramural leiomyoma of uterus |
| C1 | 345 | N811 | 974 | 7191 | 0.024 | 0.013 | 1.77 | Cystocele |
| C2 | 372 | K802 | 2849 | 5990 | 0.176 | 0.029 | 6.01 | Calculus of gallbladder without cholecystitis |
| C2 | 372 | K801 | 411 | 5990 | 0.023 | 0.004 | 5.22 | Calculus of gallbladder with other cholecystitis |
| C2 | 372 | K805 | 1027 | 5990 | 0.054 | 0.011 | 4.70 | Calculus of bile duct without cholangitis or cholecystitis |
| C2 | 372 | K800 | 468 | 5990 | 0.022 | 0.006 | 3.88 | Calculus of gallbladder with acute cholecystitis |
| C2 | 372 | R100 | 2884 | 5990 | 0.123 | 0.035 | 3.51 | Acute abdomen |
| C2 | 372 | R108 | 3700 | 5990 | 0.158 | 0.045 | 3.51 | Abdominal and pelvic pain |
| C2 | 372 | R103 | 488 | 5990 | 0.020 | 0.006 | 3.29 | Pain localized to other parts of lower abdomen |
| C2 | 372 | R102 | 566 | 5990 | 0.023 | 0.007 | 3.29 | Pelvic and perineal pain |
| C2 | 372 | N832 | 489 | 5990 | 0.020 | 0.006 | 3.28 | Other and unspecified ovarian cysts |
| C2 | 372 | D251 | 256 | 5990 | 0.010 | 0.003 | 3.19 | Intramural leiomyoma of uterus |
| C3 | 268 | R079 | 5863 | 4641 | 0.363 | 0.067 | 5.43 | Pain in throat and chest |
| C3 | 268 | G409 | 841 | 4641 | 0.043 | 0.010 | 4.23 | Epilepsy, unspecified |
| C3 | 268 | I309 | 297 | 4641 | 0.014 | 0.004 | 3.70 | Acute pericarditis, unspecified |
| C3 | 268 | M626 | 4440 | 4641 | 0.187 | 0.057 | 3.28 | Muscle strain |
| C3 | 268 | G430 | 351 | 4641 | 0.013 | 0.005 | 2.83 | Migraine without aura [common migraine] |
| C3 | 268 | R073 | 2009 | 4641 | 0.072 | 0.027 | 2.67 | Other chest pain |
| C3 | 268 | R002 | 691 | 4641 | 0.025 | 0.009 | 2.66 | Palpitations |
| C3 | 268 | R519 | 1667 | 4641 | 0.058 | 0.022 | 2.61 | Headache |
| C3 | 268 | R064 | 542 | 4641 | 0.019 | 0.007 | 2.54 | Hyperventilation |
| C3 | 268 | R072 | 367 | 4641 | 0.012 | 0.005 | 2.48 | Precordial pain |
| C4 | 520 | I489 | 7075 | 4401 | 0.995 | 0.043 | 23.14 | Atrial fibrillation and atrial flutter, unspecified |
| C4 | 520 | I495 | 482 | 4401 | 0.055 | 0.004 | 14.14 | Sick sinus syndrome |
| C4 | 520 | I480 | 364 | 4401 | 0.039 | 0.003 | 12.49 | Paroxysmal atrial fibrillation |
| C4 | 520 | I471 | 1920 | 4401 | 0.183 | 0.018 | 10.27 | Supraventricular tachycardia |
| C4 | 520 | I499 | 436 | 4401 | 0.035 | 0.005 | 7.71 | Cardiac arrhythmia, unspecified |
| C4 | 520 | I479 | 614 | 4401 | 0.045 | 0.007 | 6.73 | Paroxysmal tachycardia, unspecified |
| C4 | 520 | R001 | 394 | 4401 | 0.026 | 0.004 | 5.80 | Bradycardia, unspecified |
| C4 | 520 | R000 | 391 | 4401 | 0.025 | 0.004 | 5.51 | Tachycardia, unspecified |
| C4 | 520 | I340 | 971 | 4401 | 0.051 | 0.012 | 4.32 | Mitral (valve) insufficiency |
| C4 | 520 | I491 | 273 | 4401 | 0.013 | 0.003 | 3.76 | Atrial premature depolarization |
| C5 | 596 | E119 | 7551 | 4290 | 0.947 | 0.056 | 17.06 | Type 2 diabetes mellitus: Without complications |
| C5 | 596 | E113 | 720 | 4290 | 0.081 | 0.006 | 13.55 | Type 2 diabetes mellitus: With ophthalmic complications |
| C5 | 596 | E114 | 881 | 4290 | 0.088 | 0.008 | 11.06 | Type 2 diabetes mellitus: With neurological complications |
| C5 | 596 | E149 | 958 | 4290 | 0.095 | 0.009 | 10.82 | Unspecified diabetes mellitus: Without complications |

| Clst | Sum | ICD-10 | # code | # clst | Obs | Exp | O/E-ratio | description |
|---|---|---|---|---|---|---|---|---|
| C5 | 596 | E117 | 980 | 4290 | 0.096 | 0.009 | 10.58 | Type 2 diabetes mellitus: With multiple complications |
| C5 | 596 | E118 | 2669 | 4290 | 0.251 | 0.025 | 9.93 | Type 2 diabetes mellitus: With unspecified complications |
| C5 | 596 | H360 | 1467 | 4290 | 0.137 | 0.014 | 9.80 | Diabetic retinopathy |
| C5 | 596 | E112 | 761 | 4290 | 0.070 | 0.007 | 9.53 | Type 2 diabetes mellitus: With renal complications |
| C5 | 596 | E148 | 483 | 4290 | 0.044 | 0.005 | 9.50 | Unspecified diabetes mellitus: With unspecified complications |
| C5 | 596 | E115 | 551 | 4290 | 0.048 | 0.005 | 8.88 | Type 2 diabetes mellitus: With peripheral circulatory complications |
| C6 | 239 | E780 | 12780 | 3589 | 0.867 | 0.152 | 5.70 | Pure hypercholesterolaemia |
| C6 | 239 | E785 | 5002 | 3589 | 0.177 | 0.069 | 2.57 | Hyperlipidaemia, unspecified |
| C6 | 239 | I999 | 383 | 3589 | 0.010 | 0.005 | 1.89 | Other and unspecified disorders of circulatory system |
| C6 | 239 | I639 | 1989 | 3589 | 0.044 | 0.029 | 1.53 | Cerebral infarction, unspecified |
| C6 | 239 | I652 | 563 | 3589 | 0.011 | 0.008 | 1.39 | Occlusion and stenosis of carotid artery |
| C6 | 239 | G459 | 2066 | 3589 | 0.042 | 0.030 | 1.39 | Transient cerebral ischaemic attack, unspecified |
| C6 | 239 | R670 | 750 | 3589 | 0.014 | 0.011 | 1.24 | NA |
| C6 | 239 | E113 | 720 | 3589 | 0.013 | 0.011 | 1.21 | Type 2 diabetes mellitus: With ophthalmic complications |
| C6 | 239 | M100 | 691 | 3589 | 0.012 | 0.010 | 1.20 | Idiopathic gout |
| C6 | 239 | N434 | 294 | 3589 | 0.005 | 0.004 | 1.16 | Spermatocele |
| C7 | 374 | M171 | 2940 | 3309 | 0.359 | 0.027 | 13.06 | Other primary gonarthrosis |
| C7 | 374 | M179 | 2242 | 3309 | 0.257 | 0.022 | 11.80 | Gonarthrosis, unspecified |
| C7 | 374 | M170 | 2145 | 3309 | 0.232 | 0.022 | 10.74 | Primary gonarthrosis, bilateral |
| C7 | 374 | M234 | 258 | 3309 | 0.027 | 0.003 | 10.33 | Loose body in knee |
| C7 | 374 | M235 | 269 | 3309 | 0.028 | 0.003 | 10.19 | Chronic instability of knee |
| C7 | 374 | M232 | 2404 | 3309 | 0.238 | 0.025 | 9.42 | Derangement of meniscus due to old tear or injury |
| C7 | 374 | M238 | 532 | 3309 | 0.042 | 0.006 | 6.89 | Other internal derangements of knee |
| C7 | 374 | M239 | 1105 | 3309 | 0.081 | 0.013 | 6.21 | Internal derangement of knee, unspecified |
| C7 | 374 | M712 | 363 | 3309 | 0.025 | 0.004 | 5.63 | Synovial cyst of popliteal space [Baker] |
| C7 | 374 | M169 | 1539 | 3309 | 0.088 | 0.020 | 4.48 | Coxarthrosis, unspecified |
| C8 | 384 | H919 | 4610 | 2802 | 0.664 | 0.043 | 15.54 | Hearing loss, unspecified |
| C8 | 384 | H911 | 3527 | 2802 | 0.495 | 0.033 | 14.90 | Presbycusis |
| C8 | 384 | H905 | 1160 | 2802 | 0.151 | 0.011 | 13.13 | Sensorineural hearing loss, unspecified |
| C8 | 384 | H833 | 1412 | 2802 | 0.180 | 0.014 | 12.70 | Noise effects on inner ear |
| C8 | 384 | H838 | 254 | 2802 | 0.032 | 0.003 | 12.38 | Other specified diseases of inner ear |
| C8 | 384 | H938 | 1116 | 2802 | 0.132 | 0.012 | 11.34 | Other specified disorders of ear |
| C8 | 384 | H908 | 631 | 2802 | 0.071 | 0.007 | 10.50 | Mixed conductive and sensorineural hearing loss, unspecified |
| C8 | 384 | H931 | 1228 | 2802 | 0.123 | 0.014 | 9.01 | Tinnitus |
| C8 | 384 | H810 | 300 | 2802 | 0.029 | 0.003 | 8.64 | Ménière disease |
| C8 | 384 | H809 | 374 | 2802 | 0.034 | 0.004 | 7.82 | Otosclerosis, unspecified |
| C9 | 323 | I420 | 706 | 2581 | 0.095 | 0.007 | 13.21 | Dilated cardiomyopathy |
| C9 | 323 | I509 | 6160 | 2581 | 0.783 | 0.064 | 12.21 | Heart failure, unspecified |
| C9 | 323 | I429 | 479 | 2581 | 0.056 | 0.005 | 10.75 | Cardiomyopathy, unspecified |
| C9 | 323 | I501 | 1502 | 2581 | 0.124 | 0.018 | 6.74 | Left ventricular failure |
| C9 | 323 | I500 | 2327 | 2581 | 0.188 | 0.029 | 6.57 | Congestive heart failure |
| C9 | 323 | R570 | 320 | 2581 | 0.025 | 0.004 | 6.38 | Cardiogenic shock |
| C9 | 323 | I460 | 1028 | 2581 | 0.075 | 0.013 | 5.78 | Cardiac arrest with successful resuscitation |
| C9 | 323 | I472 | 752 | 2581 | 0.053 | 0.010 | 5.52 | Ventricular tachycardia |

| Clst | Sum | ICD-10 | # code | # clst | Obs | Exp | O/E-ratio | description |
|---|---|---|---|---|---|---|---|---|
| C9 | 323 | I110 | 273 | 2581 | 0.018 | 0.004 | 5.20 | Hypertensive heart disease with (congestive) heart failure |
| C9 | 323 | I340 | 971 | 2581 | 0.059 | 0.013 | 4.68 | Mitral (valve) insufficiency |
| C10 | 361 | H259 | 5764 | 2562 | 0.866 | 0.055 | 15.77 | Senile cataract, unspecified |
| C10 | 361 | H264 | 1015 | 2562 | 0.113 | 0.011 | 10.03 | After-cataract |
| C10 | 361 | H330 | 418 | 2562 | 0.045 | 0.005 | 9.68 | Retinal detachment with retinal break |
| C10 | 361 | H353 | 1743 | 2562 | 0.174 | 0.020 | 8.67 | Degeneration of macula and posterior pole |
| C10 | 361 | H401 | 388 | 2562 | 0.032 | 0.005 | 6.86 | Primary open-angle glaucoma |
| C10 | 361 | H438 | 291 | 2562 | 0.022 | 0.004 | 6.14 | Other disorders of vitreous body |
| C10 | 361 | H333 | 264 | 2562 | 0.019 | 0.003 | 5.74 | Retinal breaks without detachment |
| C10 | 361 | H521 | 366 | 2562 | 0.023 | 0.005 | 4.84 | Myopia |
| C10 | 361 | H348 | 337 | 2562 | 0.017 | 0.005 | 3.79 | Other retinal vascular occlusions |
| C10 | 361 | H260 | 486 | 2562 | 0.023 | 0.007 | 3.55 | Infantile, juvenile and presenile cataract |
| C11 | 328 | K409 | 3787 | 2292 | 0.984 | 0.024 | 41.64 | Unilateral or unspecified inguinal hernia, without obstruction or gangrene |
| C11 | 328 | K402 | 280 | 2292 | 0.042 | 0.003 | 14.76 | Bilateral inguinal hernia, without obstruction or gangrene |
| C11 | 328 | N433 | 367 | 2292 | 0.023 | 0.005 | 4.67 | Hydrocele, unspecified |
| C11 | 328 | N434 | 294 | 2292 | 0.010 | 0.004 | 2.40 | Spermatocele |
| C11 | 328 | K429 | 896 | 2292 | 0.030 | 0.013 | 2.32 | Umbilical hernia without obstruction or gangrene |
| C11 | 328 | N484 | 456 | 2292 | 0.011 | 0.007 | 1.71 | Impotence of organic origin |
| C11 | 328 | N508 | 300 | 2292 | 0.007 | 0.004 | 1.70 | Other specified disorders of male genital organs |
| C11 | 328 | M720 | 1004 | 2292 | 0.024 | 0.015 | 1.67 | Palmar fascial fibromatosis [Dupuytren] |
| C11 | 328 | D179 | 257 | 2292 | 0.006 | 0.004 | 1.63 | Benign lipomatous neoplasm, unspecified |
| C11 | 328 | I714 | 517 | 2292 | 0.012 | 0.008 | 1.56 | Abdominal aortic aneurysm, without mention of rupture |
| C12 | 383 | N409 | 3319 | 2213 | 0.701 | 0.027 | 25.77 | Hyperplasia of prostate |
| C12 | 383 | R339 | 1530 | 2213 | 0.239 | 0.015 | 15.55 | Retention of urine |
| C12 | 383 | R391 | 2230 | 2213 | 0.241 | 0.026 | 9.24 | Other difficulties with micturition |
| C12 | 383 | N359 | 253 | 2213 | 0.027 | 0.003 | 9.12 | Urethral stricture, unspecified |
| C12 | 383 | R319 | 3787 | 2213 | 0.323 | 0.047 | 6.82 | Unspecified haematuria |
| C12 | 383 | N459 | 612 | 2213 | 0.052 | 0.008 | 6.72 | Orchitis, epididymitis and epididymo-orchitis without abscess |
| C12 | 383 | M720 | 1004 | 2213 | 0.059 | 0.013 | 4.36 | Palmar fascial fibromatosis [Dupuytren] |
| C12 | 383 | N434 | 294 | 2213 | 0.015 | 0.004 | 3.71 | Spermatocele |
| C12 | 383 | N309 | 834 | 2213 | 0.042 | 0.011 | 3.64 | Cystitis, unspecified |
| C12 | 383 | N319 | 265 | 2213 | 0.013 | 0.004 | 3.60 | Neuromuscular dysfunction of bladder, unspecified |
| C13 | 522 | M511 | 3357 | 2070 | 0.937 | 0.022 | 42.98 | Lumbar and other intervertebral disc disorders with radiculopathy |
| C13 | 522 | M519 | 604 | 2070 | 0.120 | 0.005 | 22.05 | Intervertebral disc disorder, unspecified |
| C13 | 522 | M512 | 366 | 2070 | 0.071 | 0.003 | 21.34 | Other specified intervertebral disc displacement |
| C13 | 522 | M544 | 1209 | 2070 | 0.144 | 0.014 | 10.28 | Lumbago with sciatica |
| C13 | 522 | M543 | 491 | 2070 | 0.057 | 0.006 | 9.94 | Sciatica |
| C13 | 522 | M513 | 1585 | 2070 | 0.178 | 0.019 | 9.54 | Other specified intervertebral disc degeneration |
| C13 | 522 | M539 | 274 | 2070 | 0.026 | 0.003 | 7.54 | Dorsopathy, unspecified |
| C13 | 522 | M472 | 897 | 2070 | 0.082 | 0.011 | 7.35 | Other spondylosis with radiculopathy |
| C13 | 522 | M501 | 910 | 2070 | 0.080 | 0.011 | 7.01 | Cervical disc disorder with radiculopathy |
| C13 | 522 | M549 | 1419 | 2070 | 0.103 | 0.019 | 5.58 | Dorsalgia, unspecified |
| C14 | 676 | J440 | 743 | 2040 | 0.191 | 0.005 | 35.25 | Chronic obstructive pulmonary disease with acute lower respiratory infection |

| Clst | Sum | ICD-10 | # code | # clst | Obs | Exp | O/E-ratio | description |
|---|---|---|---|---|---|---|---|---|
| C14 | 676 | J441 | 1678 | 2040 | 0.423 | 0.013 | 33.71 | Chronic obstructive pulmonary disease with acute exacerbation, unspecified |
| C14 | 676 | J449 | 4621 | 2040 | 0.948 | 0.041 | 22.97 | Chronic obstructive pulmonary disease, unspecified |
| C14 | 676 | J439 | 317 | 2040 | 0.064 | 0.003 | 22.18 | Emphysema, unspecified |
| C14 | 676 | J448 | 396 | 2040 | 0.075 | 0.004 | 20.31 | Other specified chronic obstructive pulmonary disease |
| C14 | 676 | J429 | 714 | 2040 | 0.105 | 0.008 | 13.66 | Unspecified chronic bronchitis |
| C14 | 676 | J960 | 837 | 2040 | 0.120 | 0.009 | 13.13 | Acute respiratory failure |
| C14 | 676 | J969 | 681 | 2040 | 0.072 | 0.008 | 8.78 | Respiratory failure, unspecified |
| C14 | 676 | J159 | 1222 | 2040 | 0.103 | 0.016 | 6.62 | Bacterial pneumonia, unspecified |
| C14 | 676 | J209 | 527 | 2040 | 0.041 | 0.007 | 6.05 | Acute bronchitis, unspecified |
| C15 | 296 | E780 | 12780 | 2013 | 1.000 | 0.165 | 6.05 | Pure hypercholesterolaemia |
| C15 | 296 | I109 | 24818 | 2013 | 1.000 | 0.350 | 2.86 | Essential (primary) hypertension |
| C15 | 296 | D629 | 333 | 2013 | 0.009 | 0.005 | 1.96 | Acute posthaemorrhagic anaemia |
| C15 | 296 | I639 | 1989 | 2013 | 0.055 | 0.029 | 1.91 | Cerebral infarction, unspecified |
| C15 | 296 | I693 | 467 | 2013 | 0.013 | 0.007 | 1.91 | Sequelae of cerebral infarction |
| C15 | 296 | R670 | 750 | 2013 | 0.020 | 0.011 | 1.87 | NA |
| C15 | 296 | E114 | 881 | 2013 | 0.021 | 0.013 | 1.66 | Type 2 diabetes mellitus: With neurological complications |
| C15 | 296 | G459 | 2066 | 2013 | 0.049 | 0.030 | 1.61 | Transient cerebral ischaemic attack, unspecified |
| C15 | 296 | E118 | 2669 | 2013 | 0.063 | 0.039 | 1.60 | Type 2 diabetes mellitus: With unspecified complications |
| C15 | 296 | R072 | 367 | 2013 | 0.008 | 0.005 | 1.57 | Precordial pain |
| C16 | 365 | J189 | 5496 | 1654 | 0.743 | 0.065 | 11.40 | Pneumonia, unspecified |
| C16 | 365 | J849 | 265 | 1654 | 0.027 | 0.003 | 8.10 | Interstitial pulmonary disease, unspecified |
| C16 | 365 | C349 | 284 | 1654 | 0.026 | 0.004 | 7.06 | Malignant neoplasm: Bronchus or lung, unspecified |
| C16 | 365 | R919 | 1471 | 1654 | 0.103 | 0.020 | 5.17 | Abnormal findings on diagnostic imaging of lung |
| C16 | 365 | J909 | 291 | 1654 | 0.019 | 0.004 | 4.72 | Pleural effusion, not elsewhere classified |
| C16 | 365 | J181 | 298 | 1654 | 0.018 | 0.004 | 4.43 | Lobar pneumonia, unspecified |
| C16 | 365 | J159 | 1222 | 1654 | 0.073 | 0.017 | 4.35 | Bacterial pneumonia, unspecified |
| C16 | 365 | R091 | 418 | 1654 | 0.023 | 0.006 | 3.96 | Pleurisy |
| C16 | 365 | J180 | 257 | 1654 | 0.013 | 0.004 | 3.71 | Bronchopneumonia, unspecified |
| C16 | 365 | J969 | 681 | 1654 | 0.031 | 0.010 | 3.27 | Respiratory failure, unspecified |
| C17 | 375 | E780 | 12780 | 1281 | 1.000 | 0.175 | 5.73 | Pure hypercholesterolaemia |
| C17 | 375 | E789 | 417 | 1281 | 0.018 | 0.006 | 3.00 | Disorder of lipoprotein metabolism, unspecified |
| C17 | 375 | H350 | 548 | 1281 | 0.023 | 0.008 | 2.98 | Background retinopathy and retinal vascular changes |
| C17 | 375 | G459 | 2066 | 1281 | 0.087 | 0.030 | 2.95 | Transient cerebral ischaemic attack, unspecified |
| C17 | 375 | I639 | 1989 | 1281 | 0.084 | 0.029 | 2.92 | Cerebral infarction, unspecified |
| C17 | 375 | I109 | 24818 | 1281 | 1.000 | 0.357 | 2.80 | Essential (primary) hypertension |
| C17 | 375 | I652 | 563 | 1281 | 0.022 | 0.008 | 2.69 | Occlusion and stenosis of carotid artery |
| C17 | 375 | I693 | 467 | 1281 | 0.017 | 0.007 | 2.54 | Sequelae of cerebral infarction |
| C17 | 375 | M109 | 547 | 1281 | 0.019 | 0.008 | 2.36 | Gout, unspecified |
| C17 | 375 | I694 | 1733 | 1281 | 0.059 | 0.025 | 2.36 | Sequelae of stroke, not specified as haemorrhage or infarction |
| C18 | 535 | I702 | 2251 | 1251 | 0.812 | 0.019 | 43.33 | Atherosclerosis of arteries of extremities |
| C18 | 535 | I739 | 2027 | 1251 | 0.544 | 0.020 | 26.65 | Peripheral vascular disease, unspecified |
| C18 | 535 | I709 | 433 | 1251 | 0.081 | 0.005 | 16.02 | Generalized and unspecified atherosclerosis |
| C18 | 535 | L979 | 605 | 1251 | 0.091 | 0.007 | 12.23 | Ulcer of lower limb, not elsewhere classified |
| C18 | 535 | E105 | 416 | 1251 | 0.060 | 0.005 | 11.58 | Type 1 diabetes mellitus: With peripheral circulatory complications |
| C18 | 535 | E115 | 551 | 1251 | 0.066 | 0.007 | 9.34 | Type 2 diabetes mellitus: With peripheral circulatory complications |

| Clst | Sum | ICD-10 | # code | # clst | Obs | Exp | O/E-ratio | description |
|---|---|---|---|---|---|---|---|---|
| C18 | 535 | E148 | 483 | 1251 | 0.042 | 0.007 | 6.35 | Unspecified diabetes mellitus: With unspecified complications |
| C18 | 535 | I714 | 517 | 1251 | 0.043 | 0.007 | 6.14 | Abdominal aortic aneurysm, without mention of rupture |
| C18 | 535 | L984 | 299 | 1251 | 0.021 | 0.004 | 5.02 | Chronic ulcer of skin, not elsewhere classified |
| C18 | 535 | I999 | 383 | 1251 | 0.023 | 0.005 | 4.31 | Other and unspecified disorders of circulatory system |
| C19 | 374 | G473 | 1897 | 1168 | 0.712 | 0.016 | 44.12 | Sleep apnoea |
| C19 | 374 | R065 | 983 | 1168 | 0.271 | 0.010 | 26.88 | Mouth breathing |
| C19 | 374 | J342 | 1097 | 1168 | 0.218 | 0.013 | 17.10 | Deviated nasal septum |
| C19 | 374 | J330 | 351 | 1168 | 0.038 | 0.005 | 8.10 | Polyp of nasal cavity |
| C19 | 374 | J320 | 513 | 1168 | 0.033 | 0.007 | 4.65 | Chronic maxillary sinusitis |
| C19 | 374 | J350 | 315 | 1168 | 0.015 | 0.005 | 3.22 | Chronic tonsillitis |
| C19 | 374 | N508 | 300 | 1168 | 0.014 | 0.004 | 3.18 | Other specified disorders of male genital organs |
| C19 | 374 | E669 | 2114 | 1168 | 0.086 | 0.031 | 2.80 | Obesity, unspecified |
| C19 | 374 | M766 | 339 | 1168 | 0.013 | 0.005 | 2.62 | Achilles tendinitis |
| C19 | 374 | G510 | 365 | 1168 | 0.013 | 0.005 | 2.42 | Bell’s palsy |
| C20 | 337 | I350 | 2664 | 1119 | 0.856 | 0.026 | 33.13 | Aortic (valve) stenosis |
| C20 | 337 | I351 | 696 | 1119 | 0.090 | 0.009 | 10.02 | Aortic (valve) insufficiency |
| C20 | 337 | K053 | 360 | 1119 | 0.023 | 0.005 | 4.59 | Chronic periodontitis |
| C20 | 337 | K045 | 333 | 1119 | 0.021 | 0.005 | 4.58 | Chronic apical periodontitis |
| C20 | 337 | R040 | 1768 | 1119 | 0.100 | 0.025 | 3.99 | Epistaxis |
| C20 | 337 | D649 | 1637 | 1119 | 0.084 | 0.023 | 3.59 | Anaemia, unspecified |
| C20 | 337 | I340 | 971 | 1119 | 0.045 | 0.014 | 3.20 | Mitral (valve) insufficiency |
| C20 | 337 | M353 | 627 | 1119 | 0.028 | 0.009 | 3.07 | Polymyalgia rheumatica |
| C20 | 337 | D509 | 459 | 1119 | 0.020 | 0.007 | 2.97 | Iron deficiency anaemia, unspecified |
| C20 | 337 | K921 | 287 | 1119 | 0.012 | 0.004 | 2.80 | Melaena |
| C21 | 532 | N200 | 1391 | 1000 | 0.765 | 0.009 | 80.82 | Calculus of kidney |
| C21 | 532 | N201 | 1381 | 1000 | 0.605 | 0.012 | 51.56 | Calculus of ureter |
| C21 | 532 | N209 | 520 | 1000 | 0.202 | 0.005 | 42.01 | Urinary calculus, unspecified |
| C21 | 532 | N133 | 267 | 1000 | 0.045 | 0.003 | 13.41 | Other and unspecified hydronephrosis |
| C21 | 532 | N109 | 555 | 1000 | 0.047 | 0.008 | 6.12 | Acute tubulo-interstitial nephritis |
| C21 | 532 | R319 | 3787 | 1000 | 0.197 | 0.054 | 3.63 | Unspecified haematuria |
| C21 | 532 | N359 | 253 | 1000 | 0.009 | 0.004 | 2.44 | Urethral stricture, unspecified |
| C21 | 532 | N308 | 384 | 1000 | 0.013 | 0.006 | 2.32 | Other cystitis |
| C21 | 532 | R100 | 2884 | 1000 | 0.096 | 0.042 | 2.28 | Acute abdomen |
| C21 | 532 | A419 | 968 | 1000 | 0.032 | 0.014 | 2.26 | Sepsis, unspecified |
| C22 | 592 | M480 | 2424 | 988 | 0.932 | 0.023 | 41.03 | Spinal stenosis |
| C22 | 592 | M431 | 583 | 988 | 0.135 | 0.007 | 19.79 | Spondylolisthesis |
| C22 | 592 | M472 | 897 | 988 | 0.159 | 0.011 | 14.20 | Other spondylosis with radiculopathy |
| C22 | 592 | M513 | 1585 | 988 | 0.200 | 0.021 | 9.56 | Other specified intervertebral disc degeneration |
| C22 | 592 | M539 | 274 | 988 | 0.031 | 0.004 | 8.54 | Dorsopathy, unspecified |
| C22 | 592 | M478 | 630 | 988 | 0.072 | 0.008 | 8.50 | Other spondylosis |
| C22 | 592 | M479 | 531 | 988 | 0.059 | 0.007 | 8.21 | Spondylosis, unspecified |
| C22 | 592 | M543 | 491 | 988 | 0.046 | 0.007 | 6.76 | Sciatica |
| C22 | 592 | M503 | 520 | 988 | 0.039 | 0.007 | 5.43 | Other cervical disc degeneration |
| C22 | 592 | R522 | 1177 | 988 | 0.088 | 0.016 | 5.34 | Other chronic pain |
| C23 | 968 | E103 | 576 | 935 | 0.339 | 0.004 | 86.66 | Type 1 diabetes mellitus: With ophthalmic complications |
| C23 | 968 | E107 | 583 | 935 | 0.303 | 0.005 | 66.79 | Type 1 diabetes mellitus: With multiple complications |

| Clst | Sum | ICD-10 | # code | # clst | Obs | Exp | O/E-ratio | description |
|---|---|---|---|---|---|---|---|---|
| C23 | 968 | E104 | 434 | 935 | 0.214 | 0.004 | 60.52 | Type 1 diabetes mellitus: With neurological complications |
| C23 | 968 | E102 | 377 | 935 | 0.165 | 0.003 | 48.90 | Type 1 diabetes mellitus: With renal complications |
| C23 | 968 | E162 | 510 | 935 | 0.199 | 0.005 | 40.65 | Hypoglycaemia, unspecified |
| C23 | 968 | E108 | 1073 | 935 | 0.363 | 0.011 | 32.70 | Type 1 diabetes mellitus: With unspecified complications |
| C23 | 968 | E109 | 2680 | 935 | 0.887 | 0.028 | 31.71 | Type 1 diabetes mellitus: Without complications |
| C23 | 968 | E105 | 416 | 935 | 0.137 | 0.004 | 31.47 | Type 1 diabetes mellitus: With peripheral circulatory complications |
| C23 | 968 | H360 | 1467 | 935 | 0.398 | 0.017 | 24.05 | Diabetic retinopathy |
| C23 | 968 | H431 | 334 | 935 | 0.061 | 0.004 | 14.57 | Vitreous haemorrhage |
| C24 | 559 | C509 | 1103 | 932 | 0.835 | 0.005 | 170.04 | Malignant neoplasm: Breast, unspecified |
| C24 | 559 | N639 | 773 | 932 | 0.153 | 0.010 | 16.12 | Unspecified lump in breast |
| C24 | 559 | D249 | 758 | 932 | 0.097 | 0.010 | 9.57 | Benign neoplasm of breast |
| C24 | 559 | N602 | 712 | 932 | 0.076 | 0.010 | 7.87 | Fibroadenosis of breast |
| C24 | 559 | N950 | 699 | 932 | 0.059 | 0.010 | 6.07 | Postmenopausal bleeding |
| C24 | 559 | L905 | 295 | 932 | 0.020 | 0.004 | 4.89 | Scar conditions and fibrosis of skin |
| C24 | 559 | M819 | 1292 | 932 | 0.083 | 0.018 | 4.50 | Osteoporosis, unspecified |
| C24 | 559 | N629 | 571 | 932 | 0.036 | 0.008 | 4.50 | Hypertrophy of breast |
| C24 | 559 | N840 | 546 | 932 | 0.030 | 0.008 | 3.84 | Polyp of corpus uteri |
| C24 | 559 | E052 | 322 | 932 | 0.014 | 0.005 | 2.99 | Thyrotoxicosis with toxic multinodular goitre |
| C25 | 681 | F103 | 518 | 860 | 0.331 | 0.004 | 94.26 | Mental and behavioural disorders due to use of alcohol: Withdrawal state |
| C25 | 681 | F102 | 1189 | 860 | 0.641 | 0.010 | 66.56 | Mental and behavioural disorders due to use of alcohol: Dependence syndrome |
| C25 | 681 | F100 | 1212 | 860 | 0.535 | 0.011 | 47.14 | Mental and behavioural disorders due to use of alcohol: Acute intoxication |
| C25 | 681 | F101 | 1115 | 860 | 0.430 | 0.011 | 38.27 | Mental and behavioural disorders due to use of alcohol: Harmful use |
| C25 | 681 | F172 | 349 | 860 | 0.090 | 0.004 | 21.82 | Mental and behavioural disorders due to use of tobacco: Dependence syndrome |
| C25 | 681 | F339 | 299 | 860 | 0.053 | 0.004 | 14.01 | Recurrent depressive disorder, unspecified |
| C25 | 681 | R568 | 477 | 860 | 0.067 | 0.006 | 10.67 | Other and unspecified convulsions |
| C25 | 681 | K920 | 428 | 860 | 0.057 | 0.006 | 9.96 | Haematemesis |
| C25 | 681 | F329 | 816 | 860 | 0.093 | 0.011 | 8.38 | Depressive episode, unspecified |
| C25 | 681 | F419 | 335 | 860 | 0.036 | 0.005 | 7.86 | Anxiety disorder, unspecified |
| C26 | 540 | J459 | 2487 | 852 | 0.927 | 0.026 | 36.22 | Asthma, unspecified |
| C26 | 540 | J451 | 417 | 852 | 0.133 | 0.005 | 28.92 | Nonallergic asthma |
| C26 | 540 | J450 | 401 | 852 | 0.127 | 0.004 | 28.68 | Predominantly allergic asthma |
| C26 | 540 | J448 | 396 | 852 | 0.040 | 0.005 | 7.31 | Other specified chronic obstructive pulmonary disease |
| C26 | 540 | J209 | 527 | 852 | 0.046 | 0.007 | 6.22 | Acute bronchitis, unspecified |
| C26 | 540 | J330 | 351 | 852 | 0.029 | 0.005 | 5.97 | Polyp of nasal cavity |
| C26 | 540 | J370 | 343 | 852 | 0.028 | 0.005 | 5.85 | Chronic laryngitis |
| C26 | 540 | R490 | 415 | 852 | 0.023 | 0.006 | 3.94 | Dysphonia |
| C26 | 540 | J429 | 714 | 852 | 0.039 | 0.010 | 3.77 | Unspecified chronic bronchitis |
| C26 | 540 | J441 | 1678 | 852 | 0.083 | 0.024 | 3.44 | Chronic obstructive pulmonary disease with acute exacerbation, unspecified |
| C27 | 440 | I649 | 2991 | 823 | 0.666 | 0.037 | 18.07 | Stroke, not specified as haemorrhage or infarction |
| C27 | 440 | I694 | 1733 | 823 | 0.360 | 0.022 | 16.60 | Sequelae of stroke, not specified as haemorrhage or infarction |
| C27 | 440 | I693 | 467 | 823 | 0.079 | 0.006 | 13.03 | Sequelae of cerebral infarction |

| Clst | Sum | ICD-10 | # code | # clst | Obs | Exp | O/E-ratio | description |
|---|---|---|---|---|---|---|---|---|
| C27 | 440 | I639 | 1989 | 823 | 0.332 | 0.026 | 12.82 | Cerebral infarction, unspecified |
| C27 | 440 | I652 | 563 | 823 | 0.085 | 0.007 | 11.44 | Occlusion and stenosis of carotid artery |
| C27 | 440 | G459 | 2066 | 823 | 0.270 | 0.028 | 9.70 | Transient cerebral ischaemic attack, unspecified |
| C27 | 440 | R670 | 750 | 823 | 0.077 | 0.010 | 7.39 | NA |
| C27 | 440 | G409 | 841 | 823 | 0.080 | 0.012 | 6.86 | Epilepsy, unspecified |
| C27 | 440 | G969 | 710 | 823 | 0.055 | 0.010 | 5.45 | Disorder of central nervous system, unspecified |
| C27 | 440 | R298 | 1366 | 823 | 0.104 | 0.019 | 5.41 | Other and unspecified symptoms and signs involving the nervous and musculoskeletal systems |
| C28 | 483 | C619 | 1173 | 686 | 0.943 | 0.008 | 119.15 | Malignant neoplasm of prostate |
| C28 | 483 | R339 | 1530 | 686 | 0.131 | 0.022 | 6.05 | Retention of urine |
| C28 | 483 | N484 | 456 | 686 | 0.031 | 0.007 | 4.68 | Impotence of organic origin |
| C28 | 483 | C443 | 522 | 686 | 0.026 | 0.008 | 3.46 | Malignant neoplasm: Skin of other and unspecified parts of face |
| C28 | 483 | N133 | 267 | 686 | 0.013 | 0.004 | 3.38 | Other and unspecified hydronephrosis |
| C28 | 483 | N479 | 462 | 686 | 0.022 | 0.007 | 3.25 | Redundant prepuce, phimosis and paraphimosis |
| C28 | 483 | N409 | 3319 | 686 | 0.147 | 0.048 | 3.04 | Hyperplasia of prostate |
| C28 | 483 | R391 | 2230 | 686 | 0.096 | 0.033 | 2.95 | Other difficulties with micturition |
| C28 | 483 | A419 | 968 | 686 | 0.039 | 0.014 | 2.78 | Sepsis, unspecified |
| C28 | 483 | N359 | 253 | 686 | 0.010 | 0.004 | 2.76 | Urethral stricture, unspecified |
| C29 | 264 | A630 | 320 | 550 | 0.082 | 0.004 | 19.81 | Anogenital (venereal) warts |
| C29 | 264 | L022 | 506 | 550 | 0.076 | 0.007 | 10.96 | Cutaneous abscess, furuncle and carbuncle of trunk |
| C29 | 264 | L024 | 953 | 550 | 0.135 | 0.013 | 10.19 | Cutaneous abscess, furuncle and carbuncle of limb |
| C29 | 264 | L089 | 1001 | 550 | 0.113 | 0.014 | 7.99 | Local infection of skin and subcutaneous tissue, unspecified |
| C29 | 264 | L029 | 622 | 550 | 0.069 | 0.009 | 7.88 | Cutaneous abscess, furuncle and carbuncle, unspecified |
| C29 | 264 | I803 | 1509 | 550 | 0.142 | 0.021 | 6.60 | Phlebitis and thrombophlebitis of lower extremities, unspecified |
| C29 | 264 | A499 | 492 | 550 | 0.044 | 0.007 | 6.21 | Bacterial infection, unspecified |
| C29 | 264 | A469 | 1352 | 550 | 0.109 | 0.019 | 5.62 | Erysipelas |
| C29 | 264 | I829 | 297 | 550 | 0.016 | 0.004 | 3.78 | Embolism and thrombosis of unspecified vein |
| C29 | 264 | R509 | 907 | 550 | 0.047 | 0.013 | 3.57 | Fever, unspecified |
| C30 | 1434 | N180 | 454 | 533 | 0.610 | 0.002 | 314.82 | Chronic kidney disease |
| C30 | 1434 | N199 | 711 | 533 | 0.585 | 0.006 | 97.71 | Unspecified kidney failure |
| C30 | 1434 | N189 | 1094 | 533 | 0.848 | 0.010 | 87.98 | Chronic kidney disease, unspecified |
| C30 | 1434 | K650 | 264 | 533 | 0.148 | 0.003 | 53.36 | Acute peritonitis |
| C30 | 1434 | E102 | 377 | 533 | 0.135 | 0.005 | 29.50 | Type 1 diabetes mellitus: With renal complications |
| C30 | 1434 | N179 | 377 | 533 | 0.086 | 0.005 | 17.37 | Acute renal failure, unspecified |
| C30 | 1434 | E112 | 761 | 533 | 0.167 | 0.010 | 16.55 | Type 2 diabetes mellitus: With renal complications |
| C30 | 1434 | K053 | 360 | 533 | 0.077 | 0.005 | 16.06 | Chronic periodontitis |
| C30 | 1434 | E107 | 583 | 533 | 0.120 | 0.008 | 15.41 | Type 1 diabetes mellitus: With multiple complications |
| C30 | 1434 | N133 | 267 | 533 | 0.045 | 0.004 | 12.34 | Other and unspecified hydronephrosis |
| C31 | 727 | M059 | 682 | 520 | 0.679 | 0.005 | 137.45 | Seropositive rheumatoid arthritis, unspecified |
| C31 | 727 | M069 | 662 | 520 | 0.567 | 0.006 | 102.97 | Rheumatoid arthritis, unspecified |
| C31 | 727 | M060 | 398 | 520 | 0.246 | 0.004 | 60.73 | Seronegative rheumatoid arthritis |
| C31 | 727 | M204 | 350 | 520 | 0.063 | 0.005 | 13.34 | Other hammer toe(s) (acquired) |
| C31 | 727 | M139 | 488 | 520 | 0.067 | 0.007 | 9.90 | Arthritis, unspecified |
| C31 | 727 | M029 | 304 | 520 | 0.031 | 0.004 | 7.12 | Reactive arthropathy, unspecified |
| C31 | 727 | J849 | 265 | 520 | 0.019 | 0.004 | 5.02 | Interstitial pulmonary disease, unspecified |
| C31 | 727 | M201 | 858 | 520 | 0.060 | 0.012 | 4.80 | Hallux valgus (acquired) |

| Clst | Sum | ICD-10 | # code | # clst | Obs | Exp | O/E-ratio | description |
|---|---|---|---|---|---|---|---|---|
| C31 | 727 | M255 | 842 | 520 | 0.050 | 0.012 | 4.08 | Pain in joint |
| C31 | 727 | M190 | 837 | 520 | 0.048 | 0.012 | 3.94 | Primary arthrosis of other joints |

### S5B, Table: Bottom-10 O/E-ratios < 1 per cluster

| Clst | ICD-10 | # code | # clst | Obs | Exp | O/E-ratio | description |
|---|---|---|---|---|---|---|---|
| C1 | E107 | 583 | 7191 | 0.001 | 0.010 | 0.12 | Type 1 diabetes mellitus: With multiple complications |
| C1 | D303 | 600 | 7191 | 0.002 | 0.010 | 0.20 | Benign neoplasm: Bladder |
| C1 | E105 | 416 | 7191 | 0.001 | 0.007 | 0.20 | Type 1 diabetes mellitus: With peripheral circulatory complications |
| C1 | J441 | 1678 | 7191 | 0.006 | 0.027 | 0.21 | Chronic obstructive pulmonary disease with acute exacerbation, unspecified |
| C1 | E780 | 12780 | 7191 | 0.046 | 0.208 | 0.22 | Pure hypercholesterolaemia |
| C1 | E103 | 576 | 7191 | 0.002 | 0.009 | 0.22 | Type 1 diabetes mellitus: With ophthalmic complications |
| C1 | C619 | 1173 | 7191 | 0.004 | 0.019 | 0.23 | Malignant neoplasm of prostate |
| C1 | E102 | 377 | 7191 | 0.001 | 0.006 | 0.23 | Type 1 diabetes mellitus: With renal complications |
| C1 | H360 | 1467 | 7191 | 0.006 | 0.024 | 0.23 | Diabetic retinopathy |
| C1 | E104 | 434 | 7191 | 0.002 | 0.007 | 0.24 | Type 1 diabetes mellitus: With neurological complications |
| C2 | D303 | 600 | 5990 | 0.000 | 0.010 | 0.02 | Benign neoplasm: Bladder |
| C2 | C619 | 1173 | 5990 | 0.000 | 0.019 | 0.02 | Malignant neoplasm of prostate |
| C2 | E107 | 583 | 5990 | 0.000 | 0.010 | 0.02 | Type 1 diabetes mellitus: With multiple complications |
| C2 | E103 | 576 | 5990 | 0.000 | 0.009 | 0.02 | Type 1 diabetes mellitus: With ophthalmic complications |
| C2 | N180 | 454 | 5990 | 0.000 | 0.007 | 0.02 | Chronic kidney disease |
| C2 | H360 | 1467 | 5990 | 0.001 | 0.024 | 0.03 | Diabetic retinopathy |
| C2 | E108 | 1073 | 5990 | 0.001 | 0.017 | 0.03 | Type 1 diabetes mellitus: With unspecified complications |
| C2 | N409 | 3319 | 5990 | 0.002 | 0.054 | 0.03 | Hyperplasia of prostate |
| C2 | E104 | 434 | 5990 | 0.000 | 0.007 | 0.05 | Type 1 diabetes mellitus: With neurological complications |
| C2 | E148 | 483 | 5990 | 0.001 | 0.008 | 0.06 | Unspecified diabetes mellitus: With unspecified complications |
| C3 | E112 | 761 | 4641 | 0.000 | 0.012 | 0.02 | Type 2 diabetes mellitus: With renal complications |
| C3 | E113 | 720 | 4641 | 0.000 | 0.012 | 0.02 | Type 2 diabetes mellitus: With ophthalmic complications |
| C3 | D303 | 600 | 4641 | 0.000 | 0.010 | 0.02 | Benign neoplasm: Bladder |
| C3 | C619 | 1173 | 4641 | 0.000 | 0.019 | 0.02 | Malignant neoplasm of prostate |
| C3 | I350 | 2664 | 4641 | 0.001 | 0.043 | 0.03 | Aortic (valve) stenosis |
| C3 | M171 | 2940 | 4641 | 0.002 | 0.047 | 0.03 | Other primary gonarthrosis |
| C3 | J441 | 1678 | 4641 | 0.001 | 0.027 | 0.03 | Chronic obstructive pulmonary disease with acute exacerbation, unspecified |
| C3 | N409 | 3319 | 4641 | 0.002 | 0.053 | 0.04 | Hyperplasia of prostate |
| C3 | I509 | 6160 | 4641 | 0.004 | 0.098 | 0.04 | Heart failure, unspecified |
| C3 | M059 | 682 | 4641 | 0.000 | 0.011 | 0.04 | Seropositive rheumatoid arthritis, unspecified |
| C4 | N180 | 454 | 4401 | 0.002 | 0.007 | 0.26 | Chronic kidney disease |
| C4 | J350 | 315 | 4401 | 0.001 | 0.005 | 0.28 | Chronic tonsillitis |
| C4 | E105 | 416 | 4401 | 0.002 | 0.007 | 0.28 | Type 1 diabetes mellitus: With peripheral circulatory complications |
| C4 | E103 | 576 | 4401 | 0.003 | 0.009 | 0.30 | Type 1 diabetes mellitus: With ophthalmic complications |
| C4 | K801 | 411 | 4401 | 0.002 | 0.006 | 0.36 | Calculus of gallbladder with other cholecystitis |
| C4 | E107 | 583 | 4401 | 0.003 | 0.009 | 0.38 | Type 1 diabetes mellitus: With multiple complications |
| C4 | M235 | 269 | 4401 | 0.002 | 0.004 | 0.38 | Chronic instability of knee |

| Clst | ICD-10 | # code | # clst | Obs | Exp | O/E-ratio | description |
|---|---|---|---|---|---|---|---|
| C4 | G439 | 463 | 4401 | 0.003 | 0.007 | 0.41 | Migraine, unspecified |
| C4 | H360 | 1467 | 4401 | 0.010 | 0.023 | 0.43 | Diabetic retinopathy |
| C4 | J039 | 402 | 4401 | 0.003 | 0.006 | 0.44 | Acute tonsillitis, unspecified |
| C5 | D303 | 600 | 4290 | 0.003 | 0.009 | 0.27 | Benign neoplasm: Bladder |
| C5 | I495 | 482 | 4290 | 0.002 | 0.008 | 0.31 | Sick sinus syndrome |
| C5 | K402 | 280 | 4290 | 0.001 | 0.004 | 0.32 | Bilateral inguinal hernia, without obstruction or gangrene |
| C5 | I491 | 273 | 4290 | 0.001 | 0.004 | 0.33 | Atrial premature depolarization |
| C5 | K409 | 3787 | 4290 | 0.021 | 0.059 | 0.35 | Unilateral or unspecified inguinal hernia, without obstruction or gangrene |
| C5 | M235 | 269 | 4290 | 0.002 | 0.004 | 0.39 | Chronic instability of knee |
| C5 | M539 | 274 | 4290 | 0.002 | 0.004 | 0.44 | Dorsopathy, unspecified |
| C5 | I479 | 614 | 4290 | 0.004 | 0.009 | 0.44 | Paroxysmal tachycardia, unspecified |
| C5 | C509 | 1103 | 4290 | 0.008 | 0.017 | 0.45 | Malignant neoplasm: Breast, unspecified |
| C5 | N508 | 300 | 4290 | 0.002 | 0.005 | 0.45 | Other specified disorders of male genital organs |
| C6 | N180 | 454 | 3589 | 0.001 | 0.007 | 0.12 | Chronic kidney disease |
| C6 | C679 | 292 | 3589 | 0.001 | 0.005 | 0.12 | Malignant neoplasm: Bladder, unspecified |
| C6 | N199 | 711 | 3589 | 0.001 | 0.011 | 0.12 | Unspecified kidney failure |
| C6 | I489 | 7075 | 3589 | 0.015 | 0.110 | 0.14 | Atrial fibrillation and atrial flutter, unspecified |
| C6 | J441 | 1678 | 3589 | 0.004 | 0.026 | 0.14 | Chronic obstructive pulmonary disease with acute exacerbation, unspecified |
| C6 | J180 | 257 | 3589 | 0.001 | 0.004 | 0.14 | Bronchopneumonia, unspecified |
| C6 | C509 | 1103 | 3589 | 0.003 | 0.017 | 0.15 | Malignant neoplasm: Breast, unspecified |
| C6 | J960 | 837 | 3589 | 0.002 | 0.013 | 0.15 | Acute respiratory failure |
| C6 | J440 | 743 | 3589 | 0.002 | 0.012 | 0.17 | Chronic obstructive pulmonary disease with acute lower respiratory infection |
| C6 | K409 | 3787 | 3589 | 0.010 | 0.059 | 0.17 | Unilateral or unspecified inguinal hernia, without obstruction or gangrene |
| C7 | E107 | 583 | 3309 | 0.000 | 0.009 | 0.03 | Type 1 diabetes mellitus: With multiple complications |
| C7 | E103 | 576 | 3309 | 0.000 | 0.009 | 0.03 | Type 1 diabetes mellitus: With ophthalmic complications |
| C7 | N180 | 454 | 3309 | 0.000 | 0.007 | 0.04 | Chronic kidney disease |
| C7 | E104 | 434 | 3309 | 0.000 | 0.007 | 0.04 | Type 1 diabetes mellitus: With neurological complications |
| C7 | I709 | 433 | 3309 | 0.000 | 0.007 | 0.04 | Generalized and unspecified atherosclerosis |
| C7 | E105 | 416 | 3309 | 0.000 | 0.007 | 0.05 | Type 1 diabetes mellitus: With peripheral circulatory complications |
| C7 | E102 | 377 | 3309 | 0.000 | 0.006 | 0.05 | Type 1 diabetes mellitus: With renal complications |
| C7 | N189 | 1094 | 3309 | 0.001 | 0.017 | 0.05 | Chronic kidney disease, unspecified |
| C7 | J180 | 257 | 3309 | 0.000 | 0.004 | 0.07 | Bronchopneumonia, unspecified |
| C7 | E115 | 551 | 3309 | 0.001 | 0.009 | 0.11 | Type 2 diabetes mellitus: With peripheral circulatory complications |
| C8 | E103 | 576 | 2802 | 0.000 | 0.009 | 0.04 | Type 1 diabetes mellitus: With ophthalmic complications |
| C8 | E108 | 1073 | 2802 | 0.001 | 0.017 | 0.09 | Type 1 diabetes mellitus: With unspecified complications |
| C8 | E104 | 434 | 2802 | 0.001 | 0.007 | 0.11 | Type 1 diabetes mellitus: With neurological complications |
| C8 | E105 | 416 | 2802 | 0.001 | 0.006 | 0.11 | Type 1 diabetes mellitus: With peripheral circulatory complications |

| Clst | ICD-10 | # code | # clst | Obs | Exp | O/E-ratio | description |
|---|---|---|---|---|---|---|---|
| C8 | E115 | 551 | 2802 | 0.001 | 0.009 | 0.13 | Type 2 diabetes mellitus: With peripheral circulatory complications |
| C8 | M512 | 366 | 2802 | 0.001 | 0.006 | 0.13 | Other specified intervertebral disc displacement |
| C8 | E162 | 510 | 2802 | 0.001 | 0.008 | 0.14 | Hypoglycaemia, unspecified |
| C8 | E109 | 2680 | 2802 | 0.006 | 0.041 | 0.15 | Type 1 diabetes mellitus: Without complications |
| C8 | N180 | 454 | 2802 | 0.001 | 0.007 | 0.15 | Chronic kidney disease |
| C8 | E107 | 583 | 2802 | 0.001 | 0.009 | 0.16 | Type 1 diabetes mellitus: With multiple complications |
| C9 | M539 | 274 | 2581 | 0.000 | 0.004 | 0.09 | Dorsopathy, unspecified |
| C9 | D179 | 257 | 2581 | 0.000 | 0.004 | 0.10 | Benign lipomatous neoplasm, unspecified |
| C9 | D251 | 256 | 2581 | 0.000 | 0.004 | 0.10 | Intramural leiomyoma of uterus |
| C9 | D303 | 600 | 2581 | 0.001 | 0.009 | 0.13 | Benign neoplasm: Bladder |
| C9 | R072 | 367 | 2581 | 0.001 | 0.006 | 0.14 | Precordial pain |
| C9 | M512 | 366 | 2581 | 0.001 | 0.006 | 0.14 | Other specified intervertebral disc displacement |
| C9 | G439 | 463 | 2581 | 0.001 | 0.007 | 0.16 | Migraine, unspecified |
| C9 | M653 | 738 | 2581 | 0.002 | 0.011 | 0.17 | Trigger finger |
| C9 | M511 | 3357 | 2581 | 0.009 | 0.052 | 0.17 | Lumbar and other intervertebral disc disorders with radiculopathy |
| C9 | M431 | 583 | 2581 | 0.002 | 0.009 | 0.17 | Spondylolisthesis |
| C10 | A630 | 320 | 2562 | 0.000 | 0.005 | 0.08 | Anogenital (venereal) warts |
| C10 | K298 | 256 | 2562 | 0.000 | 0.004 | 0.10 | Duodenitis |
| C10 | I999 | 383 | 2562 | 0.001 | 0.006 | 0.13 | Other and unspecified disorders of circulatory system |
| C10 | F172 | 349 | 2562 | 0.001 | 0.005 | 0.14 | Mental and behavioural disorders due to use of tobacco: Dependence syndrome |
| C10 | M771 | 492 | 2562 | 0.001 | 0.008 | 0.16 | Lateral epicondylitis |
| C10 | N479 | 462 | 2562 | 0.001 | 0.007 | 0.16 | Redundant prepuce, phimosis and paraphimosis |
| C10 | N180 | 454 | 2562 | 0.001 | 0.007 | 0.17 | Chronic kidney disease |
| C10 | I309 | 297 | 2562 | 0.001 | 0.005 | 0.17 | Acute pericarditis, unspecified |
| C10 | I830 | 293 | 2562 | 0.001 | 0.005 | 0.17 | Varicose veins of lower extremities with ulcer |
| C10 | I802 | 280 | 2562 | 0.001 | 0.004 | 0.18 | Phlebitis and thrombophlebitis of other deep vessels of lower extremities |
| C11 | C509 | 1103 | 2292 | 0.000 | 0.017 | 0.03 | Malignant neoplasm: Breast, unspecified |
| C11 | H360 | 1467 | 2292 | 0.001 | 0.023 | 0.04 | Diabetic retinopathy |
| C11 | E113 | 720 | 2292 | 0.000 | 0.011 | 0.04 | Type 2 diabetes mellitus: With ophthalmic complications |
| C11 | E107 | 583 | 2292 | 0.000 | 0.009 | 0.05 | Type 1 diabetes mellitus: With multiple complications |
| C11 | E103 | 576 | 2292 | 0.000 | 0.009 | 0.05 | Type 1 diabetes mellitus: With ophthalmic complications |
| C11 | E104 | 434 | 2292 | 0.000 | 0.007 | 0.06 | Type 1 diabetes mellitus: With neurological complications |
| C11 | E105 | 416 | 2292 | 0.000 | 0.006 | 0.07 | Type 1 diabetes mellitus: With peripheral circulatory complications |
| C11 | E112 | 761 | 2292 | 0.001 | 0.012 | 0.07 | Type 2 diabetes mellitus: With renal complications |
| C11 | D249 | 758 | 2292 | 0.001 | 0.012 | 0.07 | Benign neoplasm of breast |
| C11 | E108 | 1073 | 2292 | 0.001 | 0.017 | 0.08 | Type 1 diabetes mellitus: With unspecified complications |
| C12 | C509 | 1103 | 2213 | 0.000 | 0.017 | 0.03 | Malignant neoplasm: Breast, unspecified |
| C12 | N921 | 875 | 2213 | 0.000 | 0.013 | 0.03 | Excessive and frequent menstruation with irregular cycle |
| C12 | E103 | 576 | 2213 | 0.000 | 0.009 | 0.05 | Type 1 diabetes mellitus: With ophthalmic complications |
| C12 | N924 | 561 | 2213 | 0.000 | 0.009 | 0.05 | Excessive bleeding in the premenopausal period |

| Clst | ICD-10 | # code | # clst | Obs | Exp | O/E-ratio | description |
|---|---|---|---|---|---|---|---|
| C12 | E148 | 483 | 2213 | 0.000 | 0.007 | 0.06 | Unspecified diabetes mellitus: With unspecified complications |
| C12 | N920 | 948 | 2213 | 0.001 | 0.015 | 0.06 | Excessive and frequent menstruation with regular cycle |
| C12 | F419 | 335 | 2213 | 0.000 | 0.005 | 0.09 | Anxiety disorder, unspecified |
| C12 | R064 | 542 | 2213 | 0.001 | 0.008 | 0.11 | Hyperventilation |
| C12 | D251 | 256 | 2213 | 0.000 | 0.004 | 0.12 | Intramural leiomyoma of uterus |
| C12 | N832 | 489 | 2213 | 0.001 | 0.008 | 0.12 | Other and unspecified ovarian cysts |
| C13 | E112 | 761 | 2070 | 0.000 | 0.012 | 0.04 | Type 2 diabetes mellitus: With renal complications |
| C13 | E102 | 377 | 2070 | 0.000 | 0.006 | 0.08 | Type 1 diabetes mellitus: With renal complications |
| C13 | N179 | 377 | 2070 | 0.000 | 0.006 | 0.08 | Acute renal failure, unspecified |
| C13 | D509 | 459 | 2070 | 0.001 | 0.007 | 0.14 | Iron deficiency anaemia, unspecified |
| C13 | H330 | 418 | 2070 | 0.001 | 0.006 | 0.15 | Retinal detachment with retinal break |
| C13 | J819 | 545 | 2070 | 0.001 | 0.008 | 0.17 | NA |
| C13 | E162 | 510 | 2070 | 0.001 | 0.008 | 0.19 | Hypoglycaemia, unspecified |
| C13 | E117 | 980 | 2070 | 0.003 | 0.015 | 0.19 | Type 2 diabetes mellitus: With multiple complications |
| C13 | I501 | 1502 | 2070 | 0.005 | 0.023 | 0.21 | Left ventricular failure |
| C13 | C679 | 292 | 2070 | 0.001 | 0.004 | 0.22 | Malignant neoplasm: Bladder, unspecified |
| C14 | H360 | 1467 | 2040 | 0.002 | 0.022 | 0.11 | Diabetic retinopathy |
| C14 | E103 | 576 | 2040 | 0.001 | 0.009 | 0.17 | Type 1 diabetes mellitus: With ophthalmic complications |
| C14 | E105 | 416 | 2040 | 0.001 | 0.006 | 0.23 | Type 1 diabetes mellitus: With peripheral circulatory complications |
| C14 | G510 | 365 | 2040 | 0.001 | 0.006 | 0.26 | Bell’s palsy |
| C14 | H431 | 334 | 2040 | 0.001 | 0.005 | 0.29 | Vitreous haemorrhage |
| C14 | E113 | 720 | 2040 | 0.003 | 0.011 | 0.31 | Type 2 diabetes mellitus: With ophthalmic complications |
| C14 | M519 | 604 | 2040 | 0.003 | 0.009 | 0.32 | Intervertebral disc disorder, unspecified |
| C14 | L905 | 295 | 2040 | 0.001 | 0.004 | 0.33 | Scar conditions and fibrosis of skin |
| C14 | E107 | 583 | 2040 | 0.003 | 0.009 | 0.33 | Type 1 diabetes mellitus: With multiple complications |
| C14 | H438 | 291 | 2040 | 0.001 | 0.004 | 0.33 | Other disorders of vitreous body |
| C15 | J439 | 317 | 2013 | 0.000 | 0.005 | 0.10 | Emphysema, unspecified |
| C15 | C679 | 292 | 2013 | 0.000 | 0.004 | 0.11 | Malignant neoplasm: Bladder, unspecified |
| C15 | K510 | 283 | 2013 | 0.000 | 0.004 | 0.12 | Ulcerative (chronic) pancolitis |
| C15 | I495 | 482 | 2013 | 0.001 | 0.007 | 0.14 | Sick sinus syndrome |
| C15 | K045 | 333 | 2013 | 0.001 | 0.005 | 0.20 | Chronic apical periodontitis |
| C15 | R570 | 320 | 2013 | 0.001 | 0.005 | 0.20 | Cardiogenic shock |
| C15 | R590 | 305 | 2013 | 0.001 | 0.005 | 0.21 | Localized enlarged lymph nodes |
| C15 | N180 | 454 | 2013 | 0.001 | 0.007 | 0.22 | Chronic kidney disease |
| C15 | D303 | 600 | 2013 | 0.002 | 0.009 | 0.22 | Benign neoplasm: Bladder |
| C15 | N133 | 267 | 2013 | 0.001 | 0.004 | 0.24 | Other and unspecified hydronephrosis |
| C16 | E149 | 958 | 1654 | 0.001 | 0.015 | 0.04 | Unspecified diabetes mellitus: Without complications |
| C16 | E114 | 881 | 1654 | 0.001 | 0.013 | 0.04 | Type 2 diabetes mellitus: With neurological complications |
| C16 | E103 | 576 | 1654 | 0.001 | 0.009 | 0.07 | Type 1 diabetes mellitus: With ophthalmic complications |
| C16 | H360 | 1467 | 1654 | 0.002 | 0.022 | 0.08 | Diabetic retinopathy |
| C16 | E105 | 416 | 1654 | 0.001 | 0.006 | 0.10 | Type 1 diabetes mellitus: With peripheral circulatory complications |
| C16 | E102 | 377 | 1654 | 0.001 | 0.006 | 0.10 | Type 1 diabetes mellitus: With renal complications |

| Clst | ICD-10 | # code | # clst | Obs | Exp | O/E-ratio | description |
|---|---|---|---|---|---|---|---|
| C16 | E108 | 1073 | 1654 | 0.002 | 0.016 | 0.11 | Type 1 diabetes mellitus: With unspecified complications |
| C16 | H431 | 334 | 1654 | 0.001 | 0.005 | 0.12 | Vitreous haemorrhage |
| C16 | E117 | 980 | 1654 | 0.002 | 0.015 | 0.12 | Type 2 diabetes mellitus: With multiple complications |
| C16 | E107 | 583 | 1654 | 0.001 | 0.009 | 0.14 | Type 1 diabetes mellitus: With multiple complications |
| C17 | N199 | 711 | 1281 | 0.001 | 0.011 | 0.07 | Unspecified kidney failure |
| C17 | E162 | 510 | 1281 | 0.001 | 0.008 | 0.10 | Hypoglycaemia, unspecified |
| C17 | E105 | 416 | 1281 | 0.001 | 0.006 | 0.12 | Type 1 diabetes mellitus: With peripheral circulatory complications |
| C17 | I469 | 372 | 1281 | 0.001 | 0.006 | 0.14 | Cardiac arrest, unspecified |
| C17 | D303 | 600 | 1281 | 0.002 | 0.009 | 0.17 | Benign neoplasm: Bladder |
| C17 | E107 | 583 | 1281 | 0.002 | 0.009 | 0.18 | Type 1 diabetes mellitus: With multiple complications |
| C17 | E103 | 576 | 1281 | 0.002 | 0.009 | 0.18 | Type 1 diabetes mellitus: With ophthalmic complications |
| C17 | K580 | 576 | 1281 | 0.002 | 0.009 | 0.18 | Irritable bowel syndrome with diarrhoea |
| C17 | K650 | 264 | 1281 | 0.001 | 0.004 | 0.20 | Acute peritonitis |
| C17 | D179 | 257 | 1281 | 0.001 | 0.004 | 0.20 | Benign lipomatous neoplasm, unspecified |
| C18 | J450 | 401 | 1251 | 0.001 | 0.006 | 0.13 | Predominantly allergic asthma |
| C18 | H521 | 366 | 1251 | 0.001 | 0.006 | 0.14 | Myopia |
| C18 | J350 | 315 | 1251 | 0.001 | 0.005 | 0.17 | Chronic tonsillitis |
| C18 | N924 | 561 | 1251 | 0.002 | 0.008 | 0.19 | Excessive bleeding in the premenopausal period |
| C18 | K402 | 280 | 1251 | 0.001 | 0.004 | 0.19 | Bilateral inguinal hernia, without obstruction or gangrene |
| C18 | N840 | 546 | 1251 | 0.002 | 0.008 | 0.19 | Polyp of corpus uteri |
| C18 | M235 | 269 | 1251 | 0.001 | 0.004 | 0.20 | Chronic instability of knee |
| C18 | M234 | 258 | 1251 | 0.001 | 0.004 | 0.20 | Loose body in knee |
| C18 | N832 | 489 | 1251 | 0.002 | 0.007 | 0.22 | Other and unspecified ovarian cysts |
| C18 | K800 | 468 | 1251 | 0.002 | 0.007 | 0.23 | Calculus of gallbladder with acute cholecystitis |
| C19 | C509 | 1103 | 1168 | 0.002 | 0.017 | 0.10 | Malignant neoplasm: Breast, unspecified |
| C19 | D649 | 1637 | 1168 | 0.003 | 0.025 | 0.10 | Anaemia, unspecified |
| C19 | F103 | 518 | 1168 | 0.001 | 0.008 | 0.11 | Mental and behavioural disorders due to use of alcohol: Withdrawal state |
| C19 | K810 | 505 | 1168 | 0.001 | 0.008 | 0.11 | Acute cholecystitis |
| C19 | N811 | 974 | 1168 | 0.002 | 0.015 | 0.12 | Cystocele |
| C19 | I429 | 479 | 1168 | 0.001 | 0.007 | 0.12 | Cardiomyopathy, unspecified |
| C19 | I709 | 433 | 1168 | 0.001 | 0.007 | 0.13 | Generalized and unspecified atherosclerosis |
| C19 | N390 | 1219 | 1168 | 0.003 | 0.018 | 0.14 | Urinary tract infection, site not specified |
| C19 | R001 | 394 | 1168 | 0.001 | 0.006 | 0.14 | Bradycardia, unspecified |
| C19 | E871 | 393 | 1168 | 0.001 | 0.006 | 0.14 | Hypo-osmolality and hyponatraemia |
| C20 | N920 | 948 | 1119 | 0.001 | 0.014 | 0.06 | Excessive and frequent menstruation with regular cycle |
| C20 | N921 | 875 | 1119 | 0.001 | 0.013 | 0.07 | Excessive and frequent menstruation with irregular cycle |
| C20 | G442 | 667 | 1119 | 0.001 | 0.010 | 0.09 | Tension-type headache |
| C20 | N924 | 561 | 1119 | 0.001 | 0.008 | 0.10 | Excessive bleeding in the premenopausal period |
| C20 | D279 | 435 | 1119 | 0.001 | 0.007 | 0.14 | NA |
| C20 | E104 | 434 | 1119 | 0.001 | 0.007 | 0.14 | Type 1 diabetes mellitus: With neurological complications |
| C20 | M224 | 423 | 1119 | 0.001 | 0.006 | 0.14 | Chondromalacia patellae |
| C20 | R104 | 391 | 1119 | 0.001 | 0.006 | 0.15 | Other and unspecified abdominal pain |
| C20 | E041 | 369 | 1119 | 0.001 | 0.006 | 0.16 | Nontoxic single thyroid nodule |

| Clst | ICD-10 | # code | # clst | Obs | Exp | O/E-ratio | description |
|---|---|---|---|---|---|---|---|
| C20 | M512 | 366 | 1119 | 0.001 | 0.006 | 0.16 | Other specified intervertebral disc displacement |
| C21 | J960 | 837 | 1000 | 0.001 | 0.013 | 0.08 | Acute respiratory failure |
| C21 | E107 | 583 | 1000 | 0.001 | 0.009 | 0.11 | Type 1 diabetes mellitus: With multiple complications |
| C21 | E103 | 576 | 1000 | 0.001 | 0.009 | 0.12 | Type 1 diabetes mellitus: With ophthalmic complications |
| C21 | I493 | 575 | 1000 | 0.001 | 0.009 | 0.12 | Ventricular premature depolarization |
| C21 | I495 | 482 | 1000 | 0.001 | 0.007 | 0.14 | Sick sinus syndrome |
| C21 | N180 | 454 | 1000 | 0.001 | 0.007 | 0.15 | Chronic kidney disease |
| C21 | R091 | 418 | 1000 | 0.001 | 0.006 | 0.16 | Pleurisy |
| C21 | E789 | 417 | 1000 | 0.001 | 0.006 | 0.16 | Disorder of lipoprotein metabolism, unspecified |
| C21 | E871 | 393 | 1000 | 0.001 | 0.006 | 0.17 | Hypo-osmolality and hyponatraemia |
| C21 | E102 | 377 | 1000 | 0.001 | 0.006 | 0.18 | Type 1 diabetes mellitus: With renal complications |
| C22 | F100 | 1212 | 988 | 0.001 | 0.018 | 0.06 | Mental and behavioural disorders due to use of alcohol: Acute intoxication |
| C22 | L979 | 605 | 988 | 0.001 | 0.009 | 0.11 | Ulcer of lower limb, not elsewhere classified |
| C22 | I429 | 479 | 988 | 0.001 | 0.007 | 0.14 | Cardiomyopathy, unspecified |
| C22 | N180 | 454 | 988 | 0.001 | 0.007 | 0.15 | Chronic kidney disease |
| C22 | M224 | 423 | 988 | 0.001 | 0.006 | 0.16 | Chondromalacia patellae |
| C22 | J448 | 396 | 988 | 0.001 | 0.006 | 0.17 | Other specified chronic obstructive pulmonary disease |
| C22 | E102 | 377 | 988 | 0.001 | 0.006 | 0.18 | Type 1 diabetes mellitus: With renal complications |
| C22 | I469 | 372 | 988 | 0.001 | 0.006 | 0.18 | Cardiac arrest, unspecified |
| C22 | R570 | 320 | 988 | 0.001 | 0.005 | 0.21 | Cardiogenic shock |
| C22 | L400 | 319 | 988 | 0.001 | 0.005 | 0.21 | Psoriasis vulgaris |
| C23 | F103 | 518 | 935 | 0.001 | 0.008 | 0.14 | Mental and behavioural disorders due to use of alcohol: Withdrawal state |
| C23 | M191 | 474 | 935 | 0.001 | 0.007 | 0.15 | Post-traumatic arthrosis of other joints |
| C23 | R490 | 415 | 935 | 0.001 | 0.006 | 0.17 | Dysphonia |
| C23 | C619 | 1173 | 935 | 0.003 | 0.018 | 0.18 | Malignant neoplasm of prostate |
| C23 | I999 | 383 | 935 | 0.001 | 0.006 | 0.18 | Other and unspecified disorders of circulatory system |
| C23 | E041 | 369 | 935 | 0.001 | 0.006 | 0.19 | Nontoxic single thyroid nodule |
| C23 | H659 | 368 | 935 | 0.001 | 0.006 | 0.19 | Nonsuppurative otitis media, unspecified |
| C23 | I480 | 364 | 935 | 0.001 | 0.005 | 0.20 | Paroxysmal atrial fibrillation |
| C23 | H908 | 631 | 935 | 0.002 | 0.010 | 0.22 | Mixed conductive and sensorineural hearing loss, unspecified |
| C23 | I839 | 1522 | 935 | 0.005 | 0.023 | 0.23 | Varicose veins of lower extremities without ulcer or inflammation |
| C24 | N409 | 3319 | 932 | 0.002 | 0.050 | 0.04 | Hyperplasia of prostate |
| C24 | C619 | 1173 | 932 | 0.001 | 0.018 | 0.06 | Malignant neoplasm of prostate |
| C24 | E108 | 1073 | 932 | 0.001 | 0.016 | 0.07 | Type 1 diabetes mellitus: With unspecified complications |
| C24 | H833 | 1412 | 932 | 0.002 | 0.021 | 0.10 | Noise effects on inner ear |
| C24 | E107 | 583 | 932 | 0.001 | 0.009 | 0.12 | Type 1 diabetes mellitus: With multiple complications |
| C24 | M109 | 547 | 932 | 0.001 | 0.008 | 0.13 | Gout, unspecified |
| C24 | E116 | 477 | 932 | 0.001 | 0.007 | 0.15 | Type 2 diabetes mellitus: With other specified complications |
| C24 | M191 | 474 | 932 | 0.001 | 0.007 | 0.15 | Post-traumatic arthrosis of other joints |
| C24 | H109 | 465 | 932 | 0.001 | 0.007 | 0.15 | Conjunctivitis, unspecified |
| C24 | N484 | 456 | 932 | 0.001 | 0.007 | 0.16 | Impotence of organic origin |
| C25 | H938 | 1116 | 860 | 0.001 | 0.017 | 0.07 | Other specified disorders of ear |
| C25 | H360 | 1467 | 860 | 0.002 | 0.022 | 0.10 | Diabetic retinopathy |
| C25 | N200 | 1391 | 860 | 0.002 | 0.021 | 0.11 | Calculus of kidney |

| Clst | ICD-10 | # code | # clst | Obs | Exp | O/E-ratio | description |
|---|---|---|---|---|---|---|---|
| C25 | H911 | 3527 | 860 | 0.007 | 0.053 | 0.13 | Presbycusis |
| C25 | C619 | 1173 | 860 | 0.002 | 0.018 | 0.13 | Malignant neoplasm of prostate |
| C25 | E107 | 583 | 860 | 0.001 | 0.009 | 0.13 | Type 1 diabetes mellitus: With multiple complications |
| C25 | E103 | 576 | 860 | 0.001 | 0.009 | 0.13 | Type 1 diabetes mellitus: With ophthalmic complications |
| C25 | I493 | 575 | 860 | 0.001 | 0.009 | 0.13 | Ventricular premature depolarization |
| C25 | H919 | 4610 | 860 | 0.010 | 0.069 | 0.15 | Hearing loss, unspecified |
| C25 | H833 | 1412 | 860 | 0.003 | 0.021 | 0.16 | Noise effects on inner ear |
| C26 | E112 | 761 | 852 | 0.001 | 0.011 | 0.10 | Type 2 diabetes mellitus: With renal complications |
| C26 | N409 | 3319 | 852 | 0.006 | 0.050 | 0.12 | Hyperplasia of prostate |
| C26 | C619 | 1173 | 852 | 0.002 | 0.018 | 0.13 | Malignant neoplasm of prostate |
| C26 | I350 | 2664 | 852 | 0.006 | 0.040 | 0.15 | Aortic (valve) stenosis |
| C26 | I714 | 517 | 852 | 0.001 | 0.008 | 0.15 | Abdominal aortic aneurysm, without mention of rupture |
| C26 | E162 | 510 | 852 | 0.001 | 0.008 | 0.15 | Hypoglycaemia, unspecified |
| C26 | H360 | 1467 | 852 | 0.004 | 0.022 | 0.16 | Diabetic retinopathy |
| C26 | I495 | 482 | 852 | 0.001 | 0.007 | 0.16 | Sick sinus syndrome |
| C26 | K800 | 468 | 852 | 0.001 | 0.007 | 0.17 | Calculus of gallbladder with acute cholecystitis |
| C26 | N180 | 454 | 852 | 0.001 | 0.007 | 0.17 | Chronic kidney disease |
| C27 | H360 | 1467 | 823 | 0.001 | 0.022 | 0.06 | Diabetic retinopathy |
| C27 | M170 | 2145 | 823 | 0.004 | 0.032 | 0.11 | Primary gonarthrosis, bilateral |
| C27 | D303 | 600 | 823 | 0.001 | 0.009 | 0.14 | Benign neoplasm: Bladder |
| C27 | C509 | 1103 | 823 | 0.002 | 0.017 | 0.15 | Malignant neoplasm: Breast, unspecified |
| C27 | J209 | 527 | 823 | 0.001 | 0.008 | 0.15 | Acute bronchitis, unspecified |
| C27 | J320 | 513 | 823 | 0.001 | 0.008 | 0.16 | Chronic maxillary sinusitis |
| C27 | E162 | 510 | 823 | 0.001 | 0.008 | 0.16 | Hypoglycaemia, unspecified |
| C27 | L022 | 506 | 823 | 0.001 | 0.008 | 0.16 | Cutaneous abscess, furuncle and carbuncle of trunk |
| C27 | K810 | 505 | 823 | 0.001 | 0.008 | 0.16 | Acute cholecystitis |
| C27 | M179 | 2242 | 823 | 0.006 | 0.034 | 0.18 | Gonarthrosis, unspecified |
| C28 | M546 | 977 | 686 | 0.001 | 0.015 | 0.10 | Pain in thoracic spine |
| C28 | D249 | 758 | 686 | 0.001 | 0.011 | 0.13 | Benign neoplasm of breast |
| C28 | E113 | 720 | 686 | 0.001 | 0.011 | 0.14 | Type 2 diabetes mellitus: With ophthalmic complications |
| C28 | E669 | 2114 | 686 | 0.004 | 0.032 | 0.14 | Obesity, unspecified |
| C28 | R002 | 691 | 686 | 0.001 | 0.010 | 0.14 | Palpitations |
| C28 | K359 | 678 | 686 | 0.001 | 0.010 | 0.14 | NA |
| C28 | H350 | 548 | 686 | 0.001 | 0.008 | 0.18 | Background retinopathy and retinal vascular changes |
| C28 | I119 | 534 | 686 | 0.001 | 0.008 | 0.18 | Hypertensive heart disease without (congestive) heart failure |
| C28 | J209 | 527 | 686 | 0.001 | 0.008 | 0.18 | Acute bronchitis, unspecified |
| C28 | E162 | 510 | 686 | 0.001 | 0.008 | 0.19 | Hypoglycaemia, unspecified |
| C29 | M171 | 2940 | 550 | 0.002 | 0.044 | 0.04 | Other primary gonarthrosis |
| C29 | I489 | 7075 | 550 | 0.005 | 0.106 | 0.05 | Atrial fibrillation and atrial flutter, unspecified |
| C29 | H919 | 4610 | 550 | 0.004 | 0.069 | 0.05 | Hearing loss, unspecified |
| C29 | I702 | 2251 | 550 | 0.002 | 0.034 | 0.05 | Atherosclerosis of arteries of extremities |
| C29 | M179 | 2242 | 550 | 0.002 | 0.034 | 0.05 | Gonarthrosis, unspecified |
| C29 | M170 | 2145 | 550 | 0.002 | 0.032 | 0.06 | Primary gonarthrosis, bilateral |
| C29 | G459 | 2066 | 550 | 0.002 | 0.031 | 0.06 | Transient cerebral ischaemic attack, unspecified |
| C29 | I639 | 1989 | 550 | 0.002 | 0.030 | 0.06 | Cerebral infarction, unspecified |
| C29 | G473 | 1897 | 550 | 0.002 | 0.028 | 0.06 | Sleep apnoea |
| C29 | N409 | 3319 | 550 | 0.004 | 0.050 | 0.07 | Hyperplasia of prostate |

| Clst | ICD-10 | # code | # clst | Obs | Exp | O/E-ratio | description |
|---|---|---|---|---|---|---|---|
| C30 | F100 | 1212 | 533 | 0.002 | 0.018 | 0.10 | Mental and behavioural disorders due to use of alcohol: Acute intoxication |
| C30 | N393 | 827 | 533 | 0.002 | 0.012 | 0.15 | Stress incontinence |
| C30 | M503 | 520 | 533 | 0.002 | 0.008 | 0.24 | Other cervical disc degeneration |
| C30 | R065 | 983 | 533 | 0.004 | 0.015 | 0.26 | Mouth breathing |
| C30 | M546 | 977 | 533 | 0.004 | 0.015 | 0.26 | Pain in thoracic spine |
| C30 | R490 | 415 | 533 | 0.002 | 0.006 | 0.30 | Dysphonia |
| C30 | M060 | 398 | 533 | 0.002 | 0.006 | 0.32 | Seronegative rheumatoid arthritis |
| C30 | F102 | 1189 | 533 | 0.006 | 0.018 | 0.32 | Mental and behavioural disorders due to use of alcohol: Dependence syndrome |
| C30 | J448 | 396 | 533 | 0.002 | 0.006 | 0.32 | Other specified chronic obstructive pulmonary disease |
| C30 | C509 | 1103 | 533 | 0.006 | 0.017 | 0.34 | Malignant neoplasm: Breast, unspecified |
| C31 | F102 | 1189 | 520 | 0.002 | 0.018 | 0.11 | Mental and behavioural disorders due to use of alcohol: Dependence syndrome |
| C31 | C619 | 1173 | 520 | 0.002 | 0.018 | 0.11 | Malignant neoplasm of prostate |
| C31 | K439 | 981 | 520 | 0.002 | 0.015 | 0.13 | Other and unspecified ventral hernia without obstruction or gangrene |
| C31 | E117 | 980 | 520 | 0.002 | 0.015 | 0.13 | Type 2 diabetes mellitus: With multiple complications |
| C31 | E112 | 761 | 520 | 0.002 | 0.011 | 0.17 | Type 2 diabetes mellitus: With renal complications |
| C31 | I351 | 696 | 520 | 0.002 | 0.010 | 0.18 | Aortic (valve) insufficiency |
| C31 | N200 | 1391 | 520 | 0.004 | 0.021 | 0.18 | Calculus of kidney |
| C31 | E118 | 2669 | 520 | 0.008 | 0.040 | 0.19 | Type 2 diabetes mellitus: With unspecified complications |
| C31 | M519 | 604 | 520 | 0.002 | 0.009 | 0.21 | Intervertebral disc disorder, unspecified |
| C31 | E107 | 583 | 520 | 0.002 | 0.009 | 0.22 | Type 1 diabetes mellitus: With multiple complications |
